# Effectiveness of COVID-19 vaccines against hospitalization and death in Canada: A multiprovincial test-negative design study

**DOI:** 10.1101/2022.04.13.22273825

**Authors:** Sharifa Nasreen, Yossi Febriani, Héctor Alexander Velásquez García, Geng Zhang, Mina Tadrous, Sarah A. Buchan, Christiaan H. Righolt, Salaheddin M. Mahmud, Naveed Zafar Janjua, Mel Krajden, Gaston De Serres, Jeffrey C. Kwong, the Canadian Immunization Research Network (CIRN) Provincial Collaborative Network (PCN) Investigators

## Abstract

**Background:** A major goal of COVID-19 vaccination is to prevent severe outcomes (hospitalizations and deaths). We estimated the effectiveness of mRNA and ChAdOx1 COVID-19 vaccines against severe outcomes in four Canadian provinces between December 2020 and September 2021.

**Methods:** We conducted this multiprovincial retrospective test-negative study among community-dwelling adults aged ≥18 years in Ontario, Quebec, British Columbia, and Manitoba using linked provincial databases and a common study protocol. Multivariable logistic regression was used to estimate province-specific vaccine effectiveness against COVID-19 hospitalization and/or death. Estimates were pooled using random effects models.

**Results:** We included 2,508,296 tested subjects, with 31,776 COVID-19 hospitalizations and 5,842 deaths. Vaccine effectiveness was 83% after a first dose, and 98% after a second dose, against both hospitalization and death (separately). Against severe outcomes (hospitalization or death), effectiveness was 87% (95%CI: 71%–94%) ≥84 days after a first dose of mRNA vaccine, increasing to 98% (95%CI: 96%–99%) ≥112 days after a second dose. Vaccine effectiveness against severe outcomes for ChAdOx1 was 88% (95%CI: 75%–94%) ≥56 days after a first dose, increasing to 97% (95%CI: 91%–99%) ≥56 days after a second dose. Lower one-dose effectiveness was observed for adults aged ≥80 years and those with comorbidities, but effectiveness became comparable after a second dose. Two doses of vaccines provided very high protection for both homologous and heterologous schedules, and against Alpha, Gamma, and Delta variants.

**Conclusions:** Two doses of mRNA or ChAdOx1 vaccines provide excellent protection against severe outcomes of hospitalization and death.

## INTRODUCTION

SARS-CoV-2 infection is associated with high morbidity and mortality [1]. A major goal of COVID-19 vaccination is to prevent hospitalizations and deaths. Provincial COVID-19 vaccination programs in Canada have involved extended intervals between first and second doses due to vaccine supply constraints, and use of heterologous (i.e., ‘mix-and-match’) vaccine schedules due to concerns regarding vaccine-induced immune thrombotic thrombocytopenia associated with ChAdOx1 (AstraZeneca Vaxzevria and COVISHIELD) and variable supplies of specific vaccine products [2, 3].

Assessing COVID-19 vaccine effectiveness (VE) against severe outcomes with longer follow-up after each dose will inform our understanding of the duration of protection. There are limited data on real-world effectiveness of heterologous vaccine schedules and extended dosing intervals against severe outcomes [4]. The aim of this study was to estimate the effectiveness of mRNA (BNT162b2 [Pfizer-BioNTech Comirnaty] and mRNA-1273 [Moderna Spikevax]) and ChAdOx1 vaccines against COVID-19 hospitalizations and deaths, including longer follow-up periods, heterologous vaccine schedules, and extended dosing intervals.

## METHODS

### Study design, setting, and population

Using a common study protocol across 4 Canadian provinces, we conducted a test-negative design study [5] involving Ontario, Quebec, British Columbia (BC), and Manitoba (total population 30 million, comprising approximately 79% of the Canadian population) among community-dwelling residents who sought SARS-CoV-2 testing. We included all residents aged ≥18 years, eligible for provincial health insurance (virtually everyone), not living in long-term care, and tested for SARS-CoV-2 between the start of vaccine availability in a province (14 December 2020 in Ontario and Quebec, 15 December 2020 in BC, 16 December 2020 in Manitoba) and 30 September 2021 and met our case or control definitions. We excluded recipients of non-Health Canada-authorized vaccines or Ad26.COV2.S (Johnson & Johnson’s Janssen) vaccine.

### Data sources and definitions

We linked data from provincial SARS-CoV-2 laboratory testing, COVID-19 public health surveillance, COVID-19 vaccination, and health administrative datasets using unique encoded identifiers in each province at: ICES (Ontario), Institut National de Santé Publique du Québec, BC Centre for Disease Control, and the University of Manitoba Vaccine and Drug Evaluation Centre (Supplemental Tables S1 and S2).

#### Outcomes

Our primary outcome was COVID-19 hospitalization or death identified from notifiable disease reporting systems and/or other administrative databases. COVID-19 hospitalization was defined as hospitalization or ICU admission with a positive SARS-CoV-2 test within 14 days prior to or 3 days after hospitalization. We excluded nosocomial cases flagged in notifiable disease reporting systems and SARS-CoV-2-positive cases with specimen collection >3 days after hospital admission. COVID-19 death was defined as death with a recent positive SARS-CoV-2 test identified from notifiable disease reporting systems or deaths occurring within 30 days following a positive SARS-CoV-2 test or within 7 days post-mortem. Subjects with COVID-19 hospitalizations and deaths were treated as test-positive cases using the earliest of the specimen collection date, hospitalization date, or death date as the index date. We included COVID-19 hospitalizations and deaths occurring until 30 September 2021, and included only the first positive test. Symptomatic subjects who tested negative during the study period were treated as test-negative controls using the specimen collection date as the index date. For controls with multiple negative tests, we randomly selected one symptomatic test-negative specimen collection date. SARS-CoV-2 lineage was determined using whole genome sequencing or screening PCR tests for various mutations to group test-positive specimens into the following mutually exclusive categories: Alpha, Beta, Gamma, Beta/Gamma, Delta, and non-VOC SARS-CoV-2 (Supplemental methods).

#### COVID-19 vaccination

Information on COVID-19 vaccination, including vaccine product, date of administration, and dose number were collected from each province’s COVID-19 vaccination information system.

#### Covariates

Information on the following covariates were obtained from relevant data sources [6-8]: age group, sex, geographic region (Supplemental Table S3), 2-week periods of test (to control for temporal changes in virus circulation and vaccine uptake), number of RT-PCR tests during the 3 months prior to the start of the study (as a proxy for frequently tested at-risk individuals), comorbidities that increase the risk of severe COVID-19 [9], receipt of 2019–2020 and/or 2020– 2021 influenza vaccination (as a proxy for health behaviours), and 4 area-level social determinants of health (median neighbourhood income, proportion of the working population employed as non-health essential workers [i.e., those unable to work from home], average number of persons per dwelling, and proportion of the population who self-identify as a visible minority) [6]. All covariates were measured as of the start of the study period, except week of SARS-CoV-2 test.

### Statistical analyses

Baseline characteristics were summarized as means (standard deviation, SD) for continuous variables and frequencies and percentages for categorical variables. Logistic regression models were used to estimate crude and adjusted odds ratios (OR) comparing the odds of being vaccinated versus unvaccinated between test-positive cases and test-negative controls separately in each province. Adjusted models accounted for all the covariates listed above.

We estimated overall ORs separately for hospitalization and death but for all vaccines combined ≥14 days after a first dose among those who had received only 1 dose at the time of testing and ≥7 days after a second dose among those who had received 2 doses. We also estimated the ORs by time since their most recent dose for mRNA vaccines and ChAdOx1separately; follow-up periods were shorter after ChAdOx1 than mRNA vaccines because there were fewer recipients of ChAdOx1. We conducted subgroup analysis by subject characteristics (age group, sex, and presence of any comorbidity), vaccine product, and SARS-CoV-2 lineage. We also estimated ORs for varying dosing intervals among individuals who received 2 doses of mRNA vaccines.

Each province conducted these analyses independently to estimate province-specific ORs. There were some variations in data sources and analyses among the provinces. Details are provided in Supplemental methods. We conducted a sensitivity analysis by also including hospitalizations and deaths from administrative databases in Ontario.

#### Meta-analyses

We pooled the log OR estimates from each province using random-effects models with inverse variance weighting [10]. We used random-effects models because provinces differed slightly in population demographics and vaccination programs that may introduce some variability. We converted province-specific and pooled ORs to VE using the formula: VE=(1–OR)*100. We assessed between-province heterogeneity using the *I*^*2*^ statistic. Pooled VE estimates were not presented if a single province contributed to the meta-analysis. Meta-analyses were conducted using meta package in R version 4.1.2 [11].

## RESULTS

Overall, we included 2,508,296 community-dwelling SARS-CoV-2-tested subjects (Table 1). We identified 33,420 COVID-19-associated severe outcomes; receipt of at least 1 dose of a COVID-19 vaccine ranged from 13% to 20% among test-positive severe outcome cases, and from 40% to 46% among symptomatic test-negative controls (Supplemental Table S4). Cases were more likely to be older, male, have had no SARS-CoV-2 tests during the 3 months before the vaccination program, have a comorbidity, have received an influenza vaccine (in Ontario and Quebec), and more likely to reside in neighbourhoods with lower income/more material deprivation, more people per dwelling, and greater proportions of essential workers (in Ontario and BC), and greater proportions of visible minorities than controls. Vaccinated subjects were more likely to be older, have a comorbidity, have received an influenza vaccine, and less likely to be male than unvaccinated subjects (Supplemental Table S5). Most vaccinated subjects received BNT162b2 (Supplemental Table S6).

**Table 1:** Baseline characteristics of study subjects in Ontario, Quebec, British Columbia, and Manitoba

| Characteristic | Ontario,<br>n (%)<sup>a</sup><br>(N=557,220) | Quebec,<br>n (%)<sup>a</sup><br>(N=954,208) | British Columbia,<br>n (%)<sup>a</sup><br>(N=876,397) | Manitoba,<br>n (%)<sup>a</sup><br>(N=120,471) |
| --- | --- | --- | --- | --- |
| **BNT162b2 Pfizer-BioNTech Comirnaty** |  |  |  |  |
| ≥14 days after dose 1 | 58,315 (10.5) | 124,274 (13.0) | 101,851 (11.6) | 34,622 (28.7) |
| ≥7 days after dose 2 | 92,771 (16.6) | 151,722 (15.9) | 138,192 (15.8) | 18,061 (15.0) |
| Interval between 2 doses (days), median (IQR) | 56 (38, 75) | 70 (58, 91) | 63 (55, 76) | 45 (22, 64) |
| **mRNA-1273 Moderna Spikevax** |  |  |  |  |
| ≥14 days after dose 1 | 15,196 (2.7) | 32,377 (3.4) | 28,452 (3.2) | 12,480 (10.4) |
| ≥7 days after dose 2 | 27,798 (5.0) | 41,415 (4.3) | 35,915 (4.1) | 7,832 (6.5) |
| Interval between 2 doses(days), median (IQR) | 48 (35, 62) | 63 (56, 78) | 61 (52, 72) | 38 (31, 46) |
| **ChAdOx1 AstraZeneca Vaxzevria<sup>b</sup>** |  |  |  |  |
| ≥14 days after dose 1 | 5,071 (0.9) | 9689 (1.0) | 8,938 (1.0) | 5,916 (4.9) |
| ≥7 days after dose 2 | 1,219 (0.2) | 3699 (0.4) | 4,590 (0.5) | 275 (0.2) |
| Interval between 2 doses(days), median (IQR) | 65 (59, 72) | 59 (55, 65) | 60 (56, 63) | 61 (38, 71) |
| **ChAdOx1 COVISHIELD** |  |  |  |  |
| ≥14 days after dose 1 | 2,554 (0.5) | 3,911 (0.4) | 3,957 (0.5) | - |
| ≥7 days after dose 2 | 33 (0.0) | 10 (0.0) | 263 (0.0) | - |
| Interval between 2 doses(days), median (IQR) | 76 (38, 81) | 71 (57, 77) | 66 (57, 71) | - |
| Age (years), mean (standard deviation) | 44 (18) | 47 (17) | 45 (18) | 44 (17) |
| Age group (years) |  |  |  |  |
| 18–29 | 141,488 (25.4) | 175,744 (18.4) | 210,248 (24.0) | 30,711 (25.5) |
| 30–39 | 128,416 (23.0) | 217,384 (22.8) | 197,183 (22.5) | 28,476 (23.6) |
| 40–49 | 92,740 (16.6) | 185,032 (19.4) | 143,403 (16.4) | 20,902 (17.4) |
| 50–59 | 80,799 (14.5) | 138,724 (14.5) | 123,970 (14.1) | 16,635 (13.8) |
| 60–69 | 58,508 (10.5) | 126,632 (13.3) | 99,396 (11.3) | 12,891 (10.7) |
| 70–79 | 33,004 (5.9) | 74,199 (7.8) | 62,240 (7.1) | 6,931 (5.8) |
| ≥80 | 22,265 (4.0) | 36,493 (3.8) | 39,957 (4.6) | 3,925 (3.3) |
| Male sex | 237,038 (42.5) | 383,234 (40.2) | 394,672 (45.0) | 51,780 (43.0) |
| Biweekly period of test |  |  |  |  |
| 14 Dec 2020 to 27 Dec 2020 | 27,460 (4.9) | 41,106 (4.3) | 45,972 (5.2) | 5,710 (4.7) |
| 28 Dec 2020 to 10 Jan 2021 | 26,092 (4.7) | 40,043 (4.2) | 44,566 (5.1) | 4,957 (4.1) |
| 11 Jan 2021 to 24 Jan 2021 | 25,458 (4.6) | 42,003 (4.4) | 25,951 (3.0) | 5,087 (4.2) |
| 25 Jan 2021 to 7 Feb 2021 | 22,759 (4.1) | 42,192 (4.4) | 18,848 (2.2) | 4,894 (4.1) |
| 8 Feb 2021 to 21 Feb 2021 | 22,836 (4.1) | 48,113 (5.0) | 46,936 (5.4) | 4,527 (3.8) |
| 22 Feb 2021 to 7 Mar 2021 | 28,004 (5.0) | 49,602 (5.2) | 43,207 (4.9) | 4,874 (4.0) |
| 8 Mar 2021 to 21 Mar 2021 | 30,121 (5.4) | 51,785 (5.4) | 46,212 (5.3) | 5,017 (4.2) |
| 22 Mar 2021 to 4 Apr 2021 | 34,060 (6.1) | 63,205 (6.6) | 43,409 (5.0) | 5,651 (4.7) |
| 5 Apr 2021 to 18 Apr 2021 | 42,157 (7.6) | 73,382 (7.7) | 51,216 (5.8) | 7,295 (6.1) |
| 19 Apr 2021 to 2 May 2021 | 39,790 (7.1) | 61,902 (6.5) | 63,033 (7.2) | 9,091 (7.5) |
| 3 May 2021 to 16 May 2021 | 29,579 (5.3) | 55,483 (5.8) | 54,286 (6.2) | 8,282 (6.9) |
| 17 May 2021 to 30 May 2021 | 18,593 (3.3) | 38,257 (4.0) | 43,828 (5.0) | 7,355 (6.1) |
| 31 May 2021 to 13 Jun 2021 | 15,137 (2.7) | 33,893 (3.6) | 32,440 (3.7) | 5,726 (4.8) |
| 14 Jun 2021 to 27 Jun 2021 | 14,502 (2.6) | 41,399 (4.3) | 26,637 (3.0) | 4,178 (3.5) |
| 28 Jun 2021 to 11 Jul 2021 | 12,106 (2.2) | 36,821 (3.9) | 23,321 (2.7) | 3,036 (2.5) |
| 12 Jul 2021 to 25 Jul 2021 | 15,486 (2.8) | 28,982 (3.0) | 18,290 (2.1) | 3,153 (2.6) |
| 26 Jul 2021 to 8 Aug 2021 | 22,823 (4.1) | 28,143 (2.9) | 19,734 (2.3) | 3,938 (3.3) |
| 9 Aug 2021 to 22 Aug 2021 | 30,381 (5.5) | 30,868 (3.2) | 27,137 (3.1) | 5,012 (4.2) |
| 23 Aug 2021 to 5 Sep 2021 | 32,022 (5.7) | 38,606 (4.0) | 41,806 (4.8) | 6,741 (5.6) |
| 6 Sep 2021 to 19 Sep 2021 | 32,792 (5.9) | 54,933 (5.8) | 53,469 (6.1) | 7,739 (6.4) |
| 20 Sep 2021 to 30 Sep 2021 | 35,062 (6.3) | 53,490 (5.6) | 106,099 (12.1) | 8,208 (6.8) |
| Number of tests in previous 3 months |  |  |  |  |
| 0 | 406,271 (72.9) | 714,551 (74.9) | 740,569 (84.5) | 89,782 (74.5) |
| 1 | 105,529 (18.9) | 171,300 (18.0) | 102,832 (11.7) | 24,934 (20.7) |
| ≥2 | 45,420 (8.2) | 68,357 (7.2) | 32,996 (3.8) | 5,755 (4.8) |
| Any comorbidity<sup>c</sup> | 262,241 (47.1) | 307,907 (32.3) | 330,599 (37.7) | 47,103 (39.1) |
| Receipt of 2019-2020 and/or 2020-2021<br>influenza vaccination | 185,440 (33.3) | 260,925 (27.3) | N/A | 56,247 (46.7) |
| Neighbourhood income quintile<sup>d</sup> |  |  |  |  |
| 1 (lowest) | 100,810 (18.1) | 166,800 (17.5) | 111,788 (12.8) | 21,938 (18.2) |
| 2 | 108,090 (19.4) | 185,658 (19.5) | 151,657 (17.3) | 23,308 (19.3) |
| 3 | 111,753 (20.1) | 196,837 (20.6) | 165,278 (18.9) | 23,230 (19.3) |
| 4 | 114,904 (20.6) | 203,589 (21.3) | 187,753 (21.4) | 23,117 (19.2) |
| 5 (highest) | 119,128 (21.4) | 201,324 (21.1) | 170,276 (19.4) | 24,129 (20.0) |
| Unknown/missing | 2,535 (0.5) | - | 89,645 (10.2) | 4,749 (3.9) |
| Essential workers quintile<sup>e</sup> |  |  |  |  |
| 1 (0%–32.5%) | 103,249 (18.5) | 220,241 (23.1) | 95,159 (10.9) | 25,333 (21.0) |
| 2 (32.5%–42.3%) | 126,153 (22.6) | 218,661 (22.9) | 161,535 (18.4) | 27,984 (23.2) |
| 3 (42.3%–49.8%) | 115,880 (20.8) | 195,285 (20.5) | 152,624 (17.4) | 23,107 (19.2) |
| 4 (50.0%–57.5%) | 108,902 (19.5) | 171,272 (17.9) | 137,267 (15.7) | 22,571 (18.7) |
| 5 (57.5%–100%) | 99,179 (17.8) | 148,749 (15.6) | 124,483 (14.2) | 19,598 (16.3) |
| Unknown/missing | 3,857 (0.7) | - | 205,329 (23.4) | 385 (0.3) |
| Persons per dwelling quintile<sup>f</sup> |  |  |  |  |
| 1 (0–2.1) | 101,530 (18.2) | 192,060 (20.1) | 181,303 (20.7) | 30,301 (25.2) |
| 2 (2.2–2.4) | 100,405 (18.0) | 143,369 (15.0) | 147,314 (16.8) | 19,583 (16.3) |
| 3 (2.5–2.6) | 71,933 (12.9) | 165,670 (17.4) | 152,321 (17.4) | 20,084 (16.7) |
| 4 (2.7–3.0) | 133,095 (23.9) | 239,881 (25.1) | 158,033 (18.0) | 25,418 (21.1) |
| 5 (3.1–5.7) | 146,240 (26.2) | 213,228 (22.3) | 186,204 (21.2) | 23,207 (19.3) |
| Unknown/missing | 4,017 (0.7) | - | 51,222 (5.8) | 385 (0.3) |
| Self-identified visible minority quintile<sup>g</sup> |  |  |  |  |
| 1 (0.0%–2.2%) | 93,149 (16.7) | 223,858 (23.5) | 100,472 (11.5) | 18,289 (15.2) |
| 2 (2.2%–7.5%) | 102,423 (18.4) | 148,256 (15.5) | 147,284 (16.8) | 23,194 (19.3) |
| 3 (7.5%–18.7%) | 101,805 (18.3) | 218,744 (22.9) | 178,848 (20.4) | 24,346 (20.2) |
| 4 (18.7%–43.5%) | 114,781 (20.6) | 199,297 (20.9) | 204,378 (23.3) | 26,293 (21.8) |
| 5 (43.5%–100%) | 141,214 (25.3) | 164,053 (17.2) | 195,193 (22.3) | 26,471 (22.0) |
| Unknown/missing | 3,848 (0.7) | - | 50,222 (5.7) | 385 (0.3) |
| SARS-CoV-2 cases with severe outcomes | 17,437 (3.1) | 7,854 (0.8) | 5,928 (0.7) | 2,201 (1.8) |
| SARS-CoV-2 lineage for those testing positive<sup>h</sup> |  |  |  |  |
| Non-VOC | 5,312 (30.5) | - | 519 (8.8) | 995 (45.2) |
| Alpha (B.1.1.7) | 7,033 (40.3) | 1,575 (20.1) | 869 (14.7) | 783 (35.6) |
| Beta/Gamma (B.1.351 or P.1) | 226 (1.3) | - | 227 (3.8) | 22 (1.0) |
| Beta (B.1.351) | 166 (1.0) | - | 5 (0.1) | 8 (0.4) |
| Gamma (P.1) | 382 (2.2) | - | 678 (11.4) | 14 (0.6) |
| Delta (B.1.617.2) | 1,684 (9.7) | 585 (7.4) | 1,257 (21.2) | 107 (4.9) |
| Unspecified | - | - | - | 294 (13.4) |

<sup>a</sup>Proportion reported, unless stated otherwise.
<sup>b</sup>AstraZeneca Vaxzevria and COVISHIELD vaccines reported only as ChAdOx1 in Manitoba.
<sup>c</sup>Comorbidities include chronic respiratory diseases, chronic heart diseases, hypertension, diabetes, immunocompromising conditions due to underlying diseases or therapy, autoimmune diseases, chronic kidney disease, advanced liver disease, dementia/frailty and history of stroke or transient ischemic attack.
<sup>d</sup>Neighbourhood income quintile has variable cut-off values in each city/Census area to account for cost of living. A dissemination area (DA) being in quintile 1 means it is among the lowest 20% of DAs in its city by income. Material deprivation index quintile used in British Columbia; quintile 1 represents 'most deprived' and quintile 5 represents 'least deprived'.
<sup>e</sup>Percentage of people in the area working in the following occupations: sales and service occupations; trades, transport and equipment operators and related occupations; natural resources, agriculture, and related production occupations; and occupations in manufacturing and utilities. Census counts for people are randomly rounded up or down to the nearest number divisible by 5, which causes some minor imprecision.
<sup>f</sup>Range of persons per dwelling.
<sup>g</sup>Percentage of people in the area who self-identified as a visible minority. Census counts for people are randomly rounded up or down to the nearest number divisible by 5, which causes some minor imprecision.
<sup>h</sup>Proportions calculated using the total number of SARS-CoV-2 cases with severe outcomes as the denominator.

### Vaccine effectiveness

In pooled analyses, the adjusted VE (aVE) was 83% (95% confidence interval [CI]: 78%–87%) against hospitalization and 83% (95%CI: 72%–90%) against death after a first dose; aVE increased to 98% against both hospitalization (95%CI: 96%–99%) and death (95%CI: 95%– 99%) after receiving a second dose (Figure 1, Supplemental Table S7).

**Figure 1:**
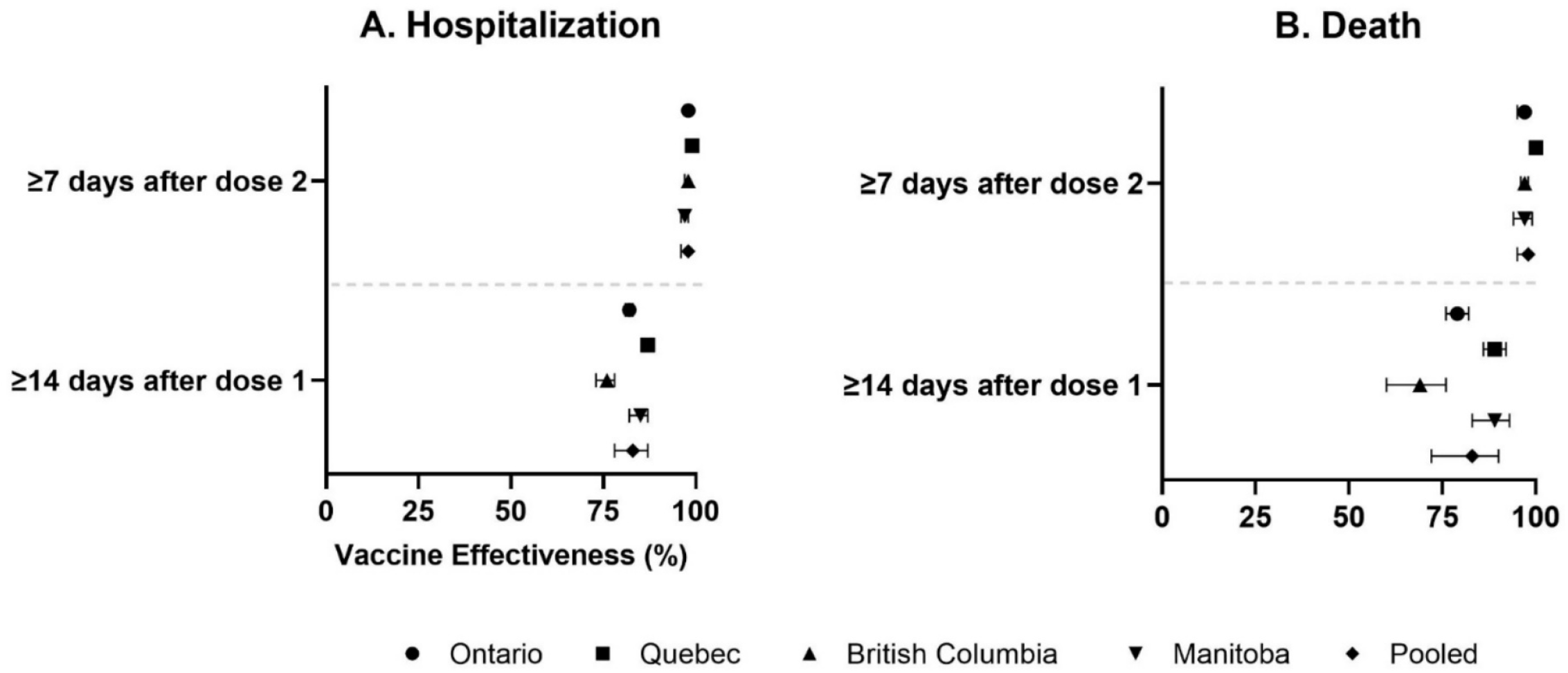
Province-specific and pooled adjusted vaccine effectiveness ≥14 days after a first dose and ≥7 days after receiving a second dose against hospitalization (panel A) and death (panel B) in Ontario, Quebec, British Columbia, and Manitoba.

Against severe outcomes of hospitalization or death, the pooled aVE increased over time from 43% (95%CI: 25%–57%) 0–13 days after a first dose to 87% (95%CI: 71%–94%) ≥84 days after a first dose for an mRNA vaccine; after receiving a second dose, pooled aVE increased from 93% (95%CI: 88%–96%) at 0–6 days to 98% (95%CI: 96%–99%) at ≥112 days (Figure 2A, Supplemental Table 7). The pooled aVE against severe outcomes for ChAdOx1 increased from 37% (95%CI: 20%–51%) 0–13 days after a first dose to 88% (95%CI: 75%–94%) ≥56 days after a first dose; aVE increased to 97% (95%CI: 91%–99%) ≥56 days after receiving a second dose (Figure 2B, Supplemental Table S8).

**Figure 2:**
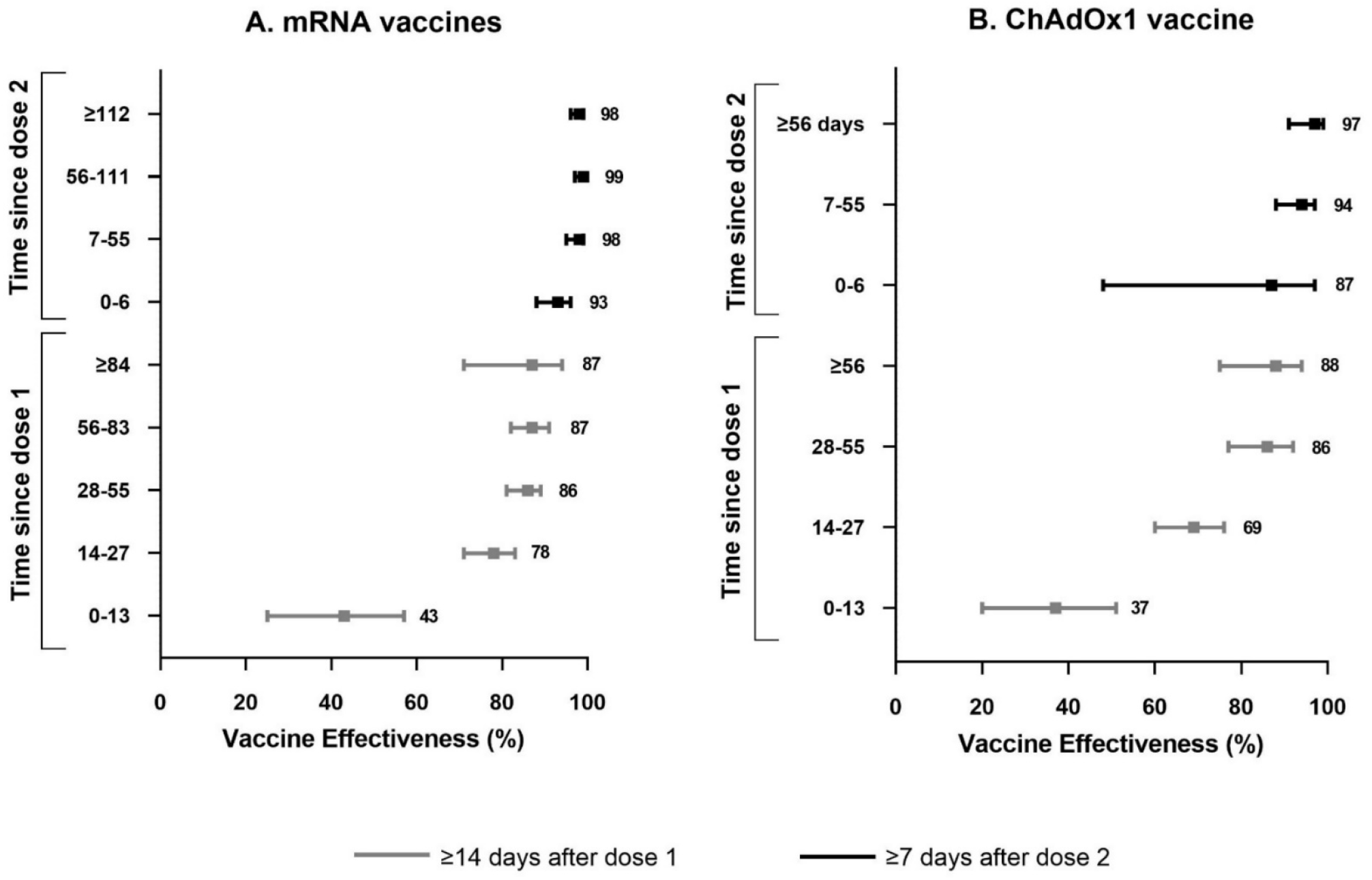
Pooled adjusted vaccine effectiveness against severe outcomes of hospitalization or death for mRNA (panel A) and ChAdOx1 (panel B) vaccines in Ontario, Quebec, British Columbia, and Manitoba.

In subgroup analyses, the pooled aVE against severe outcomes was lower for adults aged ≥80 years versus younger adults aged 18–59 years, and in subjects with comorbidities versus those without comorbidities ≥14 days after receiving a first dose; however, aVE became comparable across all subgroups ≥7 days after receipt of a second dose (Figure 3A, Supplemental Table S9). The pooled aVE against severe outcomes was >80% ≥14 days after a first dose for BNT162b2, mRNA-1273, or ChAdOx1 vaccines, which increased to ≥97% ≥7 days after receipt of a second dose. aVE against severe outcomes was similar ≥7 days after receiving a second dose of a mixed mRNA or ChAdOx1/mRNA mixed schedule (Figure 3B, Supplemental Table S10). aVE against severe outcomes caused by VOCs was lowest against Beta at 61% and highest against Delta at 89% ≥14 days after a first dose, and aVE increased to ≥97% against Alpha, Gamma, and Delta ≥7 days after a second dose (Figure 3C, Supplemental Table S11).

**Figure 3:**
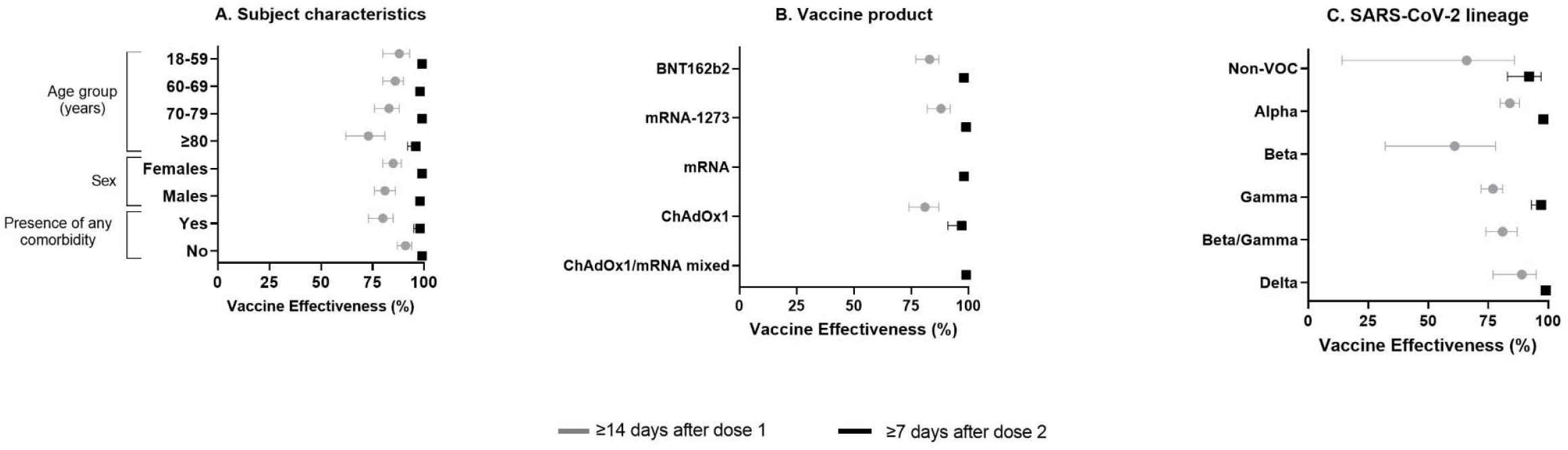
Pooled adjusted vaccine effectiveness against severe outcomes of hospitalization or death ≥14 days after a first dose and ≥7 days after receiving a second dose by subject characteristics (panel A), vaccine product (panel B) and SARS-CoV-2 lineage (panel C) in Ontario, Quebec, British Columbia, and Manitoba.

The pooled aVE against severe outcomes for mRNA vaccines 7–55 days after a second dose increased from 94% with a dosing interval of 21–34 days to ≥98% with a longer dosing interval, although 95% confidence intervals for aVE overlapped (Figure 4, Supplemental Table S12). aVE was maintained at ≥97% with longer between dose intervals from 56 days through ≥112 days after receiving a second dose.

**Figure 4:**
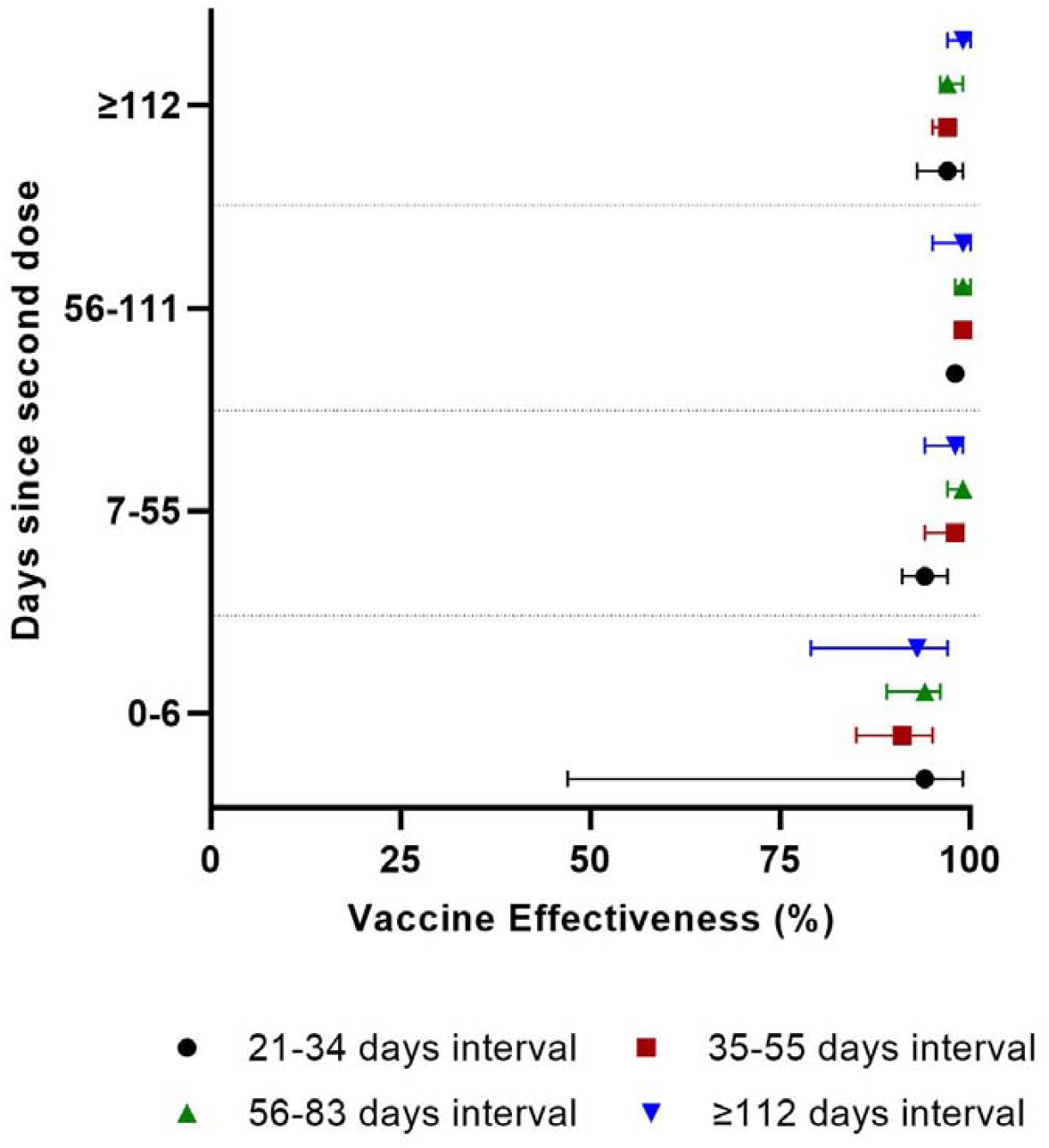
Pooled adjusted vaccine effectiveness against severe outcomes of hospitalization or death for subjects who received two doses of an mRNA vaccine by various intervals between vaccine doses and time since the second dose in Ontario, Quebec, British Columbia, and Manitoba.

Although heterogeneity between the provinces was observed, as reflected by the *I*^*2*^ statistics for most of the models (Supplemental Table S13), all province-specific VE estimates suggest the vaccines were significantly protective with some variation in the magnitude.

In sensitivity analyses with the inclusion of severe outcomes from administrative data in Ontario, we identified 22,759 severe outcomes; pooled sensitivity analyses yielded similar VE estimates to those obtained from the pooled primary analyses (Supplemental Table S14).

## DISCUSSION

In this study, we found high and very high vaccine effectiveness against hospitalization and death with 1 (83%) and 2 (98%) doses of COVID-19 vaccines, respectively. mRNA and ChAdOx1 vaccines had comparable effectiveness after first and second doses; protection increased or remained relatively stable over time after each dose without noticeable waning over this relatively short period of observation. In subgroup analyses, we observed lower one-dose VE for adults aged ≥80 years and those with comorbidities, but VE became comparable after receiving a second dose. Two doses of vaccines provided very high protection against Alpha, Gamma, and Delta variants. We observed very high level of protection with both homologous and heterologous schedules. Finally, our findings suggest that lengthening the dosing interval had minimal impact on VE against severe outcomes.

Our pooled aVE estimates against hospitalization and against death ≥14 days after receiving a single dose were higher than reported pooled VE estimates in a systematic review and meta-analysis of studies published up to 22 July 2021 (61% [95%CI: 41%–81%] against hospitalization, and 44% [95%CI: 23%–64%] against death) [12]. Our 1-dose VE estimates may have been higher due to a longer period of observation before receipt of a second dose, as VE may still be rising in the initial weeks post first dose receipt. Also, their VE estimates included other COVID-19 vaccines (e.g., CoronaVac) and different population groups (e.g., general population, health care workers, older adults, and long-term care or nursing home residents) without stratification by population subgroup. VE estimates ≥7 days after a second dose in that study (93% [95%CI: 84%–100%] against hospitalization, and 97% [95%CI: 95%–98%] against death) were comparable to our pooled aVE estimates. Another systematic review and meta-analysis that included published literature up to 25 August 2021 reported a pooled VE of 91% (95%CI: 85%–95%) and 94% (95%CI: 83%–98%) against hospitalization and a composite of severe outcomes due to Delta, respectively, after receipt of a second dose [13].

Against hospitalization or death, we observed sustained pooled aVE of 87% for mRNA vaccines at ≥12 weeks, and 88% for ChAdOx1 at ≥8 weeks with wider 95%CIs over time after a first dose. Similarly, pooled aVE of 98% at ≥16 weeks for mRNA vaccines and 97% at ≥8 weeks for ChAdOx1 vaccine after a second dose was observed. However, there were fewer vaccinated cases with longer follow-up compared to shorter follow-up, and very few subjects had an excessively long follow-up. Sustained VEs of 84–89% against hospitalizations, or hospitalizations and deaths up to 24 weeks were observed with 2 doses of an mRNA vaccine in the USA [14, 15] and Qatar [16]. A high VE of ≥95% at ≥28 weeks for mRNA vaccines and ≥93% at ≥12 weeks for ChAdOx1 was also maintained against hospitalizations in Quebec and BC [4]. However, some waning of immunity against hospitalizations and deaths after a second dose has been reported from other studies. VE against Delta variant-related hospitalization, and death decreased from 99% at 2–9 weeks to 92% at ≥20 weeks for BNT162b2 with more pronounced decline for ChAdOx1 from 95% at 2–9 weeks to 80–85% at ≥20 weeks in the UK [17]. Waning of protection against hospitalizations and deaths for BNT162b2 was observed at ≥7 months in Qatar [16]. VE against hospitalization for COVID-19 or all-cause 30-day mortality after confirmed infection for mRNA or ChAdOx1 declined from 89% (95%CI: 82%–93%) at 15–30 days to 64% (95%CI: 44% to 77%) at ≥121 days after a second dose in Sweden [18]. The cumulative VE within 6 months of receiving 2 doses of a COVID-19 vaccine (mainly mRNA and ChAdOx1) declined from 87% and 84% to 52% and 34% after 6 months against hospitalizations and deaths, respectively in Italy [19]. Confounding by indication resulting from averaging VE across subgroups with different exposure and infection risk, vaccination priority, clinical risk, and increased transmission and/or shorter interval of 3 weeks between doses with longer follow-up and rapid uptake of vaccines may explain the waning of VE observed in these studies [17, 20].

Our finding of comparable VE against severe outcomes in older and younger adults and in people with and without comorbidities after receiving a second dose aligns with findings from previous studies [6, 21-23]. However, a lower overall VE of 88% (95%CI: 82%–92%) was also reported previously in older adults aged ≥80 years compared to ≥94% VE in adults aged <80 years [4]. We found good overall protection against hospitalizations or deaths caused by Alpha and Delta (≥84%) ≥14 days after the first dose, and excellent protection (≥98%) ≥7 days after a second dose. Similar high VEs against Alpha (84%–97%) and Delta (92%–98%) with a second dose have been reported from other studies [24-27].

We observed similar high pooled aVE (≥97%) against severe outcomes ≥7 days after receiving a second dose of homologous BNT162b2, mRNA-1273, or ChAdOx1 vaccine series; these estimates were similar to our pooled aVE after receiving mixed mRNA (98%) or ChAdOx1/mRNA mixed schedule (99%), adding to the evidence of real-world effectiveness of heterologous dosing schedules. Our findings corroborate previously reported VE estimates against hospitalization using homologous and heterologous vaccine schedules from Quebec and BC [4]. Countries and jurisdictions with low 2-dose vaccine coverage and/or facing limited supplies of specific vaccine products could benefit from implementing heterologous vaccine schedules to increase population protection against severe outcomes.

We observed only a slight difference in VE between short and extended dosing intervals as reflected by only 4–5% higher VE with a dosing interval of ≥35 days compared to 21–34 days and 95%CIs overlapped. Persistently high VE was observed with longer follow-up across different dosing interval categories without evidence of considerable waning. Contrary to our findings, a previous study using data from Quebec and BC observed a higher VE against hospitalizations ≥14 days after 2 doses of mRNA vaccines with a dosing interval of 7–8 weeks (98% [95%CI: 97%–99%] and 99% [95%CI: 98%–99%], respectively) compared to a dosing interval of 3–4 weeks (87% [95%CI: 79%–92%] and 93% [95%CI: 87%–96%], respectively) [4]. This likely resulted from differences in methods and follow-up time between the studies. Deciding on the optimal interval between doses must weigh the benefits of delaying second doses against the risks of infection and subsequent severe outcomes in the context of local incidence, vaccine coverage, and vaccine supply.

This study has some limitations. First, while the test-negative design accounts for differences in healthcare seeking behaviour, indications for testing and risks of exposure to SARS-CoV-2 infection between test-positive cases and test-negative controls may differ. Testing indications also varied between the provinces and over the study period. We adjusted for biweekly period of test and number of prior tests to account for these. Second, although healthcare utilization and thresholds for hospitalization may vary between and within jurisdictions, hospital capacity was maintained to admit patients requiring hospitalization and we do not expect differential under-or over-estimation of severe outcomes, particularly death, with respect to COVID-19 vaccination status. Third, despite a common study protocol, there is likely heterogeneity among provinces in terms of differences in populations, vaccination programs (rollout logistics and priority groups), SARS-CoV-2 testing criteria, data capture, and covariates adjusted; we used random-effects models to account for statistical heterogeneity. Fourth, given the observational nature of the study, residual confounding remains possible despite adjustment for a number of potential confounders. Finally, our VE estimates may not apply to severe outcomes caused by Omicron.

In conclusion, our results from this large multiprovincial study provide strong evidence of excellent protection against severe outcomes of hospitalizations and deaths with 2 doses of mRNA or ChAdOx1 vaccines during the pre-Omicron period. With 2 doses, we found relatively stable protection through 16 weeks and beyond for mRNA vaccines and 8 weeks and beyond for ChAdOx1. Our findings further support the interchangeability of homologous and heterologous vaccine schedules. Likewise, the sustained protection from extended dosing intervals observed in our study lends evidence to delay administration of second doses in settings faced with limited vaccine supply.

## Data Availability

Data used in this article was derived from administrative health and social data as a secondary use. The dataset for Ontario from this study is held securely in coded form at ICES. While legal data sharing agreements between ICES and data providers (e.g., healthcare organizations and government) prohibit ICES from making the dataset publicly available, access may be granted to those who meet pre-specified criteria for confidential access, available at www.ices.on.ca/DAS. The full dataset creation plan and underlying analytic code are available from the authors upon request, understanding that the computer programs may rely upon coding templates or macros that are unique to ICES and are therefore either inaccessible or may require modification. The data was provided under specific data sharing agreements only for approved use at the Manitoba Centre for Health Policy (MCHP). The original source data is not owned by the researchers or MCHP and as such cannot be provided to a public repository. The original data source and approval for use has been noted in the acknowledgments of the article. Where necessary, source data specific to this article or project may be reviewed at MCHP with the consent of the original data providers, along with the required privacy and ethical review bodies.

## Contributors

JCK, SN, GDS, CHR, NZ, MT designed and oversaw the study. SN, YF, HAVG, and GZ conducted province-specific analyses. SN conducted the meta-analyses and drafted the manuscript. All authors contributed to the analysis plan, interpreted the results, critically reviewed and edited the manuscript, approved the final version, and agreed to be accountable for all aspects of the work.

## Ethics approval

ICES is a prescribed entity under Ontario’s Personal Health Information Protection Act (PHIPA). Section 45 of PHIPA authorizes ICES to collect personal health information, without consent, for the purpose of analysis or compiling statistical information with respect to the management of, evaluation or monitoring of, the allocation of resources to or planning for all or part of the health system. Projects that use data collected by ICES under section 45 of PHIPA, and use no other data, are exempt from REB review. The use of the data in this project is authorized under section 45 and approved by ICES’ Privacy and Legal Office.

## Disclaimers

This study was supported by ICES, which is funded by an annual grant from the Ontario Ministry of Health (MOH) and the Ministry of Long-Term Care (MLTC). This study was supported by the Ontario Health Data Platform (OHDP), a Province of Ontario initiative to support Ontario’s ongoing response to COVID-19 and its related impacts. The study sponsors did not participate in the design and conduct of the study; collection, management, analysis and interpretation of the data; preparation, review or approval of the manuscript; or the decision to submit the manuscript for publication. Parts of this material are based on data and/or information compiled and provided by the Canadian Institute for Health Information (CIHI) and by Ontario Health (OH). However, the analyses, conclusions, opinions and statements expressed herein are solely those of the authors, and do not reflect those of the funding or data sources; no endorsement by ICES, MOH, MLTC, OHDP, its partners, the Province of Ontario, CIHI or OH is intended or should be inferred. All inferences, opinions, and conclusions drawn in this manuscript are those of the authors, and do not reflect the opinions or policies of the Data Steward(s) in BC.

## Acknowledgments

We would like to acknowledge Public Health Ontario for access to case-level data from CCM, COVID-19 laboratory data, and COVaxON. We also thank the staff of Ontario’s public health units who are responsible for COVID-19 case and contact management and data collection within CCM. We thank IQVIA Solutions Canada Inc. for use of their Drug Information file. The authors are grateful to the residents of Ontario, Quebec, British Columbia and Manitoba without whom this research would be impossible. The authors acknowledge the Manitoba Centre for Health Policy for use of data contained in the Manitoba Population Research Data Repository under project # 2020-037 (HIPC # 2020/2021–04, REB # HS23859 (H2020:181)). The results and conclusions are those of the authors and no official endorsement by the Manitoba Centre for Health Policy, Manitoba Health, or other data providers is intended or should be inferred. Data used in this study are from the Manitoba Population Research Data Repository housed at the Manitoba Centre for Health Policy, University of Manitoba and were derived from data provided by Manitoba Health. We acknowledge the assistance of the Provincial Health Services Authority, BC Ministry of Health and Regional Health Authority staff involved in data access, procurement, and management. We gratefully acknowledge the residents of British Columbia whose data are integrated in the British Columbia COVID-19 Cohort (BCC19C).

