## Supplemental for "Effectiveness of COVID-19 vaccines against hospitalization and death in Canada: A multiprovincial test-negative design study"

**Supplemental Material**

**Supplemental methods**

*Extended dosing interval and heterologous COVID-19 vaccination policies in Canada:*

In view of limited vaccine supply, British Columbia (BC) and Quebec extended the interval between the first and second doses to six weeks and 12 weeks, respectively in January 2021 to maximize one-dose vaccine coverage [1]. Facing similar vaccine supply constraints, other provinces also delayed second dose administration by up to 16 weeks in accordance with a recommendation from Canada's National Advisory Committee on Immunization (NACI) in March 2021 [2]. In view of the safety signal of vaccine-induced immune thrombotic thrombocytopenia associated with ChAdOx1 (AstraZeneca Vaxzevria and COVISHIELD), NACI recommended the use of ChAdOx1 for adults aged ≥55 years in March 2021, and in April 2021 recommended ChAdOx1 for adults aged ≥30 years only when mRNA vaccines were contraindicated or inaccessible [3]. In June 2021, after reviewing the evidence on the interchangeability of authorized COVID-19 vaccines in Canada, NACI recommended that a different mRNA vaccine product could be used for the second dose if the initial mRNA vaccine product was unavailable; and either a ChAdOx1 or an mRNA vaccine product could be used for subsequent dose if the first dose was a ChAdOx1 vaccine [4]. Thereafter, jurisdictions in Canada started implementing heterologous or 'mix-and-match' vaccine schedules due to variable supplies of specific vaccine products.

*SARS-CoV-2 lineage ascertainment:*

SARS-CoV-2 lineage was ascertained from whole genome sequencing or screening tests for N501Y, E484K, K417N, and K417T mutations to group test-positive specimens into mutually exclusive categories. Specimens designated as the B.1.1.7 lineage or having N501Y and E484K mutations were considered Alpha; specimens designated as the B.1.351 lineage or having K417N, N501Y, and E484K mutations were considered Beta; specimens designated as the P.1 lineage or having K417T, N501Y, and E484K mutations were considered Gamma; specimens designated as the B.1.1.7 or P.1 lineage, or having N501Y and E484K mutations were considered as Beta/Gamma as these could not be separated into Beta or Gamma; and specimens designated as the B.1.617.2 lineage, and in Ontario specimens having N501Y and E484K mutations from geographic locations and dates when B.1.617.2 was known to be predominant through whole genome sequencing were considered Delta [5]. Specimens with earlier variant, or no lineage information, and absence of both N501Y and E484K mutations were considered non-VOC SARS-CoV-2. Information on non-VOC, Beta, and Gamma variants was not available in Quebec.

*Statistical analyses (province-specific variations)*:

In Quebec, information on comorbidity and area-level social determinants of health was missing for approximately 9% and 1% of the study-eligible subjects (n=1,069,031), respectively, and missing not at random. Thus, only subjects with information on all covariates were included in Quebec. Other provinces included subjects with missing covariate information in the study as a separate category. For subgroup analysis in Quebec, the model for 70–79 years, and Alpha and Delta variants did not converge when all covariates were included in the model and including the entire study period, respectively. To achieve model convergence, the model for 70–79 years was adjusted for sex, biweekly period of test, any comorbidity, number of SARS-CoV-2 tests in previous 3 months, and receipt of influenza vaccination in current or prior influenza season; analysis period was restricted from 28 December 2020 to 30 September 2021 for the Alpha variant and from 26 July 2021 to 30 September 2021 for the Delta variant in Quebec. The primary analysis was conducted by including severe outcomes identified from the provincial notifiable disease reporting system Public Health Case and Contact Management Solution (CCM) in Ontario, and severe outcomes from the COVID-19 surveillance data alone or from both COVID-19 surveillance and administrative data in other provinces.

**Supplemental Table S1: Data sources in Ontario, Quebec, British Columbia, and Manitoba**

| Category | Information | Province (2021 population, million) | | | |
|---|---|---|---|---|---|
| | | **Ontario (14.8)** | **Quebec (8.6)** | **British Columbia (5.2)** | **Manitoba (1.4)** |
| **Vaccine** | **COVID-19 vaccine** | COVaxON provincial COVID-19 vaccination management system | Vaccine registry | Provincial Immunization Registry | Public Health Information Management System (PHIMS) - COVID-19-specific database[a] |
| **COVID-19 Cases** | **Laboratory testing** | Ontario Laboratories Information System (OLIS) | Nosotech | Integrated lab data and Provincial Laboratory Information Solution (PLIS) | COVID-19 Lab Test Results data |
| | **Notifiable diseases (with course and outcome)** | Public Health Case and Contact Management Solution (CCM) | plateforme Trajectoire de santé publique | COVID-19 case surveillance dataset | PHIMS - Surveillance Case of COVID-19 |
| | **Mutations and SARS-CoV-2 lineages** | CCM | Nosotech | Integrated laboratory dataset for COVID-19 and PLIS | PHIMS - Surveillance Case of COVID-19 |
| **Outcomes** | **Death** | Reported with case data in CCM | plateforme Trajectoire de santé publique | Vital Statistics | PHIMS - Surveillance Case of COVID-19 |
| | | Administrative database: Registered Persons Database (RPDB) | - | COVID-19 case surveillance dataset | - |
| | **Hospitalization, including ICU admission** | Reported with case data in CCM | MedEcho | Provincial COVID-19 Monitoring Solution (PCMS)<br><br>COVID-19 case surveillance dataset | COVID hospitalization/ICU in surveillance data |
| | | Administrative database: Discharge Abstract | - | Discharge Abstract Database (DAD) | - |

| | | | | | |
|---|---|---|---|---|---|
| | | Database (DAD) | | | |
| **Covariates** | **Age** | RPDB | Vaccine registry | Integrated laboratory dataset for COVID-19 | Manitoba Health Population Registry Data |
| | **Sex** | RPDB | Vaccine registry | Integrated laboratory dataset for COVID-19 | Manitoba Health Population Registry Data |
| | **Public health unit region based on postal code of residence** | RPDB | Vaccine registry | Integrated laboratory dataset for COVID-19 | Manitoba Health Population Registry Data |
| | **Biweekly period of test** | OLIS | Nosotech | Integrated laboratory dataset for COVID-19 | COVID-19 Lab Test Results data |
| | **Number of tests in previous 3 months** | OLIS | Nosotech | Integrated laboratory dataset for COVID-19 | COVID-19 Lab Test Results data |
| | **Comorbidity** | Various registries and cohorts created using algorithms applied to health admin data [6] | Système intégré de surveillance des maladies chroniques du Québec (SISMACQ) | Various registries and cohorts created using algorithms applied to health admin data | Various registries and cohorts created using algorithms applied to health admin data |
| | **Influenza vaccination (physician billing claims)** | Ontario Health Insurance Plan (OHIP) | Vaccine registry | Not available | Manitoba Immunization Monitoring System (MIMS)/ Public Health Information Management System (PHIMS) |
| | **Influenza vaccination (pharmacist billing claims)** | Ontario Drug Benefit (ODB) | Vaccine registry | Not available | PHIMS |
| | **Median neighbourhood income** | 2016 Census | 2016 Census | Information on neighbourhood income not available; used Material Deprivation Index (average household income; unemployment rate; and high school education rate) from 2016 | 2016 Census |

| | | | | | |
|---|---|---|---|---|---|
| | | | | Census | |
| | **Proportion working in non-health essential services** | 2016 Census | 2016 Census | 2016 Census | 2016 Census |
| | **Persons per dwelling quintile** | 2016 Census | 2016 Census | 2016 Census | 2016 Census |
| | **Self-identified visible minority quintile** | 2016 Census | 2016 Census | 2016 Census | 2016 Census |

[a]AstraZeneca Vaxzevria and COVISHIELD vaccines were reported only as ChAdOx1

**Supplemental Table S2: Data Sources integrated within the BC COVID-19 Cohort (BCC19C)**

| British Columbia Centre for Disease Control (BCCDC), Provincial Heath Services Authority (PHSA) and Regional Health Authority data sources | Data Date Ranges |
|---|---|
| Integrated COVID-19 laboratory dataset (SARS-CoV2 tests from private/public labs) [7] | Jan,2020-onward |
| COVID-19 surveillance case data (information collected on all probable/confirmed cases as part of public health follow up) [8] | Jan,2020-onward |
| Provincial COVID-19 Monitoring Solution (critical and non-critical care hospital census data) [9] | Jan,2020-onward |
| Provincial Immunizations Registry (COVID-19 vaccination data) [10] | Dec,2020-onward |
| Provincial Laboratory Information Solution (laboratory tests from private/public labs) [11] | Jan,2020-onward |
| Public Health Reporting Data warehouse (Influenza laboratory tests) [12] | Jan,2008-onward |
| Emergency department visits (hospital-based and community-based ambulatory care) | Mar,2020-onward |
| **Ministry of Health (MoH) Administrative Data Sources** | **Data Date Ranges** |
| Client Roster (CR) (registry of enrollment in the universal public health insurance plan including residential history) [13] | 2008/9-onward |
| Discharge Abstracts Database (DAD) (hospital discharge records) [14] | 2008/9-onward |
| Medical Services Plan (MSP) (physician diagnostic and billing data for services provided through universal public health insurance plan)[15] | 2008/9-onward |
| PharmaNet (Pharma) (prescription drugs dispensed from community pharmacies, includes medications covered by public and private insurance plans) [16] | 2008/9-onward |
| BC Vital Statistics (VS) (deaths registry) [17] | 2008/9-onward |
| National Ambulatory Care Reporting System (NACRS) (hospital-based and community-based ambulatory care) [18] | 2011/12–onward |
| Chronic Disease Registry [19] | 2008/9-2018/19 |
| 811 Calls (respiratory calls only) [20] | 2014-onward |
| Health System Matrix [21] | 2018/19-onward |
| Population Grouper Methodology [22] | 2008/9-onward |

**Supplemental Table S3: Distribution of study subjects by geographic region in Ontario, Quebec, British Columbia, and Manitoba**

| Ontario | | Quebec | | British Columbia | | Manitoba | |
|---|---|---|---|---|---|---|---|
| **Public health unit region** | **n (%)** | **Heath region** | **n (%)** | **Health authority** | **n (%)** | **Regional health authority** | **n (%)** |
| Central East | 55,503 (10.0) | Bas-Saint-Laurent | 20,278 (2.1) | Fraser | 379,347 (43.3) | Interlake-Eastern | 11,779 (9.8) |
| Central West | 98,349 (17.6) | Saguenay – Lac-Saint-Jean | 35,083 (3.7) | Interior | 136,410 (15.6) | Northern | 8,146 (6.8) |
| Durham | 27,121 (4.9) | Capitale-Nationale | 110,311 (11.6) | Island | 110,506 (12.6) | Prairie Mountain | 16,606 (13.8) |
| Eastern | 26,841 (4.8) | Mauricie-et-Centre-du-Québec | 65,919 (6.9) | Northern | 33,098 (3.8) | Southern Health-Santé Sud | 12,485 (10.4) |
| North | 53,519 (9.6) | Estrie | 55,962 (5.9) | Vancouver Coastal | 199,179 (22.7) | Winnipeg | 71,097 (59.0) |
| Ottawa | 7,428 (1.3) | Montréal | 188,802 (19.8) | Unknown/ missing | 17,857 (2.0) | Public Trustee/ In CFS care | 357 (0.3) |
| Peel | 73,898 (13.3) | Outaouais | 37,842 (4.0) | | | | |
| South West | 67,253 (12.1) | Abitibi-Témiscamingue | 15,622 (1.6) | | | | |
| Toronto | 105,920 (19.0) | Côte-Nord | 9,099 (1.0) | | | | |
| York | 39,132 (7.0) | Nord-du-Québec | 1,281 (0.1) | | | | |
| Unknown/ missing | 2,256 (0.4) | Gaspésie – Îles-de-la-Madeleine | 9,807 (1.0) | | | | |
| | | Chaudière-Appalaches | 52,869 (5.5) | | | | |
| | | Laval | 44,321 (4.6) | | | | |
| | | Lanaudière | 66,461 (7.0) | | | | |
| | | Laurentides | 73,223 (7.7) | | | | |
| | | Montérégie | 165,813 (17.4) | | | | |
| | | Nunavik | 654 (0.1) | | | | |
| | | Terres-Cries-de-la-Baie-James | 861 (0.1) | | | | |

**Supplemental Table S4: Baseline characteristics by SARS-CoV-2 PCR test results in Ontario, Quebec, British Columbia, and Manitoba**

| Characteristic | Ontario | | Quebec | | British Columbia | | Manitoba | |
|---|---|---|---|---|---|---|---|---|
| | SARS-CoV-2-positive, n (%)[a] (N=17,437) | SARS-CoV-2-negative, n (%)[a] (N=539,783) | SARS-CoV-2-positive, n (%)[a] (N=7,854) | SARS-CoV-2-negative, n (%)[a] (N=946,354) | SARS-CoV-2-positive, n (%)[a] (N=5,928) | SARS-CoV-2-negative, n (%)[a] (N=870,469) | SARS-CoV-2-positive, n (%)[a] (N=2,201) | SARS-CoV-2-negative, n (%)[a] (N=118,270) |
| COVID-19 hospitalization | 16,726 | - | 7,217 | - | 5,706 | - | 2,127 | - |
| COVID-19-associated death | 3,268 | - | 1,491 | - | 766 | - | 317 | - |
| Received ≥1 dose of COVID-19 vaccine | 2,859 (16.4) | 232,711 (43.1) | 1,040 (13.2) | 439,458 (46.4) | 1,185 (20.0) | 348,062 (40.0) | 346 (15.7) | 52,682 (44.5) |
| Received 2 doses of COVID-19 vaccine | 316 (1.8) | 154,118 (28.6) | 176 (2.2) | 229,820 (24.3) | 252 (4.3) | 205,797 (23.6) | 75 (3.4) | 35,035 (29.6) |
| Age (years), mean (standard deviation) | 62 (18) | 43 (17) | 65 (18) | 47 (17) | 60 (18) | 44 (18) | 54 (20) | 44 (17) |
| Age group (years) | | | | | | | | |
| 18–29 | 804 (4.6) | 140,684 (26.1) | 278 (3.5) | 175,466 (18.5) | 331 (5.6) | 209,917 (24.1) | 344 (15.6) | 30,367 (25.7) |
| 30–39 | 1,401 (8.0) | 127,015 (23.5) | 576 (7.3) | 216,808 (22.9) | 621 (10.5) | 196,562 (22.6) | 293 (13.3) | 28,183 (23.8) |
| 40–49 | 1,966 (11.3) | 90,774 (16.8) | 795 (10.1) | 184,237 (19.5) | 753 (12.7) | 142,650 (16.4) | 296 (13.4) | 20,606 (17.4) |
| 50–59 | 3,217 (18.4) | 77,582 (14.4) | 1,302 (16.6) | 137,422 (14.5) | 1,040 (17.5) | 122,930 (14.1) | 373 (16.9) | 16,262 (13.7) |
| 60–69 | 3,441 (19.7) | 55,067 (10.2) | 1,553 (19.8) | 125,079 (13.2) | 1,233 (20.8) | 98,163 (11.3) | 369 (16.8) | 12,522 (10.6) |
| 70–79 | 3,224 (18.5) | 29,780 (5.5) | 1,605 (20.4) | 72,594 (7.7) | 1,055 (17.8) | 61,185 (7.0) | 301 (13.7) | 6,630 (5.6) |
| ≥80 | 3,384 (19.4) | 18,881 (3.5) | 1,745 (22.2) | 34,748 (3.7) | 895 (15.1) | 39,062 (4.5) | 225 (10.2) | 3,700 (3.1) |
| Male sex | 9,688 (55.6) | 227,350 (42.1) | 4,447 (56.6) | 378,787 (40.0) | 3,413 (57.6) | 391,259 (44.9) | 1,018 (46.3) | 50,762 (42.9) |
| Biweekly period of test | | | | | | | | |
| 14 Dec 2020 to 27 Dec 2020 | 1,044 (6.0) | 26,416 (4.9) | 944 (12.0) | 40,162 (4.2) | 230 (3.9) | 45,742 (5.3) | 146 (6.6) | 5,564 (4.7) |
| 28 Dec 2020 to 10 Jan 2021 | 1,450 (8.3) | 24,642 (4.6) | 1,158 (14.7) | 38,885 (4.1) | 279 (4.7) | 44,287 (5.1) | 129 (5.9) | 4,828 (4.1) |
| 11 Jan 2021 to 24 Jan 2021 | 1,091 (6.3) | 24,367 (4.5) | 839 (10.7) | 41,164 (4.3) | 134 (2.3) | 25,817 (3.0) | 121 (5.5) | 4,966 (4.2) |
| 25 Jan 2021 to 7 Feb 2021 | 781 (4.5) | 21,978 (4.1) | 508 (6.5) | 41,684 (4.4) | 143 (2.4) | 18,705 (2.1) | 75 (3.4) | 4,819 (4.1) |
| 8 Feb 2021 to 21 Feb 2021 | 632 (3.6) | 22,204 (4.1) | 384 (4.9) | 47,729 (5.0) | 272 (4.6) | 46,664 (5.4) | 49 (2.2) | 4,478 (3.8) |
| 22 Feb 2021 to 7 Mar 2021 | 641 (3.7) | 27,363 (5.1) | 346 (4.4) | 49,256 (5.2) | 237 (4.0) | 42,970 (4.9) | 49 (2.2) | 4,825 (4.1) |
| 8 Mar 2021 to 21 Mar 2021 | 966 (5.5) | 29,155 (5.4) | 376 (4.8) | 51,409 (5.4) | 274 (4.6) | 45,938 (5.3) | 52 (2.4) | 4,965 (4.2) |

| | | | | | | | | |
|---|---|---|---|---|---|---|---|---|
| 22 Mar 2021 to 4 Apr 2021 | 1,642 (9.4) | 32,418 (6.0) | 529 (6.7) | 62,676 (6.6) | 303 (5.1) | 43,106 (5.0) | 58 (2.6) | 5,593 (4.7) |
| 5 Apr 2021 to 18 Apr 2021 | 2,662 (15.3) | 39,495 (7.3) | 741 (9.4) | 72,641 (7.7) | 446 (7.5) | 50,770 (5.8) | 80 (3.6) | 7,215 (6.1) |
| 19 Apr 2021 to 2 May 2021 | 2,111 (12.1) | 37,679 (7.0) | 519 (6.6) | 61,383 (6.5) | 621 (10.5) | 62,412 (7.2) | 170 (7.7) | 8,921 (7.5) |
| 3 May 2021 to 16 May 2021 | 1,380 (7.9) | 28,199 (5.2) | 329 (4.2) | 55,154 (5.8) | 539 (9.1) | 53,747 (6.2) | 296 (13.4) | 7,986 (6.8) |
| 17 May 2021 to 30 May 2021 | 766 (4.4) | 17,827 (3.3) | 194 (2.5) | 38,063 (4.0) | 392 (6.6) | 43,436 (5.0) | 306 (13.9) | 7,049 (6.0) |
| 31 May 2021 to 13 Jun 2021 | 441 (2.5) | 14,696 (2.7) | 75 (1.0) | 33,818 (3.6) | 231 (3.9) | 32,209 (3.7) | 217 (9.9) | 5,509 (4.7) |
| 14 Jun 2021 to 27 Jun 2021 | 235 (1.3) | 14,267 (2.6) | 34 (0.4) | 41,365 (4.4) | 122 (2.1) | 26,515 (3.0) | 90 (4.1) | 4,088 (3.5) |
| 28 Jun 2021 to 11 Jul 2021 | 162 (0.9) | 11,944 (2.2) | 23 (0.3) | 36,798 (3.9) | 85 (1.4) | 23,236 (2.7) | 74 (3.4) | 2,962 (2.5) |
| 12 Jul 2021 to 25 Jul 2021 | 92 (0.5) | 15,394 (2.9) | 21 (0.3) | 28,961 (3.1) | 39 (0.7) | 18,251 (2.1) | 49 (2.2) | 3,104 (2.6) |
| 26 Jul 2021 to 8 Aug 2021 | 146 (0.8) | 22,677 (4.2) | 52 (0.7) | 28,091 (3.0) | 43 (0.7) | 19,691 (2.3) | 37 (1.7) | 3,901 (3.3) |
| 9 Aug 2021 to 22 Aug 2021 | 288 (1.7) | 30,093 (5.6) | 116 (1.5) | 30,752 (3.2) | 119 (2.0) | 27,018 (3.1) | 27 (1.2) | 4,985 (4.2) |
| 23 Aug 2021 to 5 Sep 2021 | 406 (2.3) | 31,616 (5.9) | 216 (2.8) | 38,390 (4.1) | 296 (5.0) | 41,510 (4.8) | 41 (1.9) | 6,700 (5.7) |
| 6 Sep 2021 to 19 Sep 2021 | 334 (1.9) | 32,458 (6.0) | 295 (3.8) | 54,638 (5.8) | 384 (6.5) | 53,085 (6.1) | 55 (2.5) | 7,684 (6.5) |
| 20 Sep 2021 to 30 Sep 2021 | 167 (1.0) | 34,895 (6.5) | 155 (2.0) | 53,335 (5.6) | 739 (12.5) | 105,360 (12.1) | 80 (3.6) | 8,128 (6.9) |
| Number of tests in previous 3 months | | | | | | | | |
| 0 | 14,599 (83.7) | 391,672 (72.6) | 6,104 (77.7) | 708,447 (74.9) | 5,106 (86.1) | 735,463 (84.5) | 1,751 (79.6) | 88,031 (74.4) |
| 1 | 1,860 (10.7) | 103,669 (19.2) | 984 (12.5) | 170,316 (18) | 650 (11.0) | 102,182 (11.7) | 311 (14.1) | 24,623 (20.8) |
| ≥2 | 978 (5.6) | 44,442 (8.2) | 766 (9.8) | 67,591 (7.1) | 172 (2.9) | 32,824 (3.8) | 139 (6.3) | 5,616 (4.7) |
| Any comorbidity[b] | 13,343 (76.5) | 248,898 (46.1) | 5,260 (67.0) | 302,647 (32.0) | 3,947 (66.6) | 326,652 (37.5) | 1,431 (65.0) | 45,672 (38.6) |
| Receipt of 2019-2020 and/or 2020-2021 influenza vaccination | 6,783 (38.9) | 178,657 (33.1) | 2,835 (36.1) | 258,090 (27.3) | Not available | Not available | 767 (34.8) | 55,480 (46.9) |
| Neighbourhood income quintile[c] | | | | | | | | |
| 1 (lowest) | 5,678 (32.6) | 95,132 (17.6) | 2,164 (27.6) | 164,636 (17.4) | 1,362 (23.0) | 110,426 (12.7) | 881 (40.0) | 21,057 (17.8) |

| | | | | | | | | |
|---|---|---|---|---|---|---|---|---|
| 2 | 3,824 (21.9) | 104,266 (19.3) | 1,856 (23.6) | 183,802 (19.4) | 1,259 (21.2) | 150,398 (17.3) | 389 (17.7) | 22,919 (19.4) |
| 3 | 3,319 (19.0) | 108,434 (20.1) | 1,553 (19.8) | 195,284 (20.6) | 1,110 (18.7) | 164,168 (18.9) | 356 (16.2) | 22,874 (19.3) |
| 4 | 2,524 (14.5) | 112,380 (20.8) | 1255 (16.0) | 202,334 (21.4) | 928 (15.7) | 186,825 (21.5) | 316 (14.4) | 22,801 (19.3) |
| 5 (highest) | 2,012 (11.5) | 117,116 (21.7) | 1,026 (13.1) | 200,298 (21.2) | 744 (12.6) | 169,532 (19.5) | 193 (8.8) | 23,936 (20.2) |
| Unknown/missing | 80 (0.5) | 2,455 (0.5) | - | - | 525 (8.9) | 89,120 (10.2) | 66 (3.0) | 4,683 (4.0) |
| Essential workers quintile[d] | | | | | | | | |
| 1 (0%–32.5%) | 2,679 (15.4) | 100,570 (18.6) | 1,601 (20.4) | 218,640 (23.1) | 421 (7.1) | 94,738 (10.9) | 598 (27.2) | 24,735 (20.9) |
| 2 (32.5%–42.3%) | 3,313 (19.0) | 122,840 (22.8) | 1,854 (23.6) | 216,807 (22.9) | 867 (14.6) | 160,668 (18.5) | 425 (19.3) | 27,559 (23.3) |
| 3 (42.3%–49.8%) | 3,454 (19.8) | 112,426 (20.8) | 1,661 (21.1) | 193,624 (20.5) | 918 (15.5) | 151,706 (17.4) | 393 (17.9) | 22,714 (19.2) |
| 4 (50.0%–57.5%) | 3,666 (21.0) | 105,236 (19.5) | 1,519 (19.3) | 169,753 (17.9) | 1,136 (19.2) | 136,131 (15.6) | 392 (17.8) | 22,179 (18.8) |
| 5 (57.5%–100%) | 4,226 (24.2) | 94,953 (17.6) | 1,219 (15.5) | 147,530 (15.6) | 1,300 (21.9) | 123,183 (14.2) | 354 (16.1) | 19,244 (16.3) |
| Unknown/missing | 99 (0.6) | 3,758 (0.7) | - | - | 1,286 (21.7) | 204,043 (23.4) | 28 (1.3) | 357 (0.3) |
| Persons per dwelling quintile[e] | | | | | | | | |
| 1 (0–2.1) | 3,410 (19.6) | 98,120 (18.2) | 1,745 (22.2) | 190,315 (20.1) | 1,368 (23.1) | 179,935 (20.7) | 625 (28.4) | 29,676 (25.1) |
| 2 (2.2–2.4) | 2,619 (15.0) | 97,786 (18.1) | 1,086 (13.8) | 142,283 (15.0) | 941 (15.9) | 146,373 (16.8) | 279 (12.7) | 19,304 (16.3) |
| 3 (2.5–2.6) | 1,945 (11.2) | 69,988 (13.0) | 1,337 (17.0) | 164,333 (17.4) | 956 (16.1) | 151,365 (17.4) | 264 (12.0) | 19,820 (16.8) |
| 4 (2.7–3.0) | 4,107 (23.6) | 128,988 (23.9) | 1,833 (23.3) | 238,048 (25.2) | 902 (15.2) | 157,131 (18.1) | 386 (17.5) | 25,032 (21.2) |
| 5 (3.1–5.7) | 5,249 (30.1) | 140,991 (26.1) | 1,853 (23.6) | 211,375 (22.3) | 1,605 (27.1) | 184,599 (21.2) | 608 (27.6) | 22,599 (19.1) |
| Unknown/missing | 107 (0.6) | 3,910 (0.7) | - | - | 156 (2.6) | 51,066 (5.9) | 28 (1.3) | 357 (0.3) |
| Self-identified visible minority quintile[f] | | | | | | | | |
| 1 (0.0%–2.2%) | 1,316 (7.5) | 91,833 (17.0) | 1,200 (15.3) | 222,658 (23.5) | 853 (14.4) | 99,619 (11.4) | 456 (20.7) | 17,833 (15.1) |
| 2 (2.2%–7.5%) | 1,651 (9.5) | 100,772 (18.7) | 783 (10.0) | 147,473 (15.6) | 962 (16.2) | 146,322 (16.8) | 459 (20.9) | 22,735 (19.2) |
| 3 (7.5%–18.7%) | 2,512 (14.4) | 99,293 (18.4) | 1,222 (15.6) | 217,522 (23.0) | 1,050 (17.7) | 177,798 (20.4) | 274 (12.4) | 24,072 (20.4) |
| 4 (18.7%–43.5%) | 4,225 (24.2) | 110,556 (20.5) | 1,664 (21.2) | 197,633 (20.9) | 1,205 (20.3) | 203,173 (23.3) | 375 (17.0) | 25,918 (21.9) |
| 5 (43.5%–100%) | 7,634 (43.8) | 133,580 (24.7) | 2,985 (38.0) | 161,068 (17.0) | 1,705 (28.8) | 193,488 (22.2) | 598 (27.2) | 25,873 (21.9) |
| Unknown/missing | 99 (0.6) | 3,749 (0.7) | - | - | 153 (2.6) | 50,069 (5.8) | 28 (1.3) | 357 (0.3) |

[a]Proportion reported, unless stated otherwise.

[b]Comorbidities include chronic respiratory diseases, chronic heart diseases, hypertension, diabetes, immunocompromising conditions due to underlying diseases or therapy, autoimmune diseases, chronic kidney disease, advanced liver disease, dementia/frailty and history of stroke or transient ischemic attack.

[c]Neighbourhood income quintile has variable cut-off values in each city/Census area to account for cost of living. A dissemination area (DA) being in quintile 1 means it is among the lowest 20% of DAs in its city by income. Material deprivation index quintile used in British Columbia; quintile 1 represents 'most deprived' and quintile 5 represents 'least deprived'.

[d]Percentage of people in the area working in the following occupations: sales and service occupations; trades, transport and equipment operators and related occupations; natural resources, agriculture, and related production occupations; and occupations in manufacturing and utilities. Census counts for people are randomly rounded up or down to the nearest number divisible by 5, which causes some minor imprecision.

[e]Range of persons per dwelling.

[f]Percentage of people in the area who self-identified as a visible minority. Census counts for people are randomly rounded up or down to the nearest number divisible by 5, which causes some minor imprecision.

**Supplemental Table S5: Baseline characteristics by vaccination status in Ontario, Quebec, British Columbia, and Manitoba**

| Characteristic | Ontario | | Quebec | | British Columbia | | Manitoba | |
|---|---|---|---|---|---|---|---|---|
| | Received ≥1 dose, n (%)[a] (N=235,570) | Unvaccinated, n (%)[a] (N=321,650) | Received ≥1 dose, n (%)[a] (N=440,498) | Unvaccinated, n (%)[a] (N=513,710) | Received ≥1 dose, n (%)[a] (N=349,247) | Unvaccinated, n (%)[a] (N=527,150) | Received ≥1 dose, n (%)[a] (N=53,028) | Unvaccinated, n (%)[a] (N=67,443) |
| Age (years), mean (standard deviation) | 45 (19) | 43 (17) | 49 (18) | 45 (16) | 48 (19) | 43 (17) | 46 (18) | 42 (16) |
| Age group (years) | | | | | | | | |
| 18–29 | 54,047 (22.9) | 87,441 (27.2) | 7,6075 (17.3) | 99,669 (19.4) | 67,740 (19.4) | 142,508 (27.0) | 12,200 (23.0) | 18,511 (27.4) |
| 30–39 | 53,454 (22.7) | 74,962 (23.3) | 89,211 (20.3) | 128,173 (25) | 70,671 (20.2) | 126,512 (24.0) | 11,692 (22.0) | 16,784 (24.9) |
| 40–49 | 37,775 (16.0) | 54,965 (17.1) | 77,354 (17.6) | 107,678 (21.0) | 55,917 (16.0) | 87,486 (16.6) | 9,080 (17.1) | 11,822 (17.5) |
| 50–59 | 33,292 (14.1) | 47,507 (14.8) | 62,394 (14.2) | 76,330 (14.9) | 50,322 (14.4) | 73,648 (14.0) | 7,601 (14.3) | 9,034 (13.4) |
| 60–69 | 28,015 (11.9) | 30,493 (9.5) | 66,572 (15.1) | 60,060 (11.7) | 46,080 (13.2) | 53,316 (10.1) | 6,620 (12.5) | 6,271 (9.3) |
| 70–79 | 16,818 (7.1) | 16,186 (5.0) | 45,145 (10.2) | 29,054 (5.7) | 339,31 (9.7) | 28,309 (5.4) | 3,688 (7.0) | 3,243 (4.8) |
| ≥80 | 12,169 (5.2) | 10,096 (3.1) | 23,747 (5.4) | 12,746 (2.5) | 24,586 (7.0) | 15,371 (2.9) | 2,147 (4.0) | 1,778 (2.6) |
| Male sex | 93,281 (39.6) | 143,757 (44.7) | 170,763 (38.8) | 212,471 (41.4) | 145,873 (41.8) | 248,799 (47.2) | 21,685 (40.9) | 30,095 (44.6) |
| Biweekly period of test | | | | | | | | |
| 14 Dec 2020 to 27 Dec 2020 | 13 (0.0) | 27,447 (8.5) | 105 (0.0) | 41,001 (8.0) | 276 (0.1) | 45,696 (8.7) | 11 (0.0) | 5,699 (8.5) |
| 28 Dec 2020 to 10 Jan 2021 | 266 (0.1) | 25,826 (8.0) | 587 (0.1) | 39,456 (7.7) | 1,204 (0.3) | 43,362 (8.2) | 19 (0.0) | 4,938 (7.3) |
| 11 Jan 2021 to 24 Jan 2021 | 894 (0.4) | 24,564 (7.6) | 1,747 (0.4) | 40,256 (7.8) | 1,522 (0.4) | 24,429 (4.6) | 78 (0.1) | 5,009 (7.4) |
| 25 Jan 2021 to 7 Feb 2021 | 1,077 (0.5) | 21,682 (6.7) | 2,190 (0.5) | 40,002 (7.8) | 1,392 (0.4) | 17,456 (3.3) | 133 (0.3) | 4,761 (7.1) |
| 8 Feb 2021 to 21 Feb 2021 | 975 (0.4) | 21,861 (6.8) | 2,666 (0.6) | 45,447 (8.8) | 3,366 (1.0) | 43,570 (8.3) | 132 (0.2) | 4,395 (6.5) |
| 22 Feb 2021 to 7 Mar 2021 | 1,388 (0.6) | 26,616 (8.3) | 3,330 (0.8) | 46,272 (9) | 2,673 (0.8) | 40,534 (7.7) | 175 (0.3) | 4,699 (7.0) |
| 8 Mar 2021 to 21 Mar 2021 | 2,532 (1.1) | 27,589 (8.6) | 4,863 (1.1) | 46,922 (9.1) | 3,389 (1.0) | 428,23 (8.1) | 316 (0.6) | 4,701 (7.0) |
| 22 Mar 2021 to 4 Apr 2021 | 4,683 (2.0) | 29,377 (9.1) | 8,460 (1.9) | 54,745 (10.7) | 4,771 (1.4) | 38,638 (7.3) | 533 (1.0) | 5,118 (7.6) |
| 5 Apr 2021 to 18 Apr 2021 | 8,713 (3.7) | 33,444 (10.4) | 14,720 (3.3) | 58,662 (11.4) | 7,355 (2.1%) | 43,861 (8.3%) | 1,152 (2.2) | 6,143 (9.1) |
| 19 Apr 2021 to 2 May 2021 | 11,941 (5.1) | 27,849 (8.7) | 21,751 (4.9) | 40,151 (7.8) | 12,952 (3.7) | 50,081 (9.5) | 2,474 (4.7) | 6,617 (9.8) |
| 3 May 2021 to 16 May 2021 | 13,234 (5.6) | 16,345 (5.1) | 27,484 (6.2) | 27,999 (5.5) | 17,208 (4.9) | 37,078 (7.0) | 3,290 (6.2) | 4,992 (7.4) |
| 17 May 2021 to 30 May 2021 | 11,422 (4.8) | 7,171 (2.2) | 26,631 (6.0) | 11,626 (2.3) | 18,934 (5.4) | 24,894 (4.7) | 4,490 (8.5) | 2,865 (4.2) |
| 31 May 2021 to 13 Jun 2021 | 11,059 (4.7) | 4,078 (1.3) | 29,057 (6.6) | 4,836 (0.9) | 19,715 (5.6) | 12,725 (2.4) | 3,821 (7.2) | 1,905 (2.8) |
| 14 Jun 2021 to 27 Jun 2021 | 11,426 (4.9) | 3,076 (1.0) | 37,785 (8.6) | 3,614 (0.7) | 19,311 (5.5) | 7,326 (1.4) | 3,081 (5.8) | 1,097 (1.6) |
| 28 Jun 2021 to 11 Jul 2021 | 9,814 (4.2) | 2,292 (0.7) | 33,942 (7.7) | 2,879 (0.6) | 17,325 (5.0) | 5,996 (1.1) | 2,365 (4.5) | 671 (1.0) |
| 12 Jul 2021 to 25 Jul 2021 | 12,741 (5.4) | 2,745 (0.9) | 26,855 (6.1) | 21,27 (0.4) | 1,3687 (3.9) | 4,603 (0.9) | 2,574 (4.9) | 579 (0.9) |
| 26 Jul 2021 to 8 Aug 2021 | 19,382 (8.2) | 3,441 (1.1) | 26,235 (6) | 1,908 (0.4) | 15,247 (4.4) | 4,487 (0.9) | 3,355 (6.3) | 583 (0.9) |
| 9 Aug 2021 to 22 Aug 2021 | 26,121 (11.1) | 4,260 (1.3) | 29,093 (6.6) | 1,775 (0.3) | 21,378 (6.1) | 5,759 (1.1) | 4,426 (8.3) | 586 (0.9) |
| 23 Aug 2021 to 5 Sep 2021 | 27,686 (11.8) | 4,336 (1.3) | 37,009 (8.4) | 1,597 (0.3) | 33,339 (9.5) | 8,467 (1.6) | 6,035 (11.4) | 706 (1.0) |
| 6 Sep 2021 to 19 Sep 2021 | 28,869 (12.3) | 3,923 (1.2) | 53,400 (12.1) | 1,533 (0.3) | 43,886 (12.6) | 9,583 (1.8) | 7,035 (13.3) | 704 (1.0) |
| 20 Sep 2021 to 30 Sep 2021 | 31,334 (13.3) | 3,728 (1.2) | 52,588 (11.9) | 902 (0.2) | 90,317 (25.9) | 15,782 (3.0) | 7,533 (14.2) | 675 (1.0) |
| Number of tests in previous 3 months | | | | | | | | |

| | | | | | | | | |
|---|---|---|---|---|---|---|---|---|
| 0 | 168,061 (71.3) | 238,210 (74.1) | 327,233 (74.3) | 387,318 (75.4) | 271,652 (77.8) | 468,917 (89.0) | 39,569 (74.6) | 50,213 (74.5) |
| 1 | 45,235 (19.2) | 60,294 (18.7) | 7,6736 (17.4) | 94,564 (18.4) | 55,491 (15.9) | 47,341 (9.0) | 11,047 (20.8) | 13,887 (20.6) |
| ≥2 | 22,274 (9.5) | 23,146 (7.2) | 36,529 (8.3) | 31,828 (6.2) | 22,104 (6.3) | 10,892 (2.1) | 2,412 (4.5) | 3,343 (5.0) |
| Any comorbidity[b] | 114,748 (48.7) | 147,493 (45.9) | 158,295 (35.9) | 149,612 (29.1) | 151,206 (43.3) | 179,393 (34.0) | 22,095 (41.7) | 25,008 (37.1) |
| Receipt of 2019-2020 and/or 2020-2021 influenza vaccination | 93,962 (39.9) | 91,478 (28.4) | 147,342 (33.4) | 113,583 (22.1) | Not available | Not available | 29,224 (55.1) | 27,023 (40.1) |
| Neighbourhood income quintile[c] | | | | | | | | |
| 1 (lowest) | 39,233 (16.7) | 61,577 (19.1) | 7,3547 (16.7) | 93,253 (18.2) | 42,888 (12.3) | 68,900 (13.1) | 8,832 (16.7) | 13,106 (19.4) |
| 2 | 44,969 (19.1) | 63,121 (19.6) | 84,055 (19.1) | 101,603 (19.8) | 60,126 (17.2) | 91,531 (17.4) | 9,918 (18.7) | 13,390 (19.9) |
| 3 | 48,229 (20.5) | 63,524 (19.7) | 89,999 (20.4) | 106,838 (20.8) | 66,874 (19.1) | 98,404 (18.7) | 10,393 (19.6) | 12,837 (19.0) |
| 4 | 49,665 (21.1) | 65,239 (20.3) | 94,224 (21.4) | 109,365 (21.3) | 78,876 (22.6) | 108,877 (20.7) | 10,341 (19.5) | 12,776 (18.9) |
| 5 (highest) | 52,536 (22.3) | 66,592 (20.7) | 98,673 (22.4) | 102,651 (20) | 70,720 (20.2) | 99,556 (18.9) | 11,218 (21.2) | 12,911 (19.1) |
| Unknown/missing | 938 (0.4) | 1,597 (0.5) | - | - | 29,763 (8.5) | 59,882 (11.4) | 2,326 (4.4) | 2,423 (3.6) |
| Essential workers quintile[d] | | | | | | | | |
| 1 (0%–32.5%) | 46,555 (19.8) | 56,694 (17.6) | 108,031 (24.5) | 112,210 (21.8) | 39,889 (11.4) | 55,270 (10.5) | 11,860 (22.4) | 13,473 (20.0) |
| 2 (32.5%–42.3%) | 56,274 (23.9) | 69,879 (21.7) | 102,009 (23.2) | 116,652 (22.7) | 67,543 (19.3) | 93,992 (17.8) | 12,581 (23.7) | 15,403 (22.8) |
| 3 (42.3%–49.8%) | 49,376 (21.0) | 66,504 (20.7) | 88,769 (20.2) | 106,516 (20.7) | 63,453 (18.2) | 89,171 (16.9) | 9,984 (18.8) | 13,123 (19.5) |
| 4 (50.0%–57.5%) | 44,485 (18.9) | 64,417 (20.0) | 7,6257 (17.3) | 95,015 (18.5) | 54,985 (15.7) | 82,282 (15.6) | 9,648 (18.2) | 12,923 (19.2) |
| 5 (57.5%–100%) | 37,474 (15.9) | 61,705 (19.2) | 65,432 (14.9) | 83,317 (16.2) | 48,061 (13.8) | 76,422 (14.5) | 8,088 (15.3) | 11,510 (17.1) |
| Unknown/missing | 1,406 (0.6) | 2,451 (0.8) | - | - | 75,316 (21.6) | 130,013 (24.7) | 150 (0.3) | 235 (0.3) |
| Persons per dwelling quintile[e] | | | | | | | | |
| 1 (0–2.1) | 40,938 (17.4) | 60,592 (18.8) | 87,054 (19.8) | 105,006 (20.4) | 75,494 (21.6) | 105,809 (20.1) | 13,178 (24.9) | 17,123 (25.4) |
| 2 (2.2–2.4) | 39,935 (17.0) | 60,470 (18.8) | 66,161 (15.0) | 77,208 (15.0) | 59,910 (17.2) | 87,404 (16.6) | 8,551 (16.1) | 11,032 (16.4) |
| 3 (2.5–2.6) | 29,449 (12.5) | 42,484 (13.2) | 76,064 (17.3) | 89,606 (17.4) | 62,129 (17.8) | 90,192 (17.1) | 8,785 (16.6) | 11,299 (16.8) |
| 4 (2.7–3.0) | 57,106 (24.2) | 75,989 (23.6) | 109,072 (24.8) | 130,809 (25.5) | 64,785 (18.6) | 93,248 (17.7) | 11,226 (21.2) | 14,192 (21.0) |
| 5 (3.1–5.7) | 66,656 (28.3) | 79,584 (24.7) | 102,147 (23.2) | 111,081 (21.6) | 74,026 (21.2) | 112,178 (21.3) | 10,421 (19.7) | 12,786 (19.0) |
| Unknown/missing | 1,486 (0.6) | 2,531 (0.8) | - | - | 12,903 (3.7) | 38,319 (7.3) | 150 (0.3) | 235 (0.3) |
| Self-identified visible minority quintile[f] | | | | | | | | |
| 1 (0.0%–2.2%) | 36,144 (15.3) | 57,005 (17.7) | 102,373 (23.2) | 121,485 (23.6) | 42,165 (12.1) | 58,307 (11.1) | 8,028 (15.1) | 10,261 (15.2) |
| 2 (2.2%–7.5%) | 40,698 (17.3) | 61,725 (19.2) | 67,066 (15.2) | 81,190 (15.8) | 61,771 (17.7) | 85,513 (16.2) | 10,158 (19.2) | 13,036 (19.3) |
| 3 (7.5%–18.7%) | 43,069 (18.3) | 58,736 (18.3) | 99,999 (22.7) | 118,745 (23.1) | 73,099 (20.9) | 105,749 (20.1) | 10,946 (20.6) | 13,400 (19.9) |
| 4 (18.7%–43.5%) | 50,189 (21.3) | 64,592 (20.1) | 94,020 (21.3) | 105,277 (20.5) | 83,457 (23.9) | 120,921 (22.9) | 11,547 (21.8) | 14,746 (21.9) |
| 5 (43.5%–100%) | 64,066 (27.2) | 77,148 (24.0) | 77,040 (17.5) | 87,013 (16.9) | 76,264 (21.8) | 118,929 (22.6) | 11,482 (21.7) | 14,989 (22.2) |
| Unknown/missing | 1,404 (0.6) | 2,444 (0.8) | - | - | 12,491 (3.6) | 37,731 (7.2) | 150 (0.3) | 235 (0.3) |

[a]Proportion reported, unless stated otherwise.

[b]Comorbidities include chronic respiratory diseases, chronic heart diseases, hypertension, diabetes, immunocompromising conditions due to underlying diseases or therapy, autoimmune diseases, chronic kidney disease, advanced liver disease, dementia/frailty and history of stroke or transient ischemic attack.

[c]Neighbourhood income quintile has variable cut-off values in each city/Census area to account for cost of living. A dissemination area (DA) being in quintile 1 means it is among the lowest 20% of DAs in its city by income. Material deprivation index quintile used in British Columbia; quintile 1 represents 'most deprived' and quintile 5 represents 'least deprived'.
[d]Percentage of people in the area working in the following occupations: sales and service occupations; trades, transport and equipment operators and related occupations; natural resources, agriculture, and related production occupations; and occupations in manufacturing and utilities. Census counts for people are randomly rounded up or down to the nearest number divisible by 5, which causes some minor imprecision.
[e]Range of persons per dwelling.
[f]Percentage of people in the area who self-identified as a visible minority. Census counts for people are randomly rounded up or down to the nearest number divisible by 5, which causes some minor imprecision.

**Supplemental Table S6: Baseline characteristics by vaccine product in Ontario, Quebec, British Columbia, and Manitoba**

| Characteristic | Ontario | | | Quebec | | | British Columbia | | | Manitoba | | |
|---|---|---|---|---|---|---|---|---|---|---|---|---|
| | Received ≥1 dose BNT162b2, n (%)[a] (N=151,086) | Received ≥1 dose mRNA-1273, n (%)[a] (N=42,994) | Received ≥1 dose AstraZeneca/COVISHIELD, n (%)[a] (N=10,940) | Received ≥1 dose BNT162b2, n (%)[a] (N=316,535) | Received ≥1 dose mRNA-1273, n (%)[a] (N=86,177) | Received ≥1 dose AstraZeneca/COVISHIELD, n (%)[a] (N=37,786) | Received ≥1 dose BNT162b2, n (%)[a] (N=240,043) | Received ≥1 dose mRNA-1273, n (%)[a] (N=64,367) | Received ≥1 dose AstraZeneca/COVISHIELD, n (%)[a] (N=17,748) | Received ≥1 dose BNT162b2, n (%)[a] (N=34,622) | Received ≥1 dose mRNA-1273, n (%)[a] (N=12,480) | Received ≥1 dose AstraZeneca/COVISHIELD, n (%)[a] (N=5,916) |
| Age (years), mean (standard deviation) | 46 (19) | 44 (19) | 57 (9) | 48 (19) | 46 (18) | 60 (11) | 48 (20) | 46 (19) | 51 (11) | 45 (19) | 43 (19) | 51 (10) |
| Age group (years) | | | | | | | | | | | | |
| 18–29 | 35,733 (23.7) | 11,078 (25.8) | 16 (0.1) | 57,655 (18.2) | 18,187 (21.1) | 233 (0.6) | 49,819 (20.8) | 14,667 (22.8) | 729 (4.1) | 8,545 (24.7) | 3,594 (28.8) | 59 (1.0) |
| 30–39 | 35,444 (23.5) | 10,651 (24.8) | 278 (2.5) | 68,649 (21.7) | 20,213 (23.5) | 349 (0.9) | 49,858 (20.8) | 14,246 (22.1) | 2,201 (12.4) | 8,165 (23.6) | 3,086 (24.7) | 439 (7.4) |
| 40–49 | 22,333 (14.8) | 6,368 (14.8) | 2,259 (20.6) | 57,415 (18.1) | 14,344 (16.6) | 5,595 (14.8) | 36,158 (15.1) | 9,552 (14.8) | 4,525 (25.5) | 4,665 (13.5) | 1,678 (13.4) | 2,735 (46.2) |
| 50–59 | 19,690 (13.0) | 5,641 (13.1) | 3,179 (29.1) | 34,003 (10.7) | 10,209 (11.8) | 18,182 (48.1) | 30,341 (12.6) | 8,442 (13.1) | 5,954 (33.5) | 4,438 (12.8) | 1,533 (12.3) | 1,629 (27.5) |
| 60–69 | 16,006 (10.6) | 4,047 (9.4) | 4,661 (42.6) | 48,068 (15.2) | 12,306 (14.3) | 6,198 (16.4) | 28,555 (11.9) | 7,974 (12.4) | 4,128 (23.3) | 4,544 (13.1) | 1,231 (9.9) | 842 (14.2) |
| 70–79 | 12,703 (8.4) | 2,602 (6.1) | 500 (4.6) | 32,626 (10.3) | 6,782 (7.9) | 5,737 (15.2) | 26,078 (10.9) | 5,345 (8.3) | 174 (1.0) | 2,776 (8.0) | 779 (6.2) | 133 (2.2) |
| ≥80 | 9,177 (6.1) | 2,607 (6.1) | 47 (0.4) | 18,119 (5.7) | 4,136 (4.8) | 1,492 (3.9) | 19,234 (8.0) | 4,141 (6.4) | 37 (0.2) | 1,489 (4.3) | 579 (4.6) | 79 (1.3) |
| Male sex | 56,434 (37.4) | 17,833 (41.5) | 5,305 (48.5) | 119,121 (37.6) | 33,519 (38.9) | 18,123 (48.0) | 95,935 (40.0) | 28,319 (44.0) | 9,147 (51.5) | 13,437 (38.8) | 5,520 (44.2) | 2,722 (46.0) |
| Biweekly period of test | | | | | | | | | | | | |
| 14 Dec 2020 to 27 Dec 2020 | 13 (0.0) | 0 (0.0) | 0 (0.0) | 105 (0.0) | 0 (0.0) | 0 (0.0) | 276 (0.1) | 0 (0.0) | 0 (0.0) | 11 (0.0) | ≤6 (≤0.0) | ≤6 (≤0.0) |
| 28 Dec 2020 to 10 Jan 2021 | 253 (0.2) | 13 (0.0) | 0 (0.0) | 577 (0.2) | 10 (0.0) | 0 (0.0) | 1,127 (0.5) | 77 (0.1) | 0 (0.0) | 19 (0.1) | ≤6 (≤0.0) | ≤6 (≤0.0) |
| 11 Jan 2021 to 24 Jan 2021 | 765 (0.5) | 129 (0.3) | 0 (0.0) | 1,648 (0.5) | 99 (0.1) | 0 (0.0) | 1,332 (0.6) | 190 (0.3) | 0 (0.0) | 65 (0.2) | 13 (0.1) | ≤6 (≤0.0) |
| 25 Jan 2021 to 7 Feb 2021 | 916 (0.6) | 161 (0.4) | 0 (0.0) | 2,057 (0.6) | 133 (0.2) | 0 (0.0) | 1,190 (0.5) | 202 (0.3) | 0 (0.0) | 96 (0.3) | 37 (0.3) | ≤6 (≤0.0) |

| | | | | | | | | | | | | |
|---|---|---|---|---|---|---|---|---|---|---|---|---|
| 8 Feb 2021 to 21 Feb 2021 | 766 (0.5) | 209 (0.5) | 0 (0.0) | 2,378 (0.8) | 288 (0.3) | 0 (0.0) | 2,818 (1.2) | 547 (0.8) | 1 (0.0) | 74 (0.2) | 58 (0.5) | ≤6 (≤0.0) |
| 22 Feb 2021 to 7 Mar 2021 | 1,088 (0.7) | 300 (0.7) | 0 (0.0) | 2,949 (0.9) | 381 (0.4) | 0 (0.0) | 2,124 (0.9) | 547 (0.8) | 0 (0.0) | 104 (0.3) | 71 (0.6) | ≤6 (≤0.0) |
| 8 Mar 2021 to 21 Mar 2021 | 2,107 (1.4) | 361 (0.8) | 64 (0.6) | 4,028 (1.3) | 541 (0.6) | 294 (0.8) | 2,821 (1.2) | 567 (0.9) | 0 (0.0) | 228 (0.7) | 80 (0.6) | 8 (0.1) |
| 22 Mar 2021 to 4 Apr 2021 | 3,601 (2.4) | 669 (1.6) | 413 (3.8) | 6,552 (2.1) | 1,125 (1.3) | 783 (2.1) | 4,191 (1.7) | 560 (0.9) | 18 (0.1) | 327 (0.9) | 122 (1.0) | 84 (1.4) |
| 5 Apr 2021 to 18 Apr 2021 | 6,532 (4.3) | 1,298 (3.0) | 882 (8.1) | 11,084 (3.5) | 2,021 (2.3) | 1,615 (4.3) | 5,726 (2.4) | 1,209 (1.9) | 416 (2.3) | 755 (2.2) | 249 (2.0) | 148 (2.5) |
| 19 Apr 2021 to 2 May 2021 | 8,122 (5.4) | 2,090 (4.9) | 1,729 (15.8) | 14,240 (4.5) | 3,919 (4.5) | 3,592 (9.5) | 9,602 (4.0) | 1,999 (3.1) | 1,349 (7.6) | 1,357 (3.9) | 581 (4.7) | 536 (9.1) |
| 3 May 2021 to 16 May 2021 | 9,075 (6.0) | 2,371 (5.5) | 1,787 (16.3) | 18,118 (5.7) | 5,421 (6.3) | 3,945 (10.4) | 11,731 (4.9) | 2,867 (4.5) | 2,607 (14.7) | 1,787 (5.2) | 861 (6.9) | 642 (10.9) |
| 17 May 2021 to 30 May 2021 | 8,148 (5.4) | 2,170 (5.0) | 1,104 (10.1) | 18,764 (5.9) | 5,167 (6.0) | 2,700 (7.1) | 12,649 (5.3) | 3,159 (4.9) | 3,120 (17.6) | 2,743 (7.9) | 1,173 (9.4) | 574 (9.7) |
| 31 May 2021 to 13 Jun 2021 | 7,965 (5.3) | 2,274 (5.3) | 812 (7.4) | 20,841 (6.6) | 5,918 (6.9) | 2,298 (6.1) | 14,167 (5.9) | 3,406 (5.3) | 2,123 (12.0) | 2,389 (6.9) | 1,022 (8.2) | 409 (6.9) |
| 14 Jun 2021 to 27 Jun 2021 | 8,142 (5.4) | 2,204 (5.1) | 781 (7.1) | 27,305 (8.6) | 7,533 (8.7) | 2,947 (7.8) | 14,118 (5.9) | 3,371 (5.2) | 1,651 (9.3) | 2,020 (5.8) | 750 (6.0) | 311 (5.3) |
| 28 Jun 2021 to 11 Jul 2021 | 6,546 (4.3) | 1,849 (4.3) | 323 (3.0) | 24,582 (7.8) | 6,772 (7.9) | 2,588 (6.8) | 12,633 (5.3) | 2,858 (4.4) | 1,272 (7.2) | 1,541 (4.5) | 627 (5.0) | 197 (3.3) |
| 12 Jul 2021 to 25 Jul 2021 | 7,698 (5.1) | 2,391 (5.6) | 334 (3.1) | 19,374 (6.1) | 5,431 (6.3) | 2,050 (5.4) | 9,402 (3.9) | 2,342 (3.6) | 523 (2.9) | 1,678 (4.8) | 683 (5.5) | 212 (3.6) |
| 26 Jul 2021 to 8 Aug 2021 | 11,460 (7.6) | 3,549 (8.3) | 416 (3.8) | 18,722 (5.9) | 5,372 (6.2) | 2,141 (5.7) | 10,302 (4.3) | 2,761 (4.3) | 391 (2.2) | 2,273 (6.6) | 801 (6.4) | 279 (4.7) |
| 9 Aug 2021 to 22 Aug 2021 | 15,455 (10.2) | 4,783 (11.1) | 566 (5.2) | 20,743 (6.6) | 6,008 (7.0) | 2,342 (6.2) | 14,397 (6.0) | 3,962 (6.2) | 504 (2.8) | 3,059 (8.8) | 948 (7.6) | 417 (7.0) |
| 23 Aug 2021 to 5 Sep 2021 | 16,507 (10.9) | 4,954 (11.5) | 574 (5.2) | 26,475 (8.4) | 7,642 (8.9) | 2,892 (7.7) | 21,949 (9.1) | 6,653 (10.3) | 767 (4.3) | 4,139 (12.0) | 1,322 (10.6) | 573 (9.7) |
| 6 Sep 2021 to 19 Sep 2021 | 17,119 (11.3) | 5,470 (12.7) | 589 (5.4) | 38,270 (12.1) | 11,257 (13.1) | 3,873 (10.2) | 28,633 (11.9) | 8,824 (13.7) | 991 (5.6) | 4,813 (13.9) | 1,470 (11.8) | 751 (12.7) |
| 20 Sep 2021 to 30 Sep 2021 | 18,808 (12.4) | 5,749 (13.4) | 566 (5.2) | 37,723 (11.9) | 11,139 (12.9) | 3,726 (9.9) | 58,855 (24.5) | 18,266 (28.4) | 2,015 (11.4) | 5,144 (14.9) | 1,612 (12.9) | 775 (13.1) |
| Number of tests in previous 3 months | | | | | | | | | | | | |
| 0 | 105,902 (70.1) | 30,594 (71.2) | 8,534 (78.0) | 231,000 (73.0) | 65,585 (76.1) | 30,648 (81.1) | 184,903 (77.0) | 51,341 (79.8) | 14,330 (80.7) | 25,980 (75.0) | 9,286 (74.4) | 4,294 (72.6) |
| 1 | 28,878 (19.1) | 8,414 (19.6) | 1,931 (17.7) | 55,916 (17.7) | 15,274 (17.7) | 5,546 (14.7) | 38,667 (16.1) | 9,664 (15.0) | 2,696 (15.2) | 7,113 (20.5) | 2,594 (20.8) | 1,339 (22.6) |

| | | | | | | | | | | | | |
|---|---|---|---|---|---|---|---|---|---|---|---|---|
| ≥2 | 16,306 (10.8) | 3,986 (9.3) | 475 (4.3) | 29,619 (9.4) | 5,318 (6.2) | 1,592 (4.2) | 16,473 (6.9) | 3,362 (5.2) | 722 (4.1) | 1,529 (4.4) | 600 (4.8) | 283 (4.8) |
| Any comorbidity[b] | 74,856 (49.5) | 20,733 (48.2) | 6,420 (58.7) | 111,580 (35.3) | 29,409 (34.1) | 17,306 (45.8) | 105,466 (43.9) | 26,286 (40.8) | 7,275 (41.0) | 13,844 (40.0) | 5,250 (42.1) | 2,999 (50.7) |
| Receipt of 2019-2020 and/or 2020-2021 influenza vaccination | 60,439 (40.0) | 14,658 (34.1) | 6,239 (57.0) | 108,470 (34.3) | 24,762 (28.7) | 14,110 (37.3) | N/A | N/A | N/A | 19,685 (56.9) | 5,619 (45.0) | 3,917 (66.2) |
| Neighbourhood income quintile[c] | | | | | | | | | | | | |
| 1 (lowest) | 24,925 (16.5) | 8,496 (19.8) | 1,513 (13.8) | 52,306 (16.5) | 15,323 (17.8) | 5,918 (15.7) | 28,543 (11.9) | 9,634 (15.0) | 4,294 (24.2) | 4,630 (13.4) | 3,547 (28.4) | 655 (11.1) |
| 2 | 29,058 (19.2) | 8,344 (19.4) | 1,912 (17.5) | 60,760 (19.2) | 16,358 (19.0) | 6,937 (18.4) | 40,477 (16.9) | 12,518 (19.4) | 4,220 (23.8) | 6,070 (17.5) | 2,815 (22.6) | 1,032 (17.4) |
| 3 | 31,201 (20.7) | 8,831 (20.5) | 2,050 (18.7) | 65,083 (20.6) | 17,081 (19.8) | 7,835 (20.7) | 45,915 (19.1) | 12,257 (19.0) | 3,288 (18.5) | 7,212 (20.8) | 2,018 (16.2) | 1,163 (19.7) |
| 4 | 32,293 (21.4) | 8,542 (19.9) | 2,226 (20.3) | 68,137 (21.5) | 17,913 (20.8) | 8,174 (21.6) | 55,061 (22.9) | 12,825 (19.9) | 2,709 (15.3) | 7,089 (20.5) | 1,996 (16.0) | 1,252 (21.2) |
| 5 (highest) | 33,001 (21.8) | 8,581 (20.0) | 3,213 (29.4) | 70,249 (22.2) | 19,502 (22.6) | 8,922 (23.6) | 49,446 (20.6) | 10,963 (17.0) | 2,014 (11.3) | 7,908 (22.8) | 1,746 (14.0) | 1,561 (26.4) |
| Unknown/missing | 608 (0.4) | 200 (0.5) | 26 (0.2) | - | - | - | 20,601 (8.6) | 6,170 (9.6) | 1,223 (6.9) | 1,713 (4.9) | 358 (2.9) | 253 (4.3) |
| Essential workers quintile[d] | | | | | | | | | | | | |
| 1 (0%–32.5%) | 29,176 (19.3) | 6,975 (16.2) | 2,981 (27.2) | 79,678 (25.2) | 18,980 (22.0) | 9,373 (24.8) | 28,100 (11.7) | 6,350 (9.9) | 2,291 (12.9) | 6,124 (17.7) | 4,519 (36.2) | 1,215 (20.5) |
| 2 (32.5%–42.3%) | 36,412 (24.1) | 9,741 (22.7) | 2,577 (23.6) | 73,680 (23.3) | 19,649 (22.8) | 8,680 (23.0) | 47,340 (19.7) | 10,689 (16.6) | 3,744 (21.1) | 8,720 (25.2) | 2,249 (18.0) | 1,608 (27.2) |
| 3 (42.3%–49.8%) | 31,748 (21.0) | 9,320 (21.7) | 2,125 (19.4) | 63,372 (20.0) | 17,913 (20.8) | 7,484 (19.8) | 44,474 (18.5) | 10,758 (16.7) | 3,146 (17.7) | 7,008 (20.2) | 1,841 (14.8) | 1,134 (19.2) |
| 4 (50.0%–57.5%) | 28,457 (18.8) | 8,890 (20.7) | 1,817 (16.6) | 54,441 (17.2) | 15,440 (17.9) | 6,376 (16.9) | 37,449 (15.6) | 10,799 (16.8) | 2,585 (14.6) | 6,668 (19.3) | 1,926 (15.4) | 1,051 (17.8) |
| 5 (57.5%–100%) | 24,417 (16.2) | 7,707 (17.9) | 1,408 (12.9) | 45,364 (14.3) | 14,195 (16.5) | 5,873 (15.5) | 31,525 (13.1) | 10,940 (17.0) | 2,228 (12.6) | 5,576 (16.1) | 1,701 (13.6) | S |
| Unknown/missing | 876 (0.6) | 361 (0.8) | 32 (0.3) | - | - | - | 51,155 (21.3) | | 1,4831 (23.0) | 17 (0.0) | 130 (1.0) | ≤6 (≤0.1) |
| Persons per dwelling quintile[e] | | | | | | | | | | | | |
| 1 (0–2.1) | 25,910 (17.1) | 7,635 (17.8) | 1,974 (18.0) | 63,105 (19.9) | 16,356 (19.0) | 7,593 (20.1) | 52,139 (21.7) | 14,984 (23.3) | 3,126 (17.6) | 8,770 (25.3) | 2,926 (23.4) | 1,481 (25.0) |

| | | | | | | | | | | | | |
|---|---|---|---|---|---|---|---|---|---|---|---|---|
| 2 (2.2–2.4) | 25,042 (16.6) | 7,891 (18.4) | 1,859 (17.0) | 47,085 (14.9) | 13,184 (15.3) | 5,892 (15.6) | 39,467 (16.4) | 13,809 (21.5) | 2,285 (12.9) | 5,549 (16.0) | 1,933 (15.5) | 1,068 (18.1) |
| 3 (2.5–2.6) | 18,749 (12.4) | 5,286 (12.3) | 1,527 (14.0) | 53,579 (16.9) | 15,825 (18.4) | 6,660 (17.6) | 43,367 (18.1) | 11,104 (17.3) | 2,878 (16.2) | 6,013 (17.4) | 1,729 (13.9) | 1,042 (17.6) |
| 4 (2.7–3.0) | 36,653 (24.3) | 9,683 (22.5) | 2,825 (25.8) | 78,052 (24.7) | 21,501 (24.9) | 9,519 (25.2) | 45,389 (18.9) | 9,808 (15.2) | 3,959 (22.3) | 7,977 (23.0) | 1,834 (14.7) | 1,410 (23.8) |
| 5 (3.1–5.7) | 43,798 (29.0) | 12,123 (28.2) | 2,722 (24.9) | 74,714 (23.6) | 19,311 (22.4) | 8,122 (21.5) | 51,428 (21.4) | 11,440 (17.8) | 4,898 (27.6) | 5,787 (16.7) | 3,814 (30.6) | S |
| Unknown/missing | 934 (0.6) | 376 (0.9) | 33 (0.3) | - | - | - | 8,253 (3.4) | 3,222 (5.0) | 602 (3.4) | 17 (0.0) | 130 (1.0) | ≤6 (≤0.1) |
| Self-identified visible minority quintile[f] | | | | | | | | | | | | |
| 1 (0.0%–2.2%) | 21,763 (14.4) | 9,003 (20.9) | 1,316 (12.0) | 69,145 (21.8) | 23,454 (27.2) | 9,774 (25.9) | 26,250 (10.9) | 11948 (18.6) | 1004 (5.7) | 3,227 (9.3) | 4,116 (33.0) | 683 (11.5) |
| 2 (2.2%–7.5%) | 25,841 (17.1) | 7,853 (18.3) | 1,883 (17.2) | 46,787 (14.8) | 13,945 (16.2) | 6,334 (16.8) | 42,342 (17.6) | 12,551 (19.5) | 2,264 (12.8) | 5,889 (17.0) | 3,042 (24.4) | 1,223 (20.7) |
| 3 (7.5%–18.7%) | 27,528 (18.2) | 6,988 (16.3) | 2,450 (22.4) | 71,599 (22.6) | 18,986 (22.0) | 9,414 (24.9) | 50,965 (21.2) | 11,798 (18.3) | 4,221 (23.8) | 7,481 (21.6) | 2,046 (16.4) | 1,419 (24.0) |
| 4 (18.7%–43.5%) | 32,008 (21.2) | 7,955 (18.5) | 2,760 (25.2) | 70,264 (22.2) | 16,054 (18.6) | 7,702 (20.4) | 57,748 (24.1) | 12,917 (20.1) | 5,456 (30.7) | 8,471 (24.5) | 1,703 (13.6) | 1,372 (23.2) |
| 5 (43.5%–100%) | 43,070 (28.5) | 10,834 (25.2) | 2,501 (22.9) | 58,740 (18.6) | 13,738 (15.9) | 4,562 (12.1) | 54,723 (22.8) | 12,050 (18.7) | 4,232 (23.8) | 9,028 (26.1) | 1,329 (10.6) | S |
| Unknown/missing | 876 (0.6) | 361 (0.8) | 30 (0.3) | - | - | - | 8,015 (3.3) | 3,103 (4.8) | 571 (3.2) | 17 (0.0) | 130 (1.0) | ≤6 (≤0.1) |

S = suppressed; counts are suppressed because they are either small cell counts (≤6, which cannot be disclosed because of privacy and data obligations) or they could be derived using other estimates in this table.
[a]Proportion reported, unless stated otherwise.
[b]Comorbidities include chronic respiratory diseases, chronic heart diseases, hypertension, diabetes, immunocompromising conditions due to underlying diseases or therapy, autoimmune diseases, chronic kidney disease, advanced liver disease, dementia/frailty and history of stroke or transient ischemic attack.
[c]Neighbourhood income quintile has variable cut-off values in each city/Census area to account for cost of living. A dissemination area (DA) being in quintile 1 means it is among the lowest 20% of DAs in its city by income. Material deprivation index quintile used in British Columbia; quintile 1 represents 'most deprived' and quintile 5 represents 'least deprived'.
[d]Percentage of people in the area working in the following occupations: sales and service occupations; trades, transport and equipment operators and related occupations; natural resources, agriculture, and related production occupations; and occupations in manufacturing and utilities. Census counts for people are randomly rounded up or down to the nearest number divisible by 5, which causes some minor imprecision.
[e]Range of persons per dwelling.
[f]Percentage of people in the area who self-identified as a visible minority. Census counts for people are randomly rounded up or down to the nearest number divisible by 5, which causes some minor imprecision.

**Supplemental Table S7: Province-specific and pooled vaccine effectiveness ≥14 days after a first dose and ≥7 days after receiving a second dose against hospitalization and death in Ontario, Quebec, British Columbia, and Manitoba**

| | Hospitalization | | | | Death | | | |
|---|---|---|---|---|---|---|---|---|
| | **Test-positive vaccinated cases / total test-positive cases** | **Test-negative vaccinated controls, vaccinated / total test-negative controls** | **Unadjusted vaccine effectiveness (95% CI)** | **Adjusted[a] vaccine effectiveness (95% CI)** | **Test-positive vaccinated cases / total test-positive cases** | **Test-negative vaccinated controls, vaccinated / total test-negative controls** | **Unadjusted vaccine effectiveness (95% CI)** | **Adjusted[a] vaccine effectiveness (95% CI)** |
| **Ontario** | | | | | | | | |
| ≥14 days after dose 1 | 1,289 / 15,265 | 60,443 / 36,8117 | 53 (50–56) | 82 (81–83) | 267 / 2,947 | 61,465 / 380,435 | 48 (41–54) | 79 (76–82) |
| ≥7 days after dose 2 | 270 / 14,246 | 148,552 / 456,226 | 96 (95–96) | 98 (98–98) | 58 / 2,738 | 148,764 / 467,734 | 95 (94–96) | 97 (95–97) |
| **Quebec** | | | | | | | | |
| ≥14 days after dose 1 | 465 / 6,673 | 169,766 / 676,662 | 78 (75–80) | 87 (85–88) | 108 / 1,419 | 169,766 / 676,662 | 75 (70–80) | 89 (86–92) |
| ≥7 days after dose 2 | 164 / 6,372 | 217,206 / 724,102 | 94 (93–95) | 99 (99–100) | 23 / 1,334 | 217,206 / 724,102 | 96 (94–97) | 100 (99–100) |
| **British Columbia** | | | | | | | | |
| ≥14 days after dose 1 | 520 / 5,092 | 108,127 / 630,534 | 45 (40–50) | 76 (73–78) | 95 / 681 | 108,127 / 630,534 | 22 (3–37) | 69 (60–76) |
| ≥7 days after dose 2 | 221 / 4,793 | 197,161 / 719,568 | 87 (85–89) | 98 (97–98) | 34 / 620 | 197,161 / 719,568 | 85 (78–89) | 97 (96–98) |
| **Manitoba** | | | | | | | | |
| ≥14 days after dose 1 | 168 / 1,955 | 12,616 / 78,272 | 51 (43–58) | 85 (82–87) | 24 / 294 | 12,760 / 79,933 | 54 (30–70) | 89 (83–93) |
| ≥7 days after dose 2 | 58 / 1,845 | 25,051 / 90,707 | 92 (89–93) | 97 (96–98) | 9 / 279 | 25,100 / 92,273 | 91 (83–96) | 97 (94–99) |
| **Pooled** | | | | | | | | |
| ≥14 days after dose 1 | 2,442 / 28,985 | 350,952 / 1,753,585 | 59 (39–73) | 83 (78–87) | 494 / 5341 | 352,118 / 1,767,564 | 54 (25–71) | 83 (72–90) |
| ≥7 days after dose 2 | 713 / 27,256 | 587,970 / 1,990,603 | 93 (88–96) | 98 (96–99) | 124 / 4971 | 588,231 / 2,003,677 | 93 (87–96) | 98 (95–99) |

[a]Models adjusted for age group, sex, public health unit region, biweekly period of test, number of SARS-CoV-2 tests in the 3 months prior to 14 December 2020, presence of any comorbidity that increase the risk of severe COVID-19, receipt of influenza vaccination in current or prior influenza season, and neighbourhood income (material deprivation index in British Columbia), proportion of persons employed as non-health essential workers, persons per dwelling, and self-identified visible minority quintiles. Receipt of influenza vaccination was not adjusted for in British Columbia because of lack of data.

**Supplemental Table S8: Province-specific and pooled vaccine effectiveness against severe outcomes of hospitalization or death for mRNA and ChAdOx1 vaccines in Ontario, Quebec, British Columbia, and Manitoba**

| mRNA vaccines (BNT162b2/mRNA-1273) | | | | | ChAdOx1 vaccines (AstraZeneca Vaxzevria/COVISHIELD) | | | | |
|---|---|---|---|---|---|---|---|---|---|
| **Interval from vaccine dose to index date** | **Test-positive vaccinated cases / total test-positive cases** | **Test-negative vaccinated controls, vaccinated / total test-negative controls** | **Unadjusted vaccine effectiveness (95% CI)** | **Adjusted[a] vaccine effectiveness (95% CI)** | **Interval from vaccine dose to index date** | **Test-positive vaccinated cases / total test-positive cases** | **Test-negative vaccinated controls, vaccinated / total test-negative controls** | **Unadjusted vaccine effectiveness (95% CI)** | **Adjusted[a] vaccine effectiveness (95% CI)** |
| **Ontario** | | | | | | | | | |
| **Dose 1** | | | | | **Dose 1** | | | | |
| 0–13 days | 1,116 / 15,694 | 16,346 / 323,418 | -44 (-53 to -35) | 27 (22–32) | 0-13 days | 97 / 14,675 | 1,845 / 308,917 | -11 (-36 to 10) | 49 (37–59) |
| 14–27 days | 540 / 151,18 | 17,591 / 324,663 | 35 (29–41) | 74 (71–76) | 14-27 days | 65 / 14,643 | 1,833 / 308,905 | 25 (4–42) | 73 (65–79) |
| 28–55 days | 442 / 15,020 | 23,100 / 330,172 | 60 (56–63) | 86 (84–87) | 28-55 days | 35 / 14,613 | 2,455 / 309,527 | 70 (58–78) | 91 (88–94) |
| 56–83 days | 166 / 14,744 | 10,070 / 317,142 | 65 (59–70) | 89 (87–91) | ≥56 days | 16 / 14,594 | 1,279 / 308,351 | 74 (57–84) | 93 (88–96) |
| ≥84 days | 66 / 14,644 | 4,074 / 311,146 | 66 (56–73) | 86 (81–89) | | | | | |
| **Dose 2** | | | | | **Dose 2** | | | | |
| 0–6 days | 31 / 14,609 | 5,325 / 312,397 | 88 (83–91) | 92 (88–94) | 0-6 days | Suppressed | 60 / 307,132 | 65 (-153 to 95) | 90 (22–99) |
| 7–55 days | 104 / 14,682 | 56,989 / 364,061 | 96 (95–97) | 98 (97–98) | 7-55 days | Suppressed | 1017 / 308,089 | 88 (72–94) | 94 (87–98) |
| 56–111 days | 103 / 14,681 | 66,872 / 373,944 | 97 (96–97) | 99 (98–99) | ≥56 days | 14 / 14592 | 2217 / 309,289 | 87 (77–92) | 96 (94–98) |
| ≥112 days | 50 / 14,628 | 12,729 / 319,801 | 92 (89–94) | 96 (95–97) | | | | | |
| **Quebec** | | | | | | | | | |
| **Dose 1** | | | | | **Dose 1** | | | | |
| 0–13 days | 339 / 7,153 | 35,354 / 542,250 | 29 (20–36) | 50 (44–56) | 0-13 days | 40 / 6,854 | 4,518 / 511,414 | 34 (10–52) | 37 (14–55) |
| 14–27 days | 159 / 6,973 | 37,508 / 544,404 | 68 (63–73) | 81 (78–84) | 14-27 days | 22 / 6,836 | 3,877 / 510,773 | 58 (36–72) | 68 (52–80) |
| 28–55 days | 152 / 6,966 | 66,573 / 573,469 | 83 (80–86) | 89 (86–90) | 28-55 days | 18 / 6,832 | 5,422 / 512,318 | 75 (61–84) | 85 (76–91) |
| 56–83 days | 88 / 6,902 | 35,244 / 542,140 | 81 (77–85) | 90 (88–92) | ≥56 days | 8 / 6,822 | 4,253 / 511,149 | 86 (72–93) | 94 (87–97) |
| ≥84 days | 38 / 6,852 | 16,889 / 523,785 | 83 (77–88) | 95 (93–97) | | | | | |
| **Dose 2** | | | | | **Dose 2** | | | | |

| | | | | | | | | | |
|---|---|---|---|---|---|---|---|---|---|
| 0–6 days | 8 / 6,822 | 11,704 / 518,600 | 95 (90–97) | 98 (95–99) | 0-6 days | 1 / 6,815 | 261 / 507,157 | 71 (-103 to 96) | 84 (-18 to 98) |
| 7–55 days | 51 / 6,865 | 100,440 / 607,336 | 96 (95–97) | 99 (99–99) | 7-55 days | 0 / 6,814 | 2,675 / 509,571 | Not reliably estimable | Not reliably estimable |
| 56–111 days | 87 / 6,901 | 88,104 / 595,000 | 93 (91–94) | 100 (100–100) | ≥56 days | 12 / 6,826 | 4,175 / 511,071 | 79 (62–88) | 99 (99–100) |
| ≥112 days | 11 / 6,825 | 9,932 / 516,828 | 92 (85–95) | 99 (99–100) | | | | | |
| **British Columbia** | | | | | **Dose 1** | | | | |
| **Dose 1** | | | | | | | | | |
| 0–13 days | 351 / 5,094 | 30,697 / 55,3104 | -26 (-40 to -13) | 30 (21–37) | 0-13 days | 34 / 4,777 | 3,441 / 525,848 | -9 (-53 to 22) | 14 (-21 to 39) |
| 14–27 days | 163 / 4,906 | 29,428 / 551,835 | 39 (29–48) | 71 (65–75) | 14-27 days | 19 / 4,762 | 3,115 / 525,522 | 33 (-6 to 57) | 55 (28–71) |
| 28–55 days | 181 / 4,924 | 42,624 / 565,031 | 53 (46–60) | 79 (76–82) | 28-55 days | 7 / 4,750 | 4,381 / 526,788 | 82 (63–92) | 89 (77–95) |
| 56–83 days | 93 / 4,836 | 18,845 / 541,252 | 46 (33–56) | 80 (75–84) | ≥56 days | 4 / 4,747 | 1,894 / 524,301 | 77 (38–91) | 87 (64–95) |
| ≥84 days | 81 / 4,824 | 7,840 / 530,247 | -14 (-42 to 9) | 68 (60–75) | | | | | |
| **Dose 2** | | | | | **Dose 2** | | | | |
| 0–6 days | 18 / 4,761 | 8,075 / 530,482 | 75 (61–85) | 89 (83–93) | 0-6 days | 0 / 4,743 | 180 / 522,587 | -[b] | -[b] |
| 7–55 days | 64 / 4,807 | 84,263 / 606,670 | 92 (89–93) | 98 (97–98) | 7-55 days | 3 / 4,746 | 1,777 / 524,184 | 81 (42–94) | 92 (75–97) |
| 56–111 days | 124 / 4,867 | 82,976 / 605,383 | 84 (80–86) | 98 (97–98) | ≥56 days | 12 / 4,755 | 4,397 / 526,804 | 70 (47–83) | 94 (90–97) |
| ≥112 days | 29 / 4,772 | 13,586 / 535,993 | 76 (66–84) | 96 (95–98) | | | | | |
| **Manitoba** | | | | | | | | | |
| **Dose 1** | | | | | **Dose 1** | | | | |
| 0–13 days | 89 / 1,944 | 4,365 / 69,953 | 28 (11–42) | 61 (51–69) | 0-13 days | 12 / 1867 | 659 /66247 | 36 (-14 to 64) | 39 (-10 to 66) |
| 14–27 days | 49 / 1,904 | 4,435 / 70,023 | 61 (48–71) | 84 (78–88) | 14-27 days | 6 / 1861 | 592 /66180 | 64 (20–84) | 78 (50–90) |
| 28–55 days | 56 / 1,911 | 4,733 / 70,321 | 58 (45–68) | 88 (85–91) | 28-55 days | 17 / 1872 | 914 /66502 | 34 (-6 to 59) | 72 (55–83) |
| 56–83 days | 24 / 1,879 | 1,214 / 66,802 | 30 (-5 to 53) | 87 (80–91) | ≥56 days | 13 / 1868 | 422 /66010 | -9 (-90 to 37) | 70 (46–83) |
| ≥84 days | Suppressed / 1,860 | 304 / 65,892 | 42 (-41 to 76) | 87 (67–95) | | | | | |
| **Dose 2** | | | | | **Dose 2** | | | | |
| 0–6 days | 7 / 1,862 | 1,232 / 66,820 | 80 (58–90) | 93 (86–97) | 0-6 days | Suppressed / 1,855 | 11 / 65,599 | No cases | No cases |
| 7–55 days | 37 / 1,892 | 10,919 / 76,507 | 88 (83–91) | 95 (93–96) | 7-55 days | Suppressed / 1,855 | 74 / 65,662 | No cases | No cases |

| | | | | | | | | | |
|---|---|---|---|---|---|---|---|---|---|
| 56–111 days | 19 / 1,874 | 15,375 / 80,963 | 96 (93–97) | 98 (97–99) | ≥56 days | Suppressed / 1,856 | 189 /65,777 | 81 (-34 to 97) | 89 (18–98) |
| ≥112 days | 8 / 1,863 | 4,229 / 69,817 | 93 (87–97) | 98 (97–99) | | | | | |
| **Pooled** | | | | | | | | | |
| **Dose 1** | | | | | **Dose 1** | | | | |
| 0–13 days | 1,895 / 29,885 | 86,762 / 1,488,725 | 1 (-42 to 31) | 43 (25–57) | 0-13 days | 183 / 28,173 | 10,463 / 1,412,426 | 12 (-18 to 34) | 37 (20–51) |
| 14–27 days | 911 / 28,901 | 88,962 / 1,490,925 | 53 (33–67) | 78 (71–83) | 14-27 days | 112 / 28,102 | 9,417 / 1,411,380 | 43 (20–59) | 69 (60–76) |
| 28–55 days | 831 / 28,821 | 137,030 / 1,538,993 | 66 (46–79) | 86 (81–89) | 28-55 days | 77 / 28,067 | 13,172 / 1,415,135 | 69 (47–81) | 86 (77–92) |
| 56–83 days | 371 / 28,361 | 65,373 / 1,467,336 | 61 (31–78) | 87 (82–91) | ≥56 days | 41 / 28,031 | 7,848 / 1,409,811 | 68 (22–87) | 88 (75–94) |
| ≥84 days | >185 / 28,180 | 29,107 / 1,431,070 | 56 (0–81) | 87 (71–94) | | | | | |
| **Dose 2** | | | | | **Dose 2** | | | | |
| 0–6 days | 64 / 28,054 | 26,336 / 1,428,299 | 87 (74–93) | 93 (88–96) | 0-6 days | >1 / >13413 | 512 / 1,402,475 | 68 (-24 to 92) | 87 (48–97) |
| 7–55 days | 256 / 28,246 | 252,611 / 1,654,574 | 94 (89–96) | 98 (95–99) | 7-55 days | >3 / >13415 | 5,543 / 1,407,506 | 86 (73–92) | 94 (88–97) |
| 56–111 days | 333 / 28,323 | 253,327 / 1,655,290 | 94 (87–97) | 99 (97–99) | ≥56 days | >38 / 28,029 | 10,978 / 1,412,941 | 80 (69–87) | 97 (91–99) |
| ≥112 days | 98 / 28,088 | 40,476 / 1,442,439 | 90 (81–94) | 98 (96–99) | | | | | |

[a]Models adjusted for age group, sex, public health unit region, biweekly period of test, number of SARS-CoV-2 tests in the 3 months prior to 14 December 2020, presence of any comorbidity that increase the risk of severe COVID-19, receipt of influenza vaccination in current or prior influenza season, and neighbourhood income (material deprivation index in British Columbia), proportion of persons employed as non-health essential workers, persons per dwelling, and self-identified visible minority quintiles. Receipt of influenza vaccination was not adjusted for in British Columbia because of lack of data.

[b]VE estimated as 100% based on zero vaccinated test-positive cases.

**Supplemental Table S9: Province-specific and pooled vaccine effectiveness against severe outcomes of hospitalization or death ≥14 days after a first dose (for subjects who received only 1 dose) and ≥7 days after receiving a second dose by subject characteristics in Ontario, Quebec, British Columbia, and Manitoba**

| | **Test-positive vaccinated cases / total test-positive cases** | **Test-negative vaccinated controls, vaccinated / total test-negative controls** | **Unadjusted vaccine effectiveness (95% CI)** | **Adjusted[a] vaccine effectiveness (95% CI)** |
|---|---|---|---|---|
| **Ontario** | | | | |
| **Age group** | | | | |
| 18–59 years | | | | |
| ≥14 days after dose 1 | 211 / 6,926 | 42,102 / 300,262 | 81 (78–83) | 89 (88–91) |
| ≥7 days after dose 2 | 62 / 6,777 | 117,241 / 375,401 | 98 (97–98) | 99 (98–99) |
| 60–69 years | | | | |
| ≥14 days after dose 1 | 227 / 3,123 | 8,871 / 36,468 | 76 (72–79) | 89 (87–90) |
| ≥7 days after dose 2 | 54 / 2,950 | 15,913 / 43,510 | 97 (96–98) | 98 (97–98) |
| 70–79 years | | | | |
| ≥14 days after dose 1 | 341 / 2,844 | 5,645 / 19,328 | 67 (63–71) | 80 (77–83) |
| ≥7 days after dose 2 | 56 / 2,559 | 8,970 / 22,653 | 97 (96–97) | 97 (96–98) |
| ≥80 years | | | | |
| ≥14 days after dose 1 | 551 / 3,015 | 3,784 / 11,416 | 55 (50–59) | 65 (59–70) |
| ≥7 days after dose 2 | 111 / 2,575 | 6,415 / 14,047 | 95 (93–96) | 94 (92–95) |
| **Sex** | | | | |
| Females | | | | |
| ≥14 days after dose 1 | 597 / 7,051 | 36,152 / 207,591 | 56 (52–60) | 84 (83–86) |
| ≥7 days after dose 2 | 131 / 6,585 | 90,655 / 262,094 | 96 (95–97) | 98 (98–98) |
| Males | | | | |
| ≥14 days after dose 1 | 733 / 8,857 | 24,250 / 159,883 | 50 (46–53) | 81 (80–83) |
| ≥7 days after dose 2 | 152 / 8,276 | 57,884 / 193,517 | 96 (95–96) | 98 (97–98) |
| **Presence of any comorbidity** | | | | |
| Yes | | | | |
| ≥14 days after dose 1 | 1,220 / 12,061 | 31,840 / 168,492 | 52 (49–55) | 80 (78–81) |
| ≥7 days after dose 2 | 254 / 11,095 | 68,926 / 205,578 | 95 (95–96) | 97 (97–98) |
| No | | | | |
| ≥14 days after dose 1 | 110 / 3,847 | 28,562 / 198,982 | 82 (79–85) | 92 (90–93) |
| ≥7 days after dose 2 | 29 / 3,766 | 79,613 / 250,033 | 98 (98–99) | 99 (99–99) |
| **Quebec** | | | | |
| **Age group** | | | | |
| 18–59 years | | | | |
| ≥14 days after dose 1 | 58 / 2,807 | 116,676 / 525,777 | 93 (90–94) | 94 (93–96) |
| ≥7 days after dose 2 | 40 / 2,789 | 149,114 / 558,215 | 96 (95–97) | 100 (99–100) |
| 60–69 years | | | | |
| ≥14 days after dose 1 | 85 / 1,421 | 24,606 / 83,330 | 85 (81–88) | 89 (86–92) |
| ≥7 days after dose 2 | 31 / 1,367 | 34,679 / 93,403 | 96 (94–97) | 100 (99–100) |
| 70–79 years[b] | | | | |
| ≥14 days after dose 1 | 155 / 1,461 | 18,112 / 45,860 | 82 (79–85) | 86 (82–89) |
| ≥7 days after dose 2 | 41 / 1,347 | 22,471 / 50,219 | 96 (95–97) | 100 (99–100) |
| ≥80 years | | | | |
| ≥14 days after dose 1 | 187 / 1,610 | 10,372 / 21,695 | 86 (83–88) | 84 (79–87) |
| ≥7 days after dose 2 | 55 / 1,478 | 10,942 / 22,265 | 96 (95–97) | 99 (98–99) |
| **Sex** | | | | |
| Females | | | | |
| ≥14 days after dose 1 | 195 / 3,167 | 105,664 / 403,931 | 81 (79–84) | 90 (88–92) |
| ≥7 days after dose 2 | 70 / 3,042 | 131,065 / 429,332 | 95 (93–96) | 100 (99–100) |
| Males | | | | |
| ≥14 days after dose 1 | 290 / 4,132 | 64,102 / 272,731 | 75 (72–78) | 86 (84–88) |
| ≥7 days after dose 2 | 97 / 3,939 | 86,141 / 294,770 | 94 (93–95) | 99 (99–100) |
| **Presence of any comorbidity** | | | | |
| Yes | | | | |
| ≥14 days after dose 1 | 418 / 4,846 | 62,020 / 207,204 | 78 (76–80) | 85 (83–87) |
| ≥7 days after dose 2 | 136 / 4,564 | 77,763 / 222,947 | 94 (93–95) | 99 (99–99) |
| No | | | | |
| ≥14 days after dose 1 | 67 / 2,453 | 107,746 / 469,458 | 91 (88–93) | 94 (92–95) |
| ≥7 days after dose 2 | 31 / 2,417 | 139,443 / 501,155 | 97 (95–98) | 100 (100, 100) |

| | | | | |
|---|---|---|---|---|
| **British Columbia** | | | | |
| **Age group** | | | | |
| 18–59 years | | | | |
| ≥14 days after dose 1 | 140 / 2,550 | 73,013 / 500,757 | 66 (60–71) | 79 (75–82) |
| ≥7 days after dose 2 | 49 / 2,459 | 139,843 / 567,587 | 94 (92–95) | 98 (98–99) |
| 60–69 years | | | | |
| ≥14 days after dose 1 | 104 / 1,094 | 14,348 / 66,674 | 62 (53–69) | 78 (72–83) |
| ≥7 days after dose 2 | 49 / 1,039 | 26,174 / 78,500 | 90 (87–93) | 98 (97–98) |
| 70–79 years | | | | |
| ≥14 days after dose 1 | 132 / 905 | 11,527 / 39,063 | 59 (51–66) | 75 (68–80) |
| ≥7 days after dose 2 | 61 / 834 | 18,634 / 46,170 | 88 (85–91) | 98 (97–98) |
| ≥80 years | | | | |
| ≥14 days after dose 1 | 172 / 742 | 9,239 / 24,040 | 52 (43–59) | 69 (61–75) |
| ≥7 days after dose 2 | 75 / 645 | 12,510 / 27,311 | 84 (80–88) | 94 (92–96) |
| **Sex** | | | | |
| Females | | | | |
| ≥14 days after dose 1 | 224 / 2,257 | 6,332 / 282,650 | 52 (45–58) | 78 (74–81) |
| ≥7 days after dose 2 | 93 / 2,126 | 115,220 / 391,538 | 89 (86–91) | 98 (97–98) |
| Males | | | | |
| ≥14 days after dose 1 | 324 / 3,034 | 44,795 / 290,884 | 34 (26–41) | 73 (70–77) |
| ≥7 days after dose 2 | 141 / 2,851 | 81,941 / 328,030 | 84 (81–87) | 97 (97–98) |
| **Presence of any comorbidity** | | | | |
| Yes | | | | |
| ≥14 days after dose 1 | 465 / 3,441 | 49,921 / 226,338 | 45 (39–50) | 70 (67–73) |
| ≥7 days after dose 2 | 206 / 3,182 | 82,686 / 259,103 | 85 (83–87) | 97 (96–97) |
| No | | | | |
| ≥14 days after dose 1 | 83 / 1,850 | 58,206 / 404,196 | 72 (65–78) | 85 (82–88) |
| ≥7 days after dose 2 | 28 / 1,795 | 114,475 / 460,465 | 95 (93–97) | 99 (98–99) |
| **Manitoba** | | | | |
| **Age group** | | | | |
| 18–59 years | | | | |
| ≥14 days after dose 1 | 62 / 1,218 | 8,823 / 63,818 | 67 (57–74) | 86 (82–89) |
| ≥7 days after dose 2 | 24 / 1,180 | 26,497 / 81,492 | 96 (94–97) | 98 (96–98) |
| 60–69 years | | | | |
| ≥14 days after dose 1 | 39 / 326 | 2,001 / 7,985 | 59 (43–71) | 86 (78–90) |
| ≥7 days after dose 2 | 19 / 306 | 3,809 / 9,793 | 90 (83–93) | 96 (93–98) |
| 70–79 years | | | | |
| ≥14 days after dose 1 | 37 / 278 | 1,228 / 4,230 | 62 (47–74) | 90 (84–94) |
| ≥7 days after dose 2 | 9 / 250 | 2,057 / 5,059 | 95 (89–97) | 99 (97–99) |
| ≥80 years | | | | |
| ≥14 days after dose 1 | 32 / 203 | 571 / 2,178 | 47 (22–64) | 73 (54–84) |
| ≥7 days after dose 2 | 16 / 187 | 1,343 / 2,950 | 89 (81–93) | 95 (90–97) |
| **Sex** | | | | |
| Females | | | | |
| ≥14 days after dose 1 | 83 / 1,102 | 7,255 / 43,584 | 59 (49–67) | 87 (83–90) |
| ≥7 days after dose 2 | 34 / 1,053 | 20,206 / 56,535 | 94 (92–96) | 98 (96–98) |
| Males | | | | |
| ≥14 days after dose 1 | 87 / 923 | 5,368 / 34,627 | 43 (29–55) | 83 (78–87) |
| ≥7 days after dose 2 | 34 / 870 | 13,500 / 42,759 | 91 (88–94) | 97 (95–98) |
| **Presence of any comorbidity** | | | | |
| Yes | | | | |
| ≥14 days after dose 1 | 143 / 1,303 | 5,925 / 29,773 | 50 (41–58) | 84 (80–87) |
| ≥7 days after dose 2 | 56 / 1,216 | 13,372 / 37,220 | 91 (89–93) | 96 (95–97) |
| No | | | | |
| ≥14 days after dose 1 | 27 / 722 | 6,698 / 48,438 | 76 (64–84) | 90 (86–94) |
| ≥7 days after dose 2 | 12 / 707 | 20,334 / 62,074 | 96 (94–98) | 98 (97–99) |
| **Pooled** | | | | |
| **Age group** | | | | |
| 18–59 years | | | | |
| ≥14 days after dose 1 | 471 / 13,501 | 240,614 / 139,0614 | 80 (60–90) | 88 (80–93) |
| ≥7 days after dose 2 | 175 / 13,205 | 432,695 / 1,582,695 | 96 (94–98) | 99 (98–99) |
| 60–69 years | | | | |
| ≥14 days after dose 1 | 455 / 5,964 | 49,826 / 194,457 | 73 (57–82) | 86 (80–90) |
| ≥7 days after dose 2 | 153 / 5,662 | 80,575 / 225,206 | 94 (89–97) | 98 (96–99) |

| | | | | |
|---|---|---|---|---|
| 70–79 years | | | | |
| ≥14 days after dose 1 | 665 / 5,488 | 36,512 / 108,481 | 69 (56–79) | 83 (76–88) |
| ≥7 days after dose 2 | 167 / 4,990 | 52,132 / 124,101 | 95 (90–97) | 99 (97–99) |
| ≥80 years | | | | |
| ≥14 days after dose 1 | 942 / 5,570 | 23,966 / 59,329 | 64 (35–81) | 73 (62–81) |
| ≥7 days after dose 2 | 257 / 4,885 | 31,210 / 66,573 | 92 (86–96) | 96 (92–98) |
| **Sex** | | | | |
| Females | | | | |
| ≥14 days after dose 1 | 1,099 / 13,577 | 155,403 / 937,756 | 64 (45–77) | 85 (80–89) |
| ≥7 days after dose 2 | 328 / 12,806 | 357,146 / 1,139,499 | 94 (91–96) | 99 (97–99) |
| Males | | | | |
| ≥14 days after dose 1 | 1,434 / 16,946 | 138,515 / 758,125 | 54 (29–70) | 81 (76–86) |
| ≥7 days after dose 2 | 424 / 15,936 | 239,466 / 859,076 | 92 (87–95) | 98 (96–99) |
| **Presence of any comorbidity** | | | | |
| Yes | | | | |
| ≥14 days after dose 1 | 2,246 / 21,651 | 149,706 / 631,807 | 59 (37–73) | 80 (73–85) |
| ≥7 days after dose 2 | 652 / 20,057 | 242,747 / 724,848 | 92 (87–95) | 98 (95–99) |
| No | | | | |
| ≥14 days after dose 1 | 287 / 8,872 | 201,212 / 1,121,074 | 82 (71–89) | 91 (87–94) |
| ≥7 days after dose 2 | 100 / 8,685 | 353,865 / 1,273,727 | 97 (95–98) | 99 (98–100) |

[a]Models adjusted for age group, sex, public health unit region, biweekly period of test, number of SARS-CoV-2 tests in the 3 months prior to 14 December 2020, presence of any comorbidity that increase the risk of severe COVID-19, receipt of influenza vaccination in current or prior influenza season, and neighbourhood income (material deprivation index in British Columbia), proportion of persons employed as non-health essential workers, persons per dwelling, and self-identified visible minority quintiles. Receipt of influenza vaccination was not adjusted for in British Columbia because of lack of data.

[b]Model adjusted for sex, biweekly period of test, presence of any comorbidity that increase the risk of severe COVID-19, number of SARS-CoV-2 tests in previous 3 months, and receipt of influenza vaccination in current or prior influenza season.

**Supplemental Table S10: Province-specific and pooled vaccine effectiveness against severe outcomes of hospitalization or death ≥14 days after a first dose (for subjects who received only 1 dose) and ≥7 days after receiving a second dose by vaccine product in Ontario, Quebec, British Columbia, and Manitoba**

| | Test-positive vaccinated cases / total test-positive cases | Test-negative vaccinated controls, vaccinated / total test-negative controls | Unadjusted vaccine effectiveness (95% CI) | Adjusted[a] vaccine effectiveness (95% CI) |
|---|---|---|---|---|
| **Ontario** | | | | |
| BNT162b2 | | | | |
| ≥14 days after dose 1 | 1,044 / 15,622 | 43,712 / 350,784 | 50 (46–53) | 82 (81–84) |
| ≥7 days after dose 2 | 201 / 14,779 | 89,149 / 396,221 | 95 (95–96) | 98 (97–98) |
| mRNA-1273 | | | | |
| ≥14 days after dose 1 | 170 / 14,748 | 11,123 / 318,195 | 68 (63–72) | 85 (82–87) |
| ≥7 days after dose 2 | 44 / 14,622 | 26,496 / 333,568 | 97 (95–97) | 98 (97–99) |
| BNT162b2/mRNA-1273 | | | | |
| ≥14 days after dose 1 | N/A | N/A | N/A | N/A |
| ≥7 days after dose 2 | Suppressed | 20,945 / 328,017 | 99 (98–99) | 99 (98–99) |
| ChAdOx1 | | | | |
| ≥14 days after dose 1 | 116 / 14,694 | 5,567 / 312,639 | 56 (47–64) | 86 (83–89) |
| ≥7 days after dose 2 | 20 / 14,598 | 3,234 / 310,306 | 87 (80–92) | 96 (94–97) |
| ChAdOx1/mRNA mixed schedule | | | | |
| ≥14 days after dose 1 | N/A | N/A | N/A | N/A |
| ≥7 days after dose 2 | 6 / 14,584 | 8,715 / 315,787 | 99 (97–99) | 99 (98–100) |
| **Quebec** | | | | |
| BNT162b2 | | | | |
| ≥14 days after dose 1 | 375 / 7,189 | 123,899 / 630,795 | 77 (75–80) | 87 (86–89) |
| ≥7 days after dose 2 | 116 / 6,930 | 151,606 / 658,502 | 94 (93–95) | 100 (99–100) |
| mRNA-1273 | | | | |
| ≥14 days after dose 1 | 62 / 6,876 | 32,315 / 539,211 | 86 (82–89) | 92 (90–94) |
| ≥7 days after dose 2 | 26 / 6,840 | 41,389 / 548,285 | 95 (93–97) | 100 (99–100) |
| BNT162b2/mRNA-1273 | | | | |
| ≥14 days after dose 1 | N/A | N/A | | |
| ≥7 days after dose 2 | 7 / 6,821 | 5,481 / 512,377 | 91 (80–95) | 99 (98–100) |
| ChAdOx1 | | | | |
| ≥14 days after dose 1 | 48 / 6,862 | 13,552 / 520,448 | 74 (65–80) | 85 (79–89) |
| ≥7 days after dose 2 | 12 / 6,826 | 6,850 / 513,746 | 87 (77–93) | 99 (99–100) |
| ChAdOx1/mRNA mixed schedule | | | | |
| ≥14 days after dose 1 | N/A | N/A | N/A | N/A |
| ≥7 days after dose 2 | 6 / 6,820 | 11,880 / 518,776 | 96 (92–98) | 100 (99–100) |
| **British Columbia** | | | | |
| BNT162b2 | | | | |
| ≥14 days after dose 1 | 426 / 5,169 | 77,202 / 599,609 | 39 (33–45) | 74 (71–77) |
| ≥7 days after dose 2 | 164 / 4,907 | 132,386 / 654,793 | 86 (84–88) | 97 (97–98) |
| mRNA-1273 | | | | |
| ≥14 days after dose 1 | 92 / 4,835 | 21,535 / 543,942 | 53 (42–62) | 82 (78–85) |
| ≥7 days after dose 2 | 34 / 4,777 | 33,990 / 556,397 | 89 (85–92) | 98 (97–99) |
| BNT162b2/mRNA-1273 | | | | |
| ≥14 days after dose 1 | N/A | N/A | N/A | N/A |
| ≥7 days after dose 2 | 217 / 4,960 | 180,825 / 703,232 | 87 (85–88) | 98 (97–98) |
| ChAdOx1 | | | | |
| ≥14 days after dose 1 | 30 / 4,773 | 9,390/ 531,797 | 65 (50–75) | 78 (68–84) |
| ≥7 days after dose 2 | 15 / 4,758 | 6,174 / 528,581 | 73 (56–84) | 94 (90–96) |
| ChAdOx1/mRNA mixed schedule | | | | |
| ≥14 days after dose 1 | N/A | N/A | N/A | N/A |
| ≥7 days after dose 2 | 0 / 4,743 | 1,658 / 524,065 | -[b] | -[b] |
| **Manitoba** | | | | |
| BNT162b2 | | | | |
| ≥14 days after dose 1 | 102 / 1,957 | 7,510 / 73,098 | 52 (41–61) | 84 (80–87) |
| ≥7 days after dose 2 | 30 / 1,885 | 17,380 / 82,968 | 94 (91–96) | 97 (96–98) |
| mRNA-1273 | | | | |
| ≥14 days after dose 1 | 32 / 1,887 | 3,176 / 68,764 | 64 (49–75) | 91 (87–94) |
| ≥7 days after dose 2 | 28 / 1,883 | 7,407 / 72,995 | 87 (81–91) | 97 (96–98) |

| | | | | |
|---|---|---|---|---|
| BNT162b2/mRNA-1273 | | | | |
| ≥14 days after dose 1 | N/A | N/A | N/A | N/A |
| ≥7 days after dose 2 | 6 / 1,861 | 5,736 / 71,324 | 96 (92–98) | 97 (93–99) |
| ChAdOx1 | | | | |
| ≥14 days after dose 1 | 36 / 1,891 | 1,928 / 67,516 | 34 (8–53) | 73 (61–81) |
| ≥7 days after dose 2 | Suppressed / 1,856 | 263 / 65,851 | 87 (4–98) | 93 (46–99) |
| ChAdOx1/mRNA mixed schedule | | | | |
| ≥14 days after dose 1 | N/A | N/A | N/A | N/A |
| ≥7 days after dose 2 | Suppressed / 1,858 | 2,917 / 68,505 | 96 (89–99) | 98 (93–99) |
| **Pooled** | | | | |
| BNT162b2 | | | | |
| ≥14 days after dose 1 | 1,947 / 29,937 | 252,323 / 1,654,286 | 57 (34–72) | 83 (77–87) |
| ≥7 days after dose 2 | 511 / 28,501 | 390,521 / 1,792,484 | 93 (89–96) | 98 (96–99) |
| mRNA-1273 | | | | |
| ≥14 days after dose 1 | 356 / 28,346 | 68,149 / 1,470,112 | 70 (51–82) | 88 (82–92) |
| ≥7 days after dose 2 | 132 / 28,122 | 109,282 / 1,511,245 | 93 (87–96) | 99 (97–99) |
| BNT162b2/mRNA-1273 | | | | |
| ≥14 days after dose 1 | N/A | N/A | N/A | N/A |
| ≥7 days after dose 2 | >230 / >13,642 | 212,987 / 1,614,950 | 95 (86–98) | 98 (97–99) |
| ChAdOx1 | | | | |
| ≥14 days after dose 1 | 230 / 35,034 | 30,437 / 1,939,296 | 60 (41–72) | 81 (74–87) |
| ≥7 days after dose 2 | >39 / 34,844 | 13,376 / 1,922,235 | 84 (75–89) | 97 (91–99) |
| ChAdOx1/mRNA mixed schedule | | | | |
| ≥14 days after dose 1 | N/A | N/A | N/A | N/A |
| ≥7 days after dose 2 | >12 / 28,005 | 25,170 / 1,427,133 | 97 (95–99) | 99 (98–100) |

N/A–not applicable

[a]Models adjusted for age group, sex, public health unit region, biweekly period of test, number of SARS-CoV-2 tests in the 3 months prior to 14 December 2020, presence of any comorbidity that increase the risk of severe COVID-19, receipt of influenza vaccination in current or prior influenza season, and neighbourhood income (material deprivation index in British Columbia), proportion of persons employed as non-health essential workers, persons per dwelling, and self-identified visible minority quintiles. Receipt of influenza vaccination was not adjusted for in British Columbia because of lack of data.

[b]VE estimated as 100% based on zero vaccinated test-positive cases.

**Supplemental Table S11: Province-specific and pooled vaccine effectiveness against severe outcomes of hospitalization or death ≥14 days after a first dose (for subjects who received only 1 dose) and ≥7 days after receiving a second dose by SARS-CoV-2 lineage in Ontario, Quebec, British Columbia, and Manitoba**

| | Test-positive vaccinated cases / total test-positive cases | Test-negative vaccinated controls, vaccinated / total test-negative controls | Unadjusted vaccine effectiveness (95% CI) | Adjusted[a] vaccine effectiveness (95% CI) |
|---|---|---|---|---|
| **Ontario** | | | | |
| Non-VOC | | | | |
| ≥14 days after dose 1 | 40 / 5,190 | 60,402 / 367,474 | 96 (95–97) | 71 (59–79) |
| ≥7 days after dose 2 | Suppressed | 148,539 / 455,611 | -[b] | 95 (87–98) |
| Alpha (B.1.1.7) | | | | |
| ≥14 days after dose 1 | 741 / 6,242 | 60,402 / 367,474 | 32 (26–37) | 83 (81–84) |
| ≥7 days after dose 2 | 33 / 5,534 | 148,539 / 455,611 | 99 (98–99) | 96 (95–97) |
| Beta/Gamma (B.1.3.51 or P.1) | | | | |
| ≥14 days after dose 1 | 28 / 194 | 60,402 / 367,474 | 14 (-28 to 43) | 83 (73–89) |
| ≥7 days after dose 2 | Suppressed | 148,539 / 455,611 | 98 (90–99) | 96 (82–99) |
| Beta (B.1.351) | | | | |
| ≥14 days after dose 1 | 15 / 153 | 60,402 / 367,474 | 45 (6–68) | 62 (30–79) |
| ≥7 days after dose 2 | Suppressed | 148,539 / 455,611 | 99 (89–100) | 95 (61–99) |
| Gamma (P.1) | | | | |
| ≥14 days after dose 1 | 36 / 338 | 60,402 / 367,474 | 39 (14–57) | 80 (71–87) |
| ≥7 days after dose 2 | Suppressed | 148,539 / 455,611 | 99 (95–100) | 94 (75–99) |
| Delta (B.1.617.2) | | | | |
| ≥14 days after dose 1 | 181 / 1,464 | 60,402 / 367,474 | 28 (16–39) | 85 (83–88) |
| ≥7 days after dose 2 | 147 / 1,430 | 148,539 / 455,611 | 76 (72–80) | 98 (98–99) |
| **Quebec** | | | | |
| Alpha (B.1.1.7)[c] | | | | |
| ≥14 days after dose 1 | 262 / 1,371 | 169,766 / 636,603 | 35 (26–43) | 86 (83–88) |
| ≥7 days after dose 2 | 8 / 1,117 | 217,206 / 684,043 | 98 (97–99) | 99 (98–100) |
| Delta (B.1.617.2)[d] | | | | |
| ≥14 days after dose 1 | 25 / 463 | 13,516 / 20,620 | 97 (96–98) | 97 (95–98) |
| ≥7 days after dose 2 | 94 / 532 | 179,476 / 186,580 | 99 (99–99) | -[b] |
| **British Columbia** | | | | |
| Non-VOC | | | | |
| ≥14 days after dose 1 | 38 / 489 | 108,127 / 630,534 | 59 (43–71) | 17 (-22 to 44) |
| ≥7 days after dose 2 | 2 / 453 | 197,161 / 719,568 | 99 (95–100) | 73 (-11 to 93) |
| Alpha (B.1.1.7) | | | | |
| ≥14 days after dose 1 | 122 / 784 | 108,127 / 630,534 | 11 (-8 to 27) | 79 (74–83) |
| ≥7 days after dose 2 | 4 / 666 | 197,161 / 249,401 | 98 (96–99) | 97 (93–99) |
| Beta/Gamma (B.1.3.51 or P.1) | | | | |
| ≥14 days after dose 1 | 12 / 202 | 108,127 / 630,534 | 69 (45–83) | 79 (60–89) |
| ≥7 days after dose 2 | 0 / 190 | 197,161 / 719,568 | -[b] | -[b] |
| Beta (B.1.351) | | | | |
| ≥14 days after dose 1 | 2 / 5 | 108,127 / 630,534 | -[e] | -[e] |
| ≥7 days after dose 2 | 0 / 3 | 197,161 / 719,568 | -[b] | -[b] |
| Gamma (P.1) | | | | |
| ≥14 days after dose 1 | 117 / 572 | 108,127 / 630,534 | -24 (-52 to -1) | 76 (69–81) |
| ≥7 days after dose 2 | 4 / 459 | 197,161 / 719,568 | 98 (94–99) | 98 (93–99) |
| Delta (B.1.617.2) | | | | |
| ≥14 days after dose 1 | 79 / 1,047 | 108,127 / 630,534 | 61 (50–69) | 81 (77–85) |
| ≥7 days after dose 2 | 161 / 1,129 | 197,161 / 719,568 | 56 (48–63) | 98 (97–98) |
| **Manitoba** | | | | |
| Non-VOC | | | | |
| ≥14 days after dose 1 | 36 / 939 | 12,623 / 78,211 | 79 (71–85) | 84 (76–89) |
| ≥7 days after dose 2 | 31 / 934 | 33,706 / 99,294 | 93 (90–95) | 94 (91–96) |
| Alpha (B.1.1.7) | | | | |
| ≥14 days after dose 1 | 84 / 713 | 12,623 / 78,211 | 31 (13–45) | 88 (85–91) |
| ≥7 days after dose 2 | 14 / 643 | 33,706 / 99,294 | 96 (93–97) | 97 (94–98) |
| Beta/Gamma (B.1.3.51 or P.1) | | | | |
| ≥14 days after dose 1 | Suppressed / 18 | 12,623 / 78,211 | -[e] | 74 (4–93) |
| ≥7 days after dose 2 | Suppressed / 13 | 33,706 / 99,294 | No cases | No cases |
| Beta (B.1.351) | | | | |
| ≥14 days after dose 1 | Suppressed / 8 | 12,623 / 78,211 | -[e] | -[e] |
| ≥7 days after dose 2 | Suppressed | 33,706 / 99,294 | No cases | No cases |

| | | | | |
|---|---|---|---|---|
| Gamma (P.1) | | | | |
| ≥14 days after dose 1 | Suppressed / 10 | 12,623 / 78,211 | -[e] | 73 (-79 to 96) |
| ≥7 days after dose 2 | Suppressed / 8 | 33,706 / 99,294 | No cases | No cases |
| Delta (B.1.617.2) | | | | |
| ≥14 days after dose 1 | 8 / 94 | 12,623 / 78,211 | 52 (0–77) | 86 (70–94) |
| ≥7 days after dose 2 | 9 / 95 | 33,706 / 99,294 | 80 (60–90) | 99 (97–99) |
| **Pooled** | | | | |
| Non-VOC | | | | |
| ≥14 days after dose 1 | 114 / 6618 | 181,152 / 1,076,219 | 85 (42–96) | 66 (14–86) |
| ≥7 days after dose 2 | >33 / >1387 | 230,867 / 818,862 | 99 (91–100) | 92 (83–97) |
| Alpha (B.1.1.7) | | | | |
| ≥14 days after dose 1 | 1209 / 9110 | 350,918 / 1,712,822 | 29 (19–37) | 84 (80–88) |
| ≥7 days after dose 2 | 59 / 7960 | 596,612 / 1,488,349 | 98 (96–99) | 98 (96–99) |
| Beta/Gamma (B.1.3.51 or P.1) | | | | |
| ≥14 days after dose 1 | >40 / 414 | 181,152 / 1,076,219 | -[e] | 81 (74–87) |
| ≥7 days after dose 2 | -[f] | -[f] | -[f] | -[f] |
| Beta (B.1.351) | | | | |
| ≥14 days after dose 1 | >17 / 166 | 181,152 / 1,076,219 | -[¶] | 61 (32–78) |
| ≥7 days after dose 2 | -[f] | -[f] | -[f] | -[f] |
| Gamma (P.1) | | | | |
| ≥14 days after dose 1 | >153 / 920 | 181,152 / 1,076,219 | 8 (-65 to 48) | 77 (72–81) |
| ≥7 days after dose 2 | >4 / >467 | 230,867 / 818,862 | 98 (96–99) | 97 (93–99) |
| Delta (B.1.617.2) | | | | |
| ≥14 days after dose 1 | 293 / 3068 | 194,668 / 1,096,839 | 75 (-5 to 94) | 89 (77–95) |
| ≥7 days after dose 2 | 411 / 3186 | 558,882 / 1,461,053 | 88 (34–98) | 99 (97–99) |

[a]Models adjusted for age group, sex, public health unit region, biweekly period of test, number of SARS-CoV-2 tests in the 3 months prior to 14 December 2020, presence of any comorbidity that increase the risk of severe COVID-19, receipt of influenza vaccination in current or prior influenza season, and neighbourhood income (material deprivation index in British Columbia), proportion of persons employed as non-health essential workers, persons per dwelling, and self-identified visible minority quintiles. Receipt of influenza vaccination was not adjusted for in British Columbia because of lack of data.

[b]VE estimated as 100% based on zero vaccinated test-positive cases.

[c]Analysis period restricted from 28 December 2020 to 30 September 2021.

[d]Analysis period restricted from 26 July 2021 to 30 September 2021.

[e]VE not reported due to extremely imprecise 95% confidence intervals.

[f]Not applicable as VE estimate from only one province available.

**Supplemental Table S12: Province-specific and pooled vaccine effectiveness against severe outcomes of hospitalization or death by varying dosing intervals among subjects who received 2 doses of an mRNA in Ontario, Quebec, British Columbia, and Manitoba**

| Interval between doses | Test-positive vaccinated cases / total test-positive cases | Test-negative vaccinated controls, vaccinated / total test-negative controls | Unadjusted vaccine effectiveness (95% CI) | Adjusted[a] vaccine effectiveness (95% CI) |
|---|---|---|---|---|
| **Ontario** | | | | |
| **0–6 days after dose 2** | | | | |
| 21–34 days | 14 / 14,592 | 1,323 / 308,395 | 78 (62–87) | 85 (74–91) |
| 35–55 days | Suppressed | 1,654 / 308,726 | 94 (85–97) | 93 (84–97) |
| 56–83 days | 6 / 14,584 | 1,454 / 308,526 | 91 (81–96) | 96 (90–98) |
| ≥84 days | 6 / 14,584 | 873 / 307,945 | 86 (68–94) | 92 (82–96) |
| **7–55 days after dose 2** | | | | |
| 21–34 days | 44 / 14,622 | 7,892 / 314,964 | 88 (84–91) | 94 (92–96) |
| 35–55 days | 11 / 14,589 | 20,983 / 328,055 | 99 (98–99) | 99 (98–99) |
| 56–83 days | 32 / 14,610 | 20,457 / 327,529 | 97 (95–98) | 98 (97–99) |
| ≥84 days | 17 / 14,595 | 7,575 / 314,647 | 95 (92–97) | 98 (97–99) |
| **56–111 days after dose 2** | | | | |
| 21–34 days | 22 / 14,600 | 7,922 / 314,994 | 94 (91–96) | 98 (96–98) |
| 35–55 days | 11 / 14,589 | 25,707 / 332,779 | 99 (98–100) | 99 (98–99) |
| 56–83 days | 48 / 14,626 | 24,173 / 331,245 | 96 (94–97) | 99 (98–99) |
| ≥84 days | 21 / 14,599 | 8,987 / 316,059 | 95 (92–97) | 98 (98–99) |
| **≥112 days after dose 2** | | | | |
| 21–34 days | 36 / 14,614 | 6,599 / 313,671 | 89 (84–92) | 96 (95–97) |
| 35–55 days | 8 / 14,586 | 3,709 / 310,781 | 95 (91–98) | 97 (93–98) |
| 56–83 days | Suppressed | 1,424 / 308,496 | 94 (84–98) | 97 (91–99) |
| ≥84 days | Suppressed | 830 / 307,902 | 97 (82–100) | 98 (85–100) |
| **Quebec** | | | | |
| **0–6 days after dose 2** | | | | |
| 21–34 days | 1 / 6,815 | 930 / 507,826 | 92 (43–99) | 99 (90–100) |
| 35–55 days | 1 / 6,815 | 1,483 / 508,379 | 95 (64–99) | 97 (78–100) |
| 56–83 days | 4 / 6,818 | 5,290 / 512,186 | 94 (85–98) | 94 (85–98) |
| ≥84 days | 2 / 6,816 | 3,990 / 510,886 | 96 (85–99) | 99 (95–100) |
| **7–55 days after dose 2** | | | | |
| 21–34 days | 9 / 6,823 | 4,020 / 510,916 | 83 (68–91) | 97 (95–99) |
| 35–55 days | 2 / 6,816 | 12,446 / 519,342 | 99 (95–100) | 100 (99–100) |
| 56–83 days | 19 / 6,833 | 56,807 / 563,703 | 98 (96–98) | 99 (99–100) |
| ≥84 days | 21 / 6,835 | 27,126 / 534,022 | 94 (91–96) | 99 (99–100) |
| **56–111 days after dose 2** | | | | |
| 21–34 days | 7 / 6,821 | 1,989 / 508,885 | 74 (45–88) | 97 (93–98) |
| 35–55 days | 8 / 6,822 | 11,875 / 518,771 | 95 (90–97) | 99 (99–100) |
| 56–83 days | 24 / 6,838 | 52,981 / 559,877 | 97 (95–98) | 100 (100–100) |
| ≥84 days | 48 / 6,862 | 21,225 / 528,121 | 83 (78–87) | 100 (99–100) |
| **≥112 days after dose 2** | | | | |
| 21–34 days | 2 / 6,816 | 877 / 507,773 | 83 (32–96) | 99 (96–100) |
| 35–55 days | 0 / 6,814 | 531 / 507,427 | Not estimable | Not estimable |
| 56–83 days | 7 / 6,821 | 922 / 507,818 | 44 (-19 to 73) | 98 (97–99) |
| ≥84 days | 2 / 6,816 | 7,543 / 514,439 | 98 (92–100) | 100 (99–100) |
| **British Columbia** | | | | |
| **0–6 days after dose 2** | | | | |
| 21–34 days | 0 / 4,743 | 471 / 522,878 | -[b] | -[b] |
| 35–55 days | 3 / 4,746 | 2,291 / 524,698 | 86 (55–95) | 90 (69–97) |
| 56–83 days | 8 / 4,751 | 3,784 / 526,191 | 77 (53–88) | 90 (79–95) |
| ≥84 days | 7 / 4,750 | 1,527 / 523,934 | 50 (-6 to 76) | 85 (67–93) |
| **7–55 days after dose 2** | | | | |
| 21–34 days | 6 / 4,749 | 2,045 / 524,452 | 68 (28–86) | 91 (79–96) |
| 35–55 days | 15 / 4,758 | 20,985 / 543,392 | 92 (87–95) | 97 (94–98) |

| | | | | |
|---|---|---|---|---|
| 56–83 days | 16 / 4,759 | 49,941 / 572,348 | 96 (94–98) | 99 (98–99) |
| ≥84 days | 27 / 4,770 | 11,262 / 533,669 | 74 (61–82) | 95 (93–97) |
| **56–111 days after dose 2** | | | | |
| 21–34 days | 1 / 4,744 | 1,242 / 523,649 | 91 (37–99) | 97 (80–100) |
| 35–55 days | 3 / 4,746 | 16,993 / 539,400 | 98 (94–99) | 100 (98–100) |
| 56–83 days | 62 / 4,805 | 48,252 / 570,659 | 86 (82–89) | 98 (98–99) |
| ≥84 days | 58 / 4,801 | 16,452 / 538,859 | 61 (50–70) | 96 (95–97) |
| **≥112 days after dose 2** | | | | |
| 21–34 days | 5 / 4,748 | 1,145 / 523,552 | 52 (-15 to 80) | 91 (78–96) |
| 35–55 days | 16 / 4,759 | 7,860 / 530,267 | 78 (63–86) | 96 (94–98) |
| 56–83 days | 6 / 4,749 | 1,468 / 523,875 | 55 (0–80) | 96 (92–98) |
| ≥84 days | 2 / 4,745 | 3,053 / 525,460 | 93 (71–98) | 99 (95–100) |
| **Manitoba** | | | | |
| **0–6 days after dose 2** | | | | |
| 21–34 days | Suppressed / 1,855 | 454 / 66,042 | No cases | No cases |
| 35–55 days | Suppressed / 1,860 | 436 / 66,024 | 59 (2–83) | 87 (68–95) |
| 56–83 days | Suppressed / 1,856 | 285 / 65,873 | 88 (12–98) | 97 (79–100) |
| ≥84 days | Suppressed / 1,856 | 45 / 65,633 | -[c] | 84 (-17 to 98) |
| **7–55 days after dose 2** | | | | |
| 21–34 days | 14 / 1,869 | 3,345 / 68,933 | 85 (75–91) | 93 (88–96) |
| 35–55 days | 12 / 1,867 | 4,140 / 69,728 | 90 (82–94) | 94 (90–97) |
| 56–83 days | 8 / 1,863 | 3,054 / 68,642 | 91 (81–95) | 97 (93–98) |
| ≥84 days | Suppressed / 1,858 | 343 / 65,931 | 69 (4–90) | 94 (81–98) |
| **56–111 days after dose 2** | | | | |
| 21–34 days | Suppressed / 1,860 | 4,452 / 70,040 | 96 (90–98) | 98 (96–99) |
| 35–55 days | 10 / 1,865 | 6,500 / 72,088 | 95 (90–97) | 97 (95–99) |
| 56–83 days | Suppressed / 1,859 | 4,099 / 69,687 | 97 (91–99) | 99 (97–100) |
| ≥84 days | Suppressed / 1,855 | 273 / 65,861 | No cases | No cases |
| **≥112 days after dose 2** | | | | |
| 21–34 days | Suppressed / 1,860 | 3,226 / 68,814 | 95 (87–98) | 98 (95–99) |
| 35–55 days | Suppressed / 1,858 | 692 / 66,280 | 85 (52–95) | 99 (95–100) |
| 56–83 days | Suppressed / 1,855 | 132 / 65,720 | No cases | No cases |
| ≥84 days | Suppressed / 1,855 | 16 / 65,604 | No cases | No cases |
| **Pooled** | | | | |
| **0–6 days after dose 2** | | | | |
| 21–34 days | >15 / 28,005 | 3,178 / 1,405,141 | 79 (65–88) | 94 (47–99) |
| 35–55 days | >4 / >13,421 | 5,864 / 1,407,827 | 87 (66–95) | 91 (85–95) |
| 56–83 days | >18 / 28,009 | 10,813 / 1,412,776 | 89 (77–95) | 94 (89–96) |
| ≥84 days | >15 / 28,006 | 6,435 / 1,408,398 | 79 (28–94) | 93 (79–97) |
| **7–55 days after dose 2** | | | | |
| 21–34 days | 73 / 28,063 | 17,302 / 1,419,265 | 84 (77–89) | 94 (91–97) |
| 35–55 days | 40 / 28,030 | 58,554 / 1,460,517 | 97 (89–99) | 98 (94–99) |
| 56–83 days | 75 / 28,065 | 130,259 / 1,532,222 | 96 (94–98) | 99 (97–99) |
| ≥84 days | >65 / 28,058 | 46,306 / 1,448,269 | 88 (70–96) | 98 (94–99) |
| **56–111 days after dose 2** | | | | |
| 21–34 days | >30 / 28,025 | 15,605 / 1,417,568 | 91 (79–97) | 98 (97–98) |
| 35–55 days | 32 / 28,022 | 61,075 / 1,463,038 | 97 (94–99) | 99 (98–100) |
| 56–83 days | >134 / 28,128 | 129,505 / 1,531,468 | 95 (89–97) | 99 (98–100) |
| ≥84 days | >127 / 28,117 | 46,937 / 1,448,900 | 85 (52–95) | 99 (95–100) |
| **≥112 days after dose 2** | | | | |
| 21–34 days | >43 / 28,038 | 11,847 / 1,413,810 | 85 (64–94) | 97 (93–99) |
| 35–55 days | >24 / 28,017 | 12,792 / 1,414,755 | 88 (68–96) | 97 (95–98) |
| 56–83 days | >13 / >13,425 | 3,946 / 1,405,909 | 75 (-1 to 94) | 97 (96–99) |
| ≥84 days | >4 / >13,416 | 11,442 / 1,413,405 | 97 (91–99) | 99 (97–100) |

[a]Models adjusted for age group, sex, public health unit region, biweekly period of test, number of SARS-CoV-2 tests in the 3 months prior to 14 December 2020, presence of any comorbidity that increase the risk of severe COVID-19, receipt of influenza vaccination in current or prior influenza season, and neighbourhood income (material deprivation index in British Columbia), proportion of persons employed as non-health essential workers, persons per dwelling, and self-identified visible minority quintiles. Receipt of influenza vaccination was not adjusted for in British Columbia because of lack of data.

[b]VE estimated as 100% based on zero vaccinated test-positive cases.

[c]VE not reported due to extremely imprecise 95% confidence intervals.

**Supplemental Table S13: Test of heterogeneity for all pooled primary analyses**

| Outcome/subgroup | Heterogeneity, pooled unadjusted model | Heterogeneity, pooled adjusted model |
|---|---|---|
| **Hospitalization** | | |
| ≥14 days after dose 1 | $I^2$ = 99%, $\tau^2$ = 0.1655, p < 0.01 | $I^2$ = 96%, $\tau^2$ = 0.0660, p < 0.01 |
| ≥7 days after dose 2 | $I^2$ = 98%, $\tau^2$ = 0.2413, p < 0.01 | $I^2$ = 99%, $\tau^2$ = 0.5800, p < 0.01 |
| **Death** | | |
| ≥14 days after dose 1 | $I^2$ = 96%, $\tau^2$ = 0.2252, p < 0.01 | $I^2$ = 92%, $\tau^2$ = 0.2379, p < 0.01 |
| ≥7 days after dose 2 | $I^2$ = 92%, $\tau^2$ = 0.3546, p <0.01 | $I^2$ = 92%, $\tau^2$ = 0.7433, p < 0.01 |
| **Severe outcomes (hospitalization or death)** | | |
| **mRNA vaccines** | | |
| **Dose 1** | | |
| 0–13 days | $I^2$ = 98%, $\tau^2$ = 0.1310, p < 0.01 | $I^2$ = 94%, $\tau^2$ = 0.0772, p < 0.01 |
| 14–27 days | $I^2$ = 96%, $\tau^2$ = 0.1171, p < 0.01 | $I^2$ = 88%, $\tau^2$ = 0.0658, p < 0.01 |
| 28–55 days | $I^2$ = 97%, $\tau^2$ = 0.2124, p < 0.01 | $I^2$ = 90%, $\tau^2$ = 0.0669, p < 0.01 |
| 56–83 days | $I^2$ = 95%, $\tau^2$ = 0.3143, p < 0.01 | $I^2$ = 89%, $\tau^2$ = 0.1038, p < 0.01 |
| ≥84days | $I^2$ = 97%, $\tau^2$ = 0.6669, p < 0.01 | $I^2$ = 97%, $\tau^2$ = 0.6302, p < 0.01 |
| **Dose 2** | | |
| 0–6 days | $I^2$ = 81%, $\tau^2$ = 0.3687, p < 0.01 | $I^2$ = 76%, $\tau^2$ = 0.2988, p < 0.01 |
| 7–55 days | $I^2$ = 95%, $\tau^2$ = 0.3107, p < 0.01 | $I^2$ = 97%, $\tau^2$ = 0.6757, p < 0.01 |
| 56–111 days | $I^2$ = 98%, $\tau^2$ = 0.5009, p < 0.01 | $I^2$ = 98%, $\tau^2$ = 0.6084, p < 0.01 |
| ≥112 days | $I^2$ = 88%, $\tau^2$ = 0.2864, p < 0.01 | $I^2$ = 91%, $\tau^2$ = 0.6390, p < 0.01 |
| **ChAdOx1 vaccine** | | |
| **Dose 1** | | |
| 0-13 days | $I^2$ = 71%, $\tau^2$ = 0.0603, p = 0.02 | $I^2$ = 54%, $\tau^2$ = 0.0325, p = 0.09 |
| 14-27 days | $I^2$ = 60%, $\tau^2$ = 0.0651, p = 0.06 | $I^2$ = 31%, $\tau^2$ = 0.0207, p = 0.22 |
| 28-55 days | $I^2$ = 77%, $\tau^2$ = 0.2198, p < 0.01 | $I^2$ = 80%, $\tau^2$ = 0.2015, p < 0.01 |
| ≥56 days | $I^2$ = 88%, $\tau^2$ = 0.7048, p < 0.01 | $I^2$ = 83%, $\tau^2$ = 0.4557, p < 0.01 |
| **Dose 2** | | |
| 0-6 days | $I^2$ = 0%, $\tau^2$ = 0, p = 0.88 | $I^2$ = 0%, $\tau^2$ = 0, p = 0.75 |
| 7-55 days | $I^2$ = 0%, $\tau^2$ = 0, p = 0.57 | $I^2$ = 0%, $\tau^2$ = 0, p = 0.60 |
| ≥56 days | $I^2$ = 33%, $\tau^2$ = 0.0804, p = 0.21 | $I^2$ = 88%, $\tau^2$ = 0.8968, p < 0.01 |
| **Age group** | | |
| 18–59 years | | |
| ≥14 days after dose 1 | $I^2$ = 97%, $\tau^2$ = 0.4934, p < 0.01 | $I^2$ = 96%, $\tau^2$ = 0.2888, p < 0.01 |
| ≥7 days after dose 2 | $I^2$ = 92%, $\tau^2$ = 0.2064, p < 0.01 | $I^2$ = 94%, $\tau^2$ = 0.4852, p < 0.01 |
| 60–69 years | | |
| ≥14 days after dose 1 | $I^2$ = 94%, $\tau^2$ = 0.1928, p < 0.01 | $I^2$ = 88%, $\tau^2$ = 0.1008, p < 0.01 |
| ≥7 days after dose 2 | $I^2$ = 93%, $\tau^2$ = 0.3422, p < 0.01 | $I^2$ = 95%, $\tau^2$ = 1.0229, p < 0.01 |
| 70–79 years | | |
| ≥14 days after dose 1 | $I^2$ = 94%, $\tau^2$ = 0.1271, p < 0.01 | $I^2$ = 85%, $\tau^2$ = 0.1184, p < 0.01 |
| ≥7 days after dose 2 | $I^2$ = 94%, $\tau^2$ = 0.3061, p < 0.01 | $I^2$ = 94%, $\tau^2$ = 0.7209, p < 0.01 |
| ≥80 years | | |
| ≥14 days after dose 1 | $I^2$ = 98%, $\tau^2$ = 0.3697, p < 0.01 | $I^2$ = 89%, $\tau^2$ = 0.1080, p < 0.01 |
| ≥7 days after dose 2 | $I^2$ = 96%, $\tau^2$ = 0.3839, p < 0.01 | $I^2$ = 89%, $\tau^2$ = 0.3947, p < 0.01 |
| **Sex** | | |
| Females | | |
| ≥14 days after dose 1 | $I^2$ = 98%, $\tau^2$ = 0.1888, p < 0.01 | $I^2$ = 94%, $\tau^2$ = 0.1044, p < 0.01 |
| ≥7 days after dose 2 | $I^2$ = 95%, $\tau^2$ = 0.1877, p < 0.01 | $I^2$ = 97%, $\tau^2$ = 0.5889, p < 0.01 |
| Males | | |
| ≥14 days after dose 1 | $I^2$ = 98%, $\tau^2$ = 0.1878, p < 0.01 | $I^2$ = 93%, $\tau^2$ = 0.0698, p < 0.01 |
| ≥7 days after dose 2 | $I^2$ = 98%, $\tau^2$ = 0.2939, p < 0.01 | $I^2$ = 98%, $\tau^2$ = 0.7051, p < 0.01 |
| **Presence of any comorbidity** | | |

| | | |
|---|---|---|
| Yes | | |
| ≥14 days after dose 1 | $I^2$ = 99%, $\tau^2$ = 0.1757, p < 0.01 | $I^2$ = 96%, $\tau^2$ = 0.0881, p < 0.01 |
| ≥7 days after dose 2 | $I^2$ = 98%, $\tau^2$ = 0.2575, p < 0.01 | $I^2$ = 98%, $\tau^2$ = 0.5554, p < 0.01 |
| No | | |
| ≥14 days after dose 1 | $I^2$ = 94%, $\tau^2$ = 0.2181, p < 0.01 | $I^2$ = 90%, $\tau^2$ = 0.1364, p < 0.01 |
| ≥7 days after dose 2 | $I^2$ = 83%, $\tau^2$ = 0.1745, p < 0.01 | $I^2$ = 92%, $\tau^2$ = 0.4905, p < 0.01 |
| **Vaccine product** | | |
| BNT162b2 | | |
| ≥14 days after dose 1 | $I^2$ = 99%, $\tau^2$ = 0.1901, p < 0.01 | $I^2$ = 96%, $\tau^2$ = 0.0867, p < 0.01 |
| ≥7 days after dose 2 | $I^2$ = 97%, $\tau^2$ = 0.2179, p < 0.01 | $I^2$ = 98%, $\tau^2$ = 0.6920, p < 0.01 |
| mRNA-1273 | | |
| ≥14 days after dose 1 | $I^2$ = 95%, $\tau^2$ = 0.2477, p < 0.01 | $I^2$ = 91%, $\tau^2$ = 0.1568, p < 0.01 |
| ≥7 days after dose 2 | $I^2$ = 93%, $\tau^2$ = 0.3952, p < 0.01 | $I^2$ = 95%, $\tau^2$ = 0.6847, p < 0.01 |
| BNT162b2/mRNA-1273 | | |
| ≥14 days after dose 1 | - | - |
| ≥7 days after dose 2 | $I^2$ = 96%, $\tau^2$ = 1.1175, p <0.01 | $I^2$ = 80%, $\tau^2$ = 0.2465, p < 0.01 |
| ChAdOx1 | | |
| ≥14 days after dose 1 | $I^2$ = 84%, $\tau^2$ = 0.1230, p < 0.01 | $I^2$ = 80%, $\tau^2$ = 0.0833, p < 0.01 |
| ≥7 days after dose 2 | $I^2$ = 45%, $\tau^2$ =0.0934, p = 0.14 | $I^2$ = 90%, $\tau^2$ = 0.9094, p < 0.01 |
| ChAdOx1/mRNA mixed schedule | | |
| ≥14 days after dose 1 | - | - |
| ≥7 days after dose 2 | $I^2$ = 39%, $\tau^2$ = 0.1377, p = 0.19 | $I^2$ = 62%, $\tau^2$ = 0.3639, p = 0.07 |
| **SARS-CoV-2 lineage** | | |
| Non-VOC | | |
| ≥14 days after dose 1 | $I^2$ = 98%, $\tau^2$ = 1.4166, p < 0.01 | $I^2$ = 95%, $\tau^2$ = 0.6307, p < 0.01 |
| ≥7 days after dose 2 | $I^2$ = 97%, τ2 = 3.0728, p < 0.01 | $I^2$ = 58%, $\tau^2$ = 0.3056, p = 0.09 |
| Alpha (B.1.1.7) | | |
| ≥14 days after dose 1 | $I^2$ = 61%, τ2 = 0.0104, p = 0.05 | $I^2$ = 82%, $\tau^2$ = 0.0491, p < 0.01 |
| ≥7 days after dose 2 | $I^2$ = 81%, τ2 = 0.2803, p < 0.01 | $I^2$ = 70%, $\tau^2$ = 0.2549, p = 0.02 |
| Beta/Gamma (B.1.3.51 or P.1) | | |
| ≥14 days after dose 1 | $I^2$ = 85%, τ2 = 0.6831, p < 0.01 | $I^2$ = 0%, $\tau^2$ = 0, p = 0.73 |
| ≥7 days after dose 2 | - | - |
| Beta (B.1.351) | | |
| ≥14 days after dose 1 | $I^2$ = 73%, τ2 = 0.8952, p = 0.03 | $I^2$ = 0%, $\tau^2$ = 0, p = 0.93 |
| ≥7 days after dose 2 | - | - |
| Gamma (P.1) | | |
| ≥14 days after dose 1 | $I^2$ = 85%, τ2 = 0.1785, p < 0.01 | $I^2$ = 0%, $\tau^2$ = 0, p = 0.67 |
| ≥7 days after dose 2 | $I^2$ = 0%, $\tau^2$ = 0, p = 0.53 | $I^2$ = 0%, $\tau^2$ = 0, p = 0.32 |
| Delta (B.1.617.2) | | |
| ≥14 days after dose 1 | $I^2$ = 99%, $\tau^2$ = 2.0504, p < 0.01 | $I^2$ = 94%, $\tau^2$ = 0.5761, p < 0.01 |
| ≥7 days after dose 2 | $I^2$ = 100%, $\tau^2$ = 3.1546, p < 0.01 | $I^2$ = 98%, $\tau^2$ = 0.6445, p < 0.01 |
| **Interval between doses** | | |
| **0–6 days after dose 2** | | |
| 21–34 days | $I^2$ = 2%, $\tau^2$ = 0.0093, p = 0.31 | $I^2$ = 81%, $\tau^2$ = 2.1665, p = 0.02 |
| 35–55 days | $I^2$ = 70%, $\tau^2$ = 0.6279, p = 0.02 | $I^2$ = 0%, $\tau^2$ = 0, p = 0.51 |
| 56–83 days | $I^2$ = 55%, $\tau^2$ = 0.2817, p = 0.08 | $I^2$ = 19%, $\tau^2$ = 0.0936, p = 0.29 |
| ≥84 days | $I^2$ = 79%, $\tau^2$ = 1.2437, p < 0.01 | $I^2$ = 70%, $\tau^2$ = 0.8153, p = 0.02 |
| **7–55 days after dose 2** | | |
| 21–34 days | $I^2$ = 51%, $\tau^2$ = 0.0677, p = 0.11 | $I^2$ = 62%, $\tau^2$ = 0.1501, p = 0.05 |
| 35–55 days | $I^2$ = 92%, $\tau^2$ = 1.2793, p < 0.01 | $I^2$ = 86%, $\tau^2$ = 1.1650, p < 0.01 |
| 56–83 days | $I^2$ = 71%, $\tau^2$ = 0.1895, p = 0.02 | $I^2$ = 87%, $\tau^2$ = 0.4281, p < 0.01 |
| ≥84 days | $I^2$ = 94%, $\tau^2$ = 0.8531, p < 0.01 | $I^2$ = 94%, $\tau^2$ = 0.9583, p < 0.01 |
| **56–111 days after dose 2** | | |

| | | |
|---|---|---|
| 21–34 days | $I^2$ = 80%, $\tau^2$ = 0.6144, p < 0.01 | $I^2$ = 0%, $\tau^2$ = 0, p = 0.78 |
| 35–55 days | $I^2$ = 87%, $\tau^2$ = 0.6703, p < 0.01 | $I^2$ = 74%, $\tau^2$ = 0.3732, p < 0.01 |
| 56–83 days | $I^2$ = 95%, $\tau^2$ = 0.4620, p < 0.01 | $I^2$ = 96%, $\tau^2$ = 0.8032, p < 0.01 |
| ≥84 days | $I^2$ = 97%, $\tau^2$ = 1.0410, p < 0.01 | $I^2$ = 98%, $\tau^2$ = 1.2924, p < 0.01 |
| **≥112 days after dose 2** | | |
| 21–34 days | $I^2$ = 77%, $\tau^2$ = 0.6388, p < 0.01 | $I^2$ = 69%, $\tau^2$ = 0.4948, p = 0.02 |
| 35–55 days | $I^2$ = 86%, $\tau^2$ = 0.6369, p < 0.01 | $I^2$ = 18%, $\tau^2$ < 0.0001, p = 0.30 |
| 56–83 days | $I^2$ = 87%, τ2 = 1.3047, p < 0.01 | $I^2$ = 15%, $\tau^2$ = 0.0482, p = 0.31 |
| ≥84 days | $I^2$ = 0%, $\tau^2$ = 0.0606, p = 0.39 | $I^2$ = 61%, $\tau^2$ = 0.9336, p = 0.08 |

**Supplemental Table S14: Pooled vaccine effectiveness from the sensitivity analyses**

| Outcome/subgroup | Test-positive vaccinated cases / total test-positive cases | Test-negative vaccinated controls, vaccinated / total test-negative controls | Unadjusted model | | Adjusted model[a] | |
|---|---|---|---|---|---|---|
| | | | Vaccine effectiveness (95% CI) | Heterogeneity | Vaccine effectiveness (95% CI) | Heterogeneity |
| **Hospitalization** | | | | | | |
| ≥14 days after dose 1 | 2,799 / 33,924 | 351,068 / 1,753,334 | 59 (73–40) | $I^2$ = 99%, $\tau^2$ = 0.1620, p < 0.01 | 83 (87–78) | $I^2$ = 96%, $\tau^2$ = 0.0658, p < 0.01 |
| ≥7 days after dose 2 | 753 / 31,878 | 588,166 / 1,990,432 | 93 (96–88) | $I^2$ = 99%, $\tau^2$ = 0.3050, p < 0.01 | 98 (99–96) | $I^2$ = 99%, $\tau^2$ = 0.6086, p < 0.01 |
| **Death** | | | | | | |
| ≥14 days after dose 1 | 567 / 6,064 | 352,518 / 1,771,529 | 53 (71–24) | $I^2$ = 96%, $\tau^2$ = 0.2276, p < 0.01 | 83 (90–71) | $I^2$ = 93%, $\tau^2$ = 0.2525, p < 0.01 |
| ≥7 days after dose 2 | 144 / 5,641 | 588,447 / 2,007,458 | 93 (96–87) | $I^2$ = 92%, $\tau^2$ = 0.3302, p < 0.01 | 98 (99–95) | $I^2$ = 94%, $\tau^2$ = 0.8657, p < 0.01 |
| **Severe outcomes (hospitalization or death)** | | | | | | |
| **mRNA vaccines** | | | | | | |
| **Dose 1** | | | | | | |
| 0–13 days | 2,202 / 34,738 | 86,729 / 1,488,361 | 2 (31 to -40) | $I^2$ = 98%, $\tau^2$ = 0.1242, p < 0.01 | 44 (57–26) | $I^2$ = 94%, $\tau^2$ = 0.0703, p < 0.01 |
| 14–27 days | 1,054 / 33,590 | 88,982 / 1,490,614 | 53 (66–34) | $I^2$ = 96%, $\tau^2$ = 0.1092, p < 0.01 | 78 (83–71) | $I^2$ = 88%, $\tau^2$ = 0.0617, p < 0.01 |
| 28–55 days | 939 / 33,475 | 137,105 / 1,538,737 | 67 (79–47) | $I^2$ = 97%, $\tau^2$ = 0.2063, p < 0.01 | 86 (89–82) | $I^2$ = 90%, $\tau^2$ = 0.0670, p < 0.01 |
| 56–83 days | 432 / 32,968 | 65,346 / 1,466,978 | 61 (77–31) | $I^2$ = 95%, $\tau^2$ = 0.3111, p < 0.01 | 87 (90–82) | $I^2$ = 88%, $\tau^2$ = 0.0919, p < 0.01 |
| ≥84days | 198 / 32,739 | 29,104 / 1,430,736 | 57 (82–1) | $I^2$ = 97%, $\tau^2$ = 0.6837, p < 0.01 | 86 (94–69) | $I^2$ = 97%, $\tau^2$ = 0.6443, p < 0.01 |
| **Dose 2** | | | | | | |
| 0–6 days | 72 / 32,608 | 26,332 / 1,427,964 | 87 (93–74) | $I^2$ = 82%, $\tau^2$ = 0.3715, p < 0.01 | 93 (96–88) | $I^2$ = 77%, $\tau^2$ = 0.3116, p < 0.01 |
| 7–55 days | 283 / 32,819 | 252,683 / 1,654,315 | 94 (97–89) | $I^2$ = 95%, $\tau^2$ = 0.3258, p < 0.01 | 98 (99–95) | $I^2$ = 97%, $\tau^2$ = 0.7018, p < 0.01 |
| 56–111 days | 346 / 32,882 | 253,388 / 1,655,020 | 94 (97–87) | $I^2$ = 98%, $\tau^2$ = 0.5800, p < 0.01 | 99 (99–97) | $I^2$ = 98%, $\tau^2$ = 0.6339, p < 0.01 |
| ≥112 days | 102 / 32,638 | 40,541 / 1,442,173 | 90 (95–82) | $I^2$ = 91%, $\tau^2$ = 0.3372, p < 0.01 | 98 (99–95) | $I^2$ = 92%, $\tau^2$ = 0.7196, p < 0.01 |
| **ChAdOx1 vaccine** | | | | | | |
| **Dose 1** | | | | | | |
| 0-13 days | 233 / 32,769 | 10,441 / 1,412,073 | 8 (36 to -31) | $I^2$ = 83%, $\tau^2$ = 0.1006, p < 0.01 | 35 (45–23) | $I^2$ = 15%, $\tau^2$ = 0.0075, p = 0.32 |
| 14-27 days | 134 / 32,670 | 9,460 / 1,411,092 | 43 (59–20) | $I^2$ = 62%, $\tau^2$ = 0.0663, p = 0.05 | 69 (76–60) | $I^2$ = 35%, $\tau^2$ = 0.0231, p = 0.20 |
| 28-55 days | 89 / 32,625 | 13,182 / 1,414,814 | 68 (81–47) | $I^2$ = 77%, $\tau^2$ = 0.2165, p < 0.01 | 86 (91–77) | $I^2$ = 80%, $\tau^2$ = 0.1913, p < 0.01 |
| ≥56 days | 45 / 32,581 | 7,840 / 1,409,472 | 69 (87–23) | $I^2$ = 89%, $\tau^2$ = 0.7094, p < 0.01 | 88 (94–75) | $I^2$ = 83%, $\tau^2$ = 0.4465, p < 0.01 |
| **Dose 2** | | | | | | |
| 0-6 days | >1 / >13,413 | 511 / 1,402,143 | 56 (86 to -34) | $I^2$ = 0%, $\tau^2$ = 0, p = 0.60 | 82 (94–44) | $I^2$ = 0%, $\tau^2$ = 0, p = 0.92 |
| 7-55 days | >3 / >13,415 | 5,543 / 1,407,175 | 87 (93–76) | $I^2$ = 0%, $\tau^2$ = 0, p = 0.44 | 93 (96–88) | $I^2$ = 0%, $\tau^2$ = 0, p = 0.65 |
| ≥56 days | >38 / 32,575 | 10,957 / 1,412,589 | 81 (89–67) | $I^2$ = 64%, $\tau^2$ = 0.1949, p = 0.04 | 97 (99–91) | $I^2$ = 89%, $\tau^2$ = 0.9065, p < 0.01 |
| **Subject characteristics** | | | | | | |
| **Age group** | | | | | | |
| 18–59 years | | | | | | |
| ≥14 days after dose 1 | 528 / 16,089 | 240,726 / 1,390,441 | 80 (90–60) | $I^2$ = 97%, $\tau^2$ = 0.4977, p < 0.01 | 89 (93–80) | $I^2$ = 96%, $\tau^2$ = 0.2915, p < 0.01 |
| ≥7 days after dose 2 | 190 / 15,751 | 432,893 / 1,582,608 | 96 (98–94) | $I^2$ = 94%, $\tau^2$ = 0.2570, p < 0.01 | 99 (99–97) | $I^2$ = 94%, $\tau^2$ = 0.4949, p < 0.01 |
| 60–69 years | | | | | | |
| ≥14 days after dose 1 | 541 / 6,916 | 49,857 / 194,472 | 72 (82–57) | $I^2$ = 93%, $\tau^2$ = 0.1889, p < 0.01 | 86 (90–80) | $I^2$ = 87%, $\tau^2$ = 0.0936, p < 0.01 |

| | | | | | | |
|---|---|---|---|---|---|---|
| ≥7 days after dose 2 | 158 / 6,533 | 80,525 / 225,140 | 94 (97–89) | $I^2$ = 95%, $\tau^2$ = 0.4230, p < 0.01 | 98 (99–96) | $I^2$ = 96%, $\tau^2$ = 1.0389, p < 0.01 |
| 70–79 years | | | | | | |
| ≥14 days after dose 1 | 765 / 6,260 | 36,481 / 108,451 | 69 (79–55) | $I^2$ = 94%, $\tau^2$ = 0.1285, p < 0.01 | 83 (88–76) | $I^2$ = 86%, $\tau^2$ = 0.1218, p < 0.01 |
| ≥7 days after dose 2 | 178 / 5,673 | 52,158 / 124,128 | 95 (97–90) | $I^2$ = 95%, $\tau^2$ = 0.3267, p < 0.01 | 99 (99–97) | $I^2$ = 95%, $\tau^2$ = 0.8140, p < 0.01 |
| ≥80 years | | | | | | |
| ≥14 days after dose 1 | 765 / 6,260 | 36,481 / 108,451 | 64 (81–34) | $I^2$ = 98%, $\tau^2$ = 0.3725, p < 0.01 | 73 (81–61) | $I^2$ = 90%, $\tau^2$ = 0.1220, p < 0.01 |
| ≥7 days after dose 2 | 178 / 5,673 | 52,158 / 124,128 | 92 (96–86) | $I^2$ = 96%, $\tau^2$ = 0.3968, p < 0.01 | 96 (98–92) | $I^2$ = 91%, $\tau^2$ = 0.4413, p < 0.01 |
| **Sex** | | | | | | |
| Females | | | | | | |
| ≥14 days after dose 1 | 1,267 / 15,767 | 155,536 / 937,579 | 65 (77–46) | $I^2$ = 97%, $\tau^2$ = 0.1846, p < 0.01 | 85 (89–80) | $I^2$ = 94%, $\tau^2$ = 0.1047, p < 0.01 |
| ≥7 days after dose 2 | 352 / 14,852 | 357,374 / 1,139,417 | 94 (96–90) | $I^2$ = 96%, $\tau^2$ = 0.2250, p < 0.01 | 98 (99–97) | $I^2$ = 97%, $\tau^2$ = 0.6326, p < 0.01 |
| Males | | | | | | |
| ≥14 days after dose 1 | 1,629 / 19,665 | 138,492 / 758,081 | 54 (70–30) | $I^2$ = 98%, $\tau^2$ = 0.1859, p < 0.01 | 81 (86–76) | $I^2$ = 93%, $\tau^2$ = 0.0697, p < 0.01 |
| ≥7 days after dose 2 | 447 / 18,483 | 239,427 / 859,016 | 92 (96–86) | $I^2$ = 98%, $\tau^2$ = 0.3486, p < 0.01 | 98 (99–96) | $I^2$ = 98%, $\tau^2$ = 0.7320, p < 0.01 |
| **Presence of any comorbidity** | | | | | | |
| Yes | | | | | | |
| ≥14 days after dose 1 | 2,570 / 25,151 | 149,773 / 631,585 | 59 (73–38) | $I^2$ = 99%, $\tau^2$ = 0.1731, p < 0.01 | 80 (85–73) | $I^2$ = 96%, $\tau^2$ = 0.0880, p < 0.01 |
| ≥7 days after dose 2 | 694 / 23,275 | 242,901 / 724,713 | 93 (96–87) | $I^2$ = 98%, $\tau^2$ = 0.2964, p < 0.01 | 98 (99–95) | $I^2$ = 98%, $\tau^2$ = 0.5856, p < 0.01 |
| No | | | | | | |
| ≥14 days after dose 1 | 326 / 10,281 | 201,255 / 1,121,075 | 82 (89–71) | $I^2$ = 94%, $\tau^2$ = 0.2178, p < 0.01 | 91 (94–87) | $I^2$ = 90%, $\tau^2$ = 0.1335, p < 0.01 |
| ≥7 days after dose 2 | 105 / 10,060 | 353,900 / 1,273,720 | 97 (98–95) | $I^2$ = 88%, $\tau^2$ = 0.2499, p < 0.01 | 99 (100–98) | $I^2$ = 92%, $\tau^2$ = 0.5062, p < 0.01 |
| **Vaccine product** | | | | | | |
| BNT162b2 | | | | | | |
| ≥14 days after dose 1 | 2,215 / 34,751 | 252,380 / 1,654,012 | 58 (73–35) | $I^2$ = 99%, $\tau^2$ = 0.1855, p < 0.01 | 83 (87–77) | $I^2$ = 96%, $\tau^2$ = 0.0865, p < 0.01 |
| ≥7 days after dose 2 | 534 / 33,070 | 390,671 / 1,792,303 | 93 (96–89) | $I^2$ = 98%, $\tau^2$ = 0.2676, p < 0.01 | 98 (99–96) | $I^2$ = 99%, $\tau^2$ = 0.7192, p < 0.01 |
| mRNA-1273 | | | | | | |
| ≥14 days after dose 1 | 413 / 32,949 | 68,157 / 1,469,789 | 70 (82–51) | $I^2$ = 95%, $\tau^2$ = 0.2484, p < 0.01 | 88 (92–82) | $I^2$ = 92%, $\tau^2$ = 0.1694, p < 0.01 |
| ≥7 days after dose 2 | 152 / 32,688 | 109,328 / 1,510,960 | 93 (96–87) | $I^2$ = 93%, $\tau^2$ = 0.3531, p < 0.01 | 98 (99–96) | $I^2$ = 96%, $\tau^2$ = 0.7751, p < 0.01 |
| BNT162b2/mRNA-1273 | | | | | | |
| ≥14 days after dose 1 | - | - | - | - | - | - |
| ≥7 days after dose 2 | 230 / 13,642 | 212,989 / 1,614,621 | 95 (99–85) | $I^2$ = 97%, $\tau^2$ = 1.3097, p < 0.01 | 98 (99–97) | $I^2$ = 78%, $\tau^2$ = 0.2082, p < 0.01 |
| ChAdOx1 | | | | | | |
| ≥14 days after dose 1 | 268 / 39,618 | 30,482 / 1,939,010 | 60 (72–41) | $I^2$ = 84%, $\tau^2$ = 0.1227, p < 0.01 | 81 (86–75) | $I^2$ = 80%, $\tau^2$ = 0.0771, p < 0.01 |
| ≥7 days after dose 2 | 19 / 20,246 | 13,355 / 1,921,883 | 85 (91–75) | $I^2$ = 64%, $\tau^2$ = 0.1585, p = 0.04 | 97 (99–91) | $I^2$ = 91%, $\tau^2$ = 0.9271, p < 0.01 |
| ChAdOx1/mRNA mixed schedule | | | | | | |
| ≥14 days after dose 1 | - | - | - | - | - | - |
| ≥7 days after dose 2 | 6 / 13,421 | 25,182 / 1,426,814 | 97 (99–95) | $I^2$ = 47%, $\tau^2$ = 0.1633, p = 0.15 | 99 (100–98) | $I^2$ = 67%, $\tau^2$ = 0.3962, p = 0.05 |
| **SARS-CoV-2 lineage** | | | | | | |
| Non-VOC | | | | | | |
| ≥14 days after dose 1 | 125 / 8,290 | 181,262 / 1,075,998 | 85 (96–41) | $I^2$ = 99%, $\tau^2$ = 1.4743, p < 0.01 | 66 (86–14) | $I^2$ = 95%, $\tau^2$ = 0.6239, p < 0.01 |
| ≥7 days after dose 2 | 33 / 1,387 | 230,867 / 818,862 | 99 (100–91) | $I^2$ = 97%, $\tau^2$ = 3.0144, p < 0.01 | 92 (96–84) | $I^2$ = 57%, $\tau^2$ = 0.2131, p = 0.10 |
| Alpha (B.1.1.7) | | | | | | |
| ≥14 days after dose 1 | 1,372 / 11,171 | 351,028 / 1,712,601 | 30 (40–19) | $I^2$ = 77%, $\tau^2$ = 0.0189, p < 0.01 | 85 (88–81) | $I^2$ = 79%, $\tau^2$ = 0.0441, p < 0.01 |

| | | | | | | |
|---|---|---|---|---|---|---|
| ≥7 days after dose 2 | 67 / 9866 | 596,801 / 1,488,207 | 98 (99–96) | $I^2$ = 84%, $\tau^2$ = 0.3136, p < 0.01 | 98 (99–95) | $I^2$ = 74%, $\tau^2$ = 0.2842, p < 0.01 |
| Beta/Gamma (B.1.3.51 or P.1) | | | | | | |
| ≥14 days after dose 1 | >50 / 480 | 181,262 / 1,075,998 | -[b] | $I^2$ = 85%, $\tau^2$ = 0.6820, p < 0.01 | 81 (86–74) | $I^2$ = 0%, $\tau^2$ = 0, p = 0.75 |
| ≥7 days after dose 2 | >0 / 203 | 230,867 / 818,862 | -[c] | | -[c] | |
| Beta (B.1.351) | | | | | | |
| ≥14 days after dose 1 | >19 / 196 | 181,262 / 1,075,998 | -[b] | $I^2$ = 75%, $\tau^2$ = 0.9788, p = 0.02 | 63 (78–37) | $I^2$ = 0%, $\tau^2$ = 0, p = 0.92 |
| ≥7 days after dose 2 | >0 / >3 | 230,867 / 818,862 | -[c] | | -[c] | |
| Gamma (P.1) | | | | | | |
| ≥14 days after dose 1 | >168 / 1,034 | 181,262 / 1,075,998 | 6 (45 to -61) | $I^2$ = 85%, $\tau^2$ = 0.1542, p < 0.01 | 76 (80–71) | $I^2$ = 0%, $\tau^2$ = 0, p = 0.99 |
| ≥7 days after dose 2 | >4 / >467 | 230,867 / 818,862 | 98 (99–96) | $I^2$ = 0%, $\tau^2$ = 0, p = 0.58 | 96 (99–86) | $I^2$ = 58%, $\tau^2$ = 0.4345, p = 0.12 |
| Delta (B.1.617.2) | | | | | | |
| ≥14 days after dose 1 | 321 / 3223 | 194778 / 1,096,618 | 74 (94–-7) | $I^2$ = 99%, $\tau^2$ = 2.0844, p < 0.01 | 89 (95–77) | $I^2$ = 94%, $\tau^2$ = 0.5667, p < 0.01 |
| ≥7 days after dose 2 | 417 / 3319 | 559071 / 1,460,911 | 89 (98–35) | $I^2$ = 100%, $\tau^2$ = 3.1280, p < 0.01 | 99 (99–97) | $I^2$ = 98%, $\tau^2$ = 0.6547, p < 0.01 |
| **Interval between doses** | | | | | | |
| **0–6 days after dose 2** | | | | | | |
| 21–34 days | >19 / 32,555 | 3,155 / 1,404,787 | 79 (87–66) | $I^2$ = 3%, $\tau^2$ = 0.0133, p = 0.31 | 94 (99–38) | $I^2$ = 83%, $\tau^2$ = 2.4291, p = 0.02 |
| 35–55 days | >10 / 32,551 | 5,878 / 1,407,510 | 87 (95–66) | $I^2$ = 74%, $\tau^2$ = 0.7026, p < 0.01 | 91 (95–86) | $I^2$ = 0%, $\tau^2$ = 0, p = 0.51 |
| 56–83 days | >12 / 13,425 | 9,359 / 1,104,250 | 89 (94–77) | $I^2$ = 56%, $\tau^2$ = 0.2723, p = 0.08 | 94 (96–89) | $I^2$ = 12%, $\tau^2$ = 0.0663, p = 0.33 |
| ≥84 days | >16 / 32,553 | 6,444 / 1,408,076 | 80 (94–30) | $I^2$ = 80%, $\tau^2$ = 1.2881, p < 0.01 | 93 (98–79) | $I^2$ = 70%, $\tau^2$ = 0.8055, p = 0.02 |
| **7–55 days after dose 2** | | | | | | |
| 21–34 days | 29 / 32,627 | 9,410 / 1,104,301 | 84 (89–78) | $I^2$ = 44%, $\tau^2$ = 0.0439, p = 0.15 | 94 (96–90) | $I^2$ = 71%, $\tau^2$ = 0.2059, p = 0.02 |
| 35–55 days | 29 / 13,441 | 37,571 / 1,132,462 | 97 (99–89) | $I^2$ = 94%, $\tau^2$ = 1.3166, p < 0.01 | 98 (99–94) | $I^2$ = 86%, $\tau^2$ = 1.1059, p < 0.01 |
| 56–83 days | 43 / 13,455 | 109,802 / 1,204,693 | 96 (98–94) | $I^2$ = 74%, $\tau^2$ = 0.2187, p = 0.01 | 99 (99–97) | $I^2$ = 88%, $\tau^2$ = 0.4462, p < 0.01 |
| ≥84 days | >48 / 13,463 | 38,731 / 1,133,622 | 89 (96–69) | $I^2$ = 94%, $\tau^2$ = 0.9692, p < 0.01 | 98 (99–94) | $I^2$ = 94%, $\tau^2$ = 0.9409, p < 0.01 |
| **56–111 days after dose 2** | | | | | | |
| 21–34 days | >40 / 32,581 | 15,620 / 1,417,252 | 91 (96–79) | $I^2$ = 79%, $\tau^2$ = 0.5797, p < 0.01 | 97 (98–96) | $I^2$ = 0%, $\tau^2$ = 0, p = 0.63 |
| 35–55 days | 34 / 32,570 | 61,129 / 1,462,761 | 97 (99–93) | $I^2$ = 89%, $\tau^2$ = 0.7648, p < 0.01 | 99 (99–98) | $I^2$ = 73%, $\tau^2$ = 0.3612, p = 0.01 |
| 56–83 days | >135 / 32,675 | 129,501 / 1,531,133 | 95 (98–89) | $I^2$ = 96%, $\tau^2$ = 0.5262, p < 0.01 | 99 (100–98) | $I^2$ = 96%, $\tau^2$ = 0.8204, p < 0.01 |
| ≥84 days | >127 / 32,663 | 46,942 / 1,448,574 | 86 (96–48) | $I^2$ = 98%, $\tau^2$ = 1.3642, p < 0.01 | 99 (100–95) | $I^2$ = 98%, $\tau^2$ = 1.3064, p < 0.01 |
| **≥112 days after dose 2** | | | | | | |
| 21–34 days | >7 / 13,424 | 5,248 / 1,100,139 | 86 (94–65) | $I^2$ = 80%, $\tau^2$ = 0.6880, p < 0.01 | 97 (99–92) | $I^2$ = 71%, $\tau^2$ = 0.5472, p = 0.01 |
| 35–55 days | >24 / 32,563 | 12,805 / 1,414,437 | 89 (97–66) | $I^2$ = 90%, $\tau^2$ = 0.9139, p < 0.01 | 97 (98–95) | $I^2$ = 19%, $\tau^2$ < 0.0001, p = 0.29 |
| 56–83 days | >13 / 13,425 | 2,522 / 1,097,413 | 77 (95 to -10) | $I^2$ = 89%, $\tau^2$ = 1.7106, p < 0.01 | 97 (99–95) | $I^2$ = 21%, $\tau^2$ = 0.0609, p = 0.28 |
| ≥84 days | >4 / >13,416 | 11,450 / 1,413,082 | 97 (99–92) | $I^2$ = 6%, $\tau^2$ = 0.1071, p = 0.34 | 99 (100–97) | $I^2$ = 61%, $\tau^2$ = 0.9402, p = 0.08 |

[a]Models adjusted for age group, sex, public health unit region, biweekly period of test, number of SARS-CoV-2 tests in the 3 months prior to 14 December 2020, presence of any comorbidity that increase the risk of severe COVID-19, receipt of influenza vaccination in current or prior influenza season, and neighbourhood income (material deprivation index in British Columbia), proportion of persons employed as non-health essential workers, persons per dwelling, and self-identified visible minority quintiles. Receipt of influenza vaccination was not adjusted for in British Columbia because of lack of data.

[b]VE not reported due to extremely imprecise 95% confidence intervals.

[c]Not applicable as VE estimate from only one province available.
